# Heterogeneity of use, access and retention of insecticide-treated nets: implications for subnational tailoring to maximise malaria control

**DOI:** 10.1101/2025.08.27.25334550

**Authors:** Andrew C Glover, Hannah Koenker, El Hadji Amadou Niang, Kate Kolaczinski, Thomas S Churcher

## Abstract

Insecticide-treated nets (ITNs) are the most impactful and cost-effective control tool against malaria. ITNs are primarily distributed through triennial mass campaigns across Africa, though overall ITN use remains modest in many areas as most ITNs do not last three years. In times of funding constraints and a lack of economic alternative antimalarial interventions it is unclear whether disease control could be best improved by distributing more effective ITNs (e.g. dual active-ingredient ITNs) and/or deploying nets more frequently. There are increased calls to improve allocation of resources through sub-national tailoring of interventions, though benefits will depend on how long people use ITNs and how this varies between subnational regions. However, subnational variation in ITN retention and the duration that ITNs remain in use have not previously been quantified. Here we estimate subnational differences in ITN use, access and retention for six countries, Burkina Faso, Ghana, Malawi, Mali, Mozambique and Senegal. These estimates are used to calibrate a *Plasmodium falciparum* transmission dynamics model to generate sub-national estimates of ITN use and cases averted under different ITN distribution strategies. On average, people use their ITNs for 21 months, though this varies substantially between subnational regions from 12 to 38 months. Shifting from triennial to biennial campaigns is predicted to lead to mean population-weighted use across all regions increasing from 45.4% to 53.9%. No regions of the 146 investigated were estimated to maintain use over 80% even under biennial distribution, though switching to dual active-ingredient ITNs would likely avert more cases under present distribution frequencies. Our results highlight that although transmission intensity remains an important factor for subnational tailoring of malaria control interventions, other factors, such as ITN use given access, meaningfully influence optimal deployment strategies. The framework highlights how routinely collected data can aid policymakers in tailoring disease control programmes at subnational levels.

## Introduction

The widespread use of insecticide-treated nets (ITNs) has prevented more malaria than any other intervention. For populations at risk of malaria, national malaria programmes (NMPs) have historically aimed to achieve ‘universal coverage’ of ITNs, which is defined as a measure of both universal access to, and use of, ITNs (***World Health Organization, 2014***). The 2005-2015 ***Roll Back Malaria Partnership (2005***) Global Strategic Plan first proposed universal coverage targets of 80% of people at risk of malaria by 2010 (***Roll Back Malaria Partnership, 2008; Smith et al., 2009***). The ***World Health Organization (2024b***) Guidelines for Malaria recommended achieving universal coverage by implementing mass distribution campaigns, and through the use of complimentary continuous distribution channels; these are typically conducted via antenatal care (ANC), the Expanded Programme for Immunization (EPI) and through school-based distribution (***World Health Organization, 2024b; Koenker et al., 2022***). Mass campaigns have historically been recommended to be conducted every three years with a defined number of nets distributed per person (***World Health Organization, 2024b; Koenker et al., 2022***). These policies are estimated to have increased ITN use across sub-Saharan Africa from median point estimates of approximately 2% to 49% between 2000 to 2021 (***Bertozzi-Villa et al., 2021***). Access to a net (assuming an ITN could be used by up to two people) has also similarly been estimated to have increased from median estimates of 3% to 56% over the same period (***Bertozzi-Villa et al., 2021***). While this falls short of the original recommendation, ITNs have been instrumental in reducing the global malaria burden, having averted an estimated 450 million cases between 2000 and 2015 (***Bhatt et al., 2015***), or two-thirds of all cases averted through preventative interventions over the same period. Nevertheless, the disparity between recent estimates of ITN use and the original 80% targets are stark. High population use may be unachievable with the status quo of triennial campaigns, since it is thought people do not use their nets for three years, with previous median estimates of national median ITN retention times being lower in 87.5% of sub-Saharan African countries (***Bertozzi-Villa et al., 2021***). There is also likely to be substantial subnational variability due to environmental and behavioural differences, though evidence is lacking due to the sporadic nature of net surveillance activities. Since 2019, WHO good practice statements have shifted from advocating for ‘universal’ to ‘optimal’ coverage guided by “an explicit prioritization process guiding resource allocation decisions” (***World Health Organization, 2024b***). However, it is unclear what the optimum coverage is, how it can be estimated, and how this might vary with net retention time.

The epidemiological impact of mass distributions of ITNs will depend on how long individuals retain and sleep under nets and how effective ITNs are at killing mosquitoes and inhibiting blood-feeding. The original rationale for triennial mass campaigns is unclear and is likely based on the durability of first-generation ITNs treated solely with pyrethroid-class insecticides (***World Health Organization, 2005, 2013***). The real-world effectiveness of pyrethroid-based ITNs is under threat from increasingly high levels of pyrethroid resistance in anopheline mosquitoes across sub-Saharan Africa, which is highly heterogenous over national and sub-national scales (***Coleman et al., 2017; Moyes et al., 2020; Hancock et al., 2020, 2022***). Experimental hut trials have indicated increased pyrethroid resistance in the vector population not only reduces the initial mortality conferred by new ITNs, but also increases the rate they lose their insecticidal properties following distribution (***Nash et al., 2021; Sherrard-Smith et al., 2022; Churcher et al., 2024***). WHO recommendations suggest that more regular mass campaigns or altering the distribution ratio might be appropriate given local data-driven evidence (***World Health Organization, 2024b; Koenker et al., 2022***). Nevertheless, international procurers of ITNs, such as the Global Fund, have not historically advised altering the regularity of ITN mass campaigns, nor is this considered in sub-national tailoring exercises, despite this potentially being more cost-effective than layering multiple interventions in the same region.

The World Health Organization has recently expanded its guidance on different types of ITN, with three classes now recommended in areas of pyrethroid resistance. WHO recommendations cascade in the following order: i) pyrethroid-chlorfenapyr nets are strongly recommended for use in place of the standard pyrethroid-only nets; ii) pyrethroid-piperonyl butoxide (PBO) treated nets are conditionally recommended in favour of the standard pyrethroid-only nets, and iii) pyrethroid-pyriproxifen (PPF) treated nets are conditionally recommended in favour of pyrethroid-only nets, but are recommended not to be used in place of pyrethroid-PBO nets (***World Health Organization, 2024b***). These recommendations are based on results from cluster randomised controlled trials showing that pyrethroid-PBO (***Protopopoff et al., 2018; Staedke et al., 2020***) and pyrethroidchlorfenapyr (***Accrombessi et al., 2023; Churcher et al., 2024***) products can both mitigate pyrethroid-resistance associated decreases in ITN effectiveness, while superiority trials for pyrethroid-PPF nets showed mixed results (***Accrombessi et al., 2023***). The durability of different classes of nets is also thought to vary substantially, further exacerbating the need to consider the optimum regularity of mass campaigns.

NMPs therefore face two key challenges for planning future ITN interventions. Firstly, to increase ITN use, more frequent mass campaigns may be necessary, given ITNs are typically retained for shorter durations than the status quo mass campaign interval of three years. Secondly, in Africa, where pyrethroid resistance is widespread, newer, yet costlier ITN products are advisable. Addressing either of these challenges would require increases in funding, yet budgets are increasingly under threat and there are increasing calls to tailor ITN distribution within countries to prevent waste and maximise benefit. This is recommended by the WHO (***World Health Organization, 2024a***) but has not yet been implemented at scale. The impact of both these policy choices may not only depend on how access to ITNs, pyrethroid resistance and transmission intensity vary between and within countries, but may also depend on how likely individuals with access to ITNs are to use them, in addition to the duration that ITNs continue to be used. It is unclear which of these factors are likely to be most associated with improvements in health outcomes through more regular mass campaigns, or more effective ITNs. Moreover, it is unknown if net retention varies systematically within a country or if ITNs remain in use for shorter durations than they are retained for.

To investigate optimal ITN deployment, we estimate the mean duration of ITN access and use from Demographic and Health Survey (DHS) data for different subnational regions in six African countries (Burkina Faso, Ghana, Malawi, Mali, Mozambique and Senegal). Here, (subnational) regions are defined as the first administrative unit below the country level and are further divided into rural and urban areas to align with DHS stratification. A bespoke mechanistic statistical model is fitted to subnational DHS data from 2005 to 2024. It allows regions with sporadic DHS surveys to be directly compared and generates estimates of ITN use, access and use given access over time, in addition to the contribution towards these by ITNs distributed through continuous channels (ITNs obtained through the private sector, any school- or community-based distribution programme, or through routine ANC and EPI channels). Projections are made for how these metrics change from shifting from triennial to biennial mass campaigns. Using these subnational estimates we calibrated individual-based models of *Plasmodium falciparum* malaria transmission to generate subnational projections of clinical cases averted over 2025 to 2030 under different ITN distribution strategies. Our findings should enable assessment of the epidemiological benefit of different ITN strategies and provide data-driven evidence to improve ITN priorisation guidance.

## Results

### Retention time and duration of use

The model estimates people keep and use ITNs for considerably less than three years. Across all regions in all six countries studied, the population-weighted mean duration of ITN access, herein referred to as ITN retention time, was 26.8 months (95% CrI: 26.7, 26.9). Retention times were estimated to vary notably between and within countries (figure 1), with median estimates ranging from 14.8 months (95% CrI: 14.5, 15.1) in rural Tete, Mozambique, to 41.5 months (95% CrI: 40.7, 42.2) in rural Ségou, Mali, with only 23 of 146 (15.8%) regions estimated to have a mean retention time greater than three years. The mean durations of ITN use similarly varied notably both subnationally and between countries and were estimated to be consistently lower than mean retention times (figure 1), with a population-weighted mean across all regions of 21.0 months (95% CrI: 20.9, 21.0). The mean duration that ITNs remain in use was also estimated to be shortest in rural Tete with a median estimate of 12.0 months (95% CrI: 11.8, 12.3). Meanwhile, the longest mean duration of use was in urban Est, Burkina Faso (figure 5 – figure supplement 1.A) at 37.8 months (95% CrI: 34.9, 40.6), which was the only region (0.7%) with a median estimate greater than three years. Caution is needed when comparing retention times between countries, due to differences in the timings of surveys relative to mass campaigns, but these are likely to be minimised over subnational scales within a country. While no systematic differences in the durations of use or access were estimated between urban and rural regions in Senegal, differences were observed in some other countries, such as in Ghana, where values were estimated to be broadly lower in urban settings (figure 5 – figure supplement 2.A). For most regions, the gap between use and access was estimated to widen over time following mass campaigns, indicating people stop using ITNs substantially before they stop owning them, so that use given access is not constant but typically wanes as ITNs age (figure 2.A).

**Figure 1.**
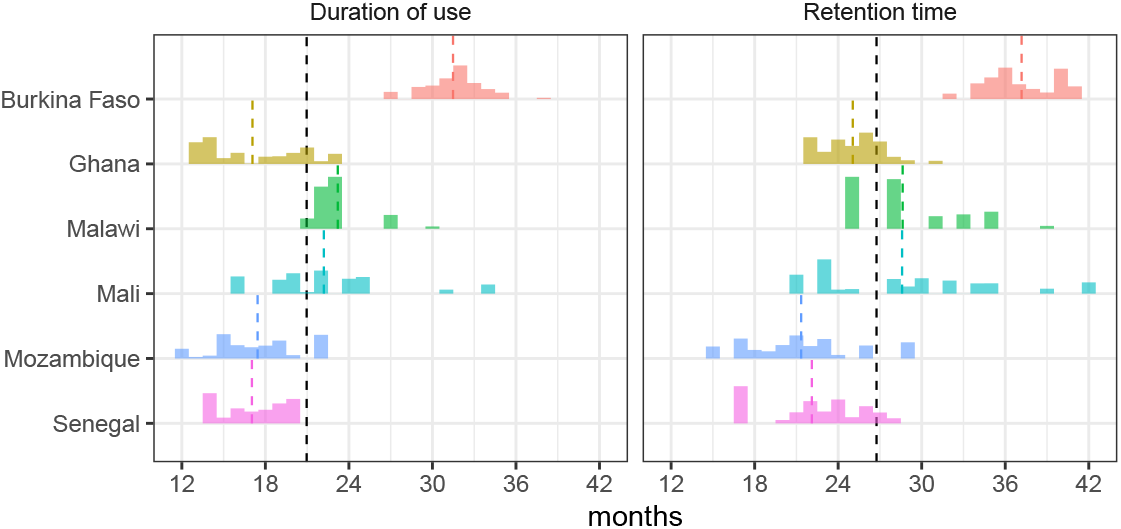
Population-weighted distribution of expected ITN retention times and duration of use by country. Median estimates of the mean ITN retention time and duration of use for each subnational region were weighted by population to produce country-level distributions. These histograms represent probability distributions of retention time and duration of use for an individual randomly selected from each country, assuming model-derived median estimates for all subnational regions. Subnational variability in retention times and the duration of use reflects differences in model-derived estimates between regions, since the model assumes these values are constant within regions. Vertical dashed lines indicate the overall population-weighted mean (black) and the population-weighted mean for each country (coloured).

**Figure 2.**
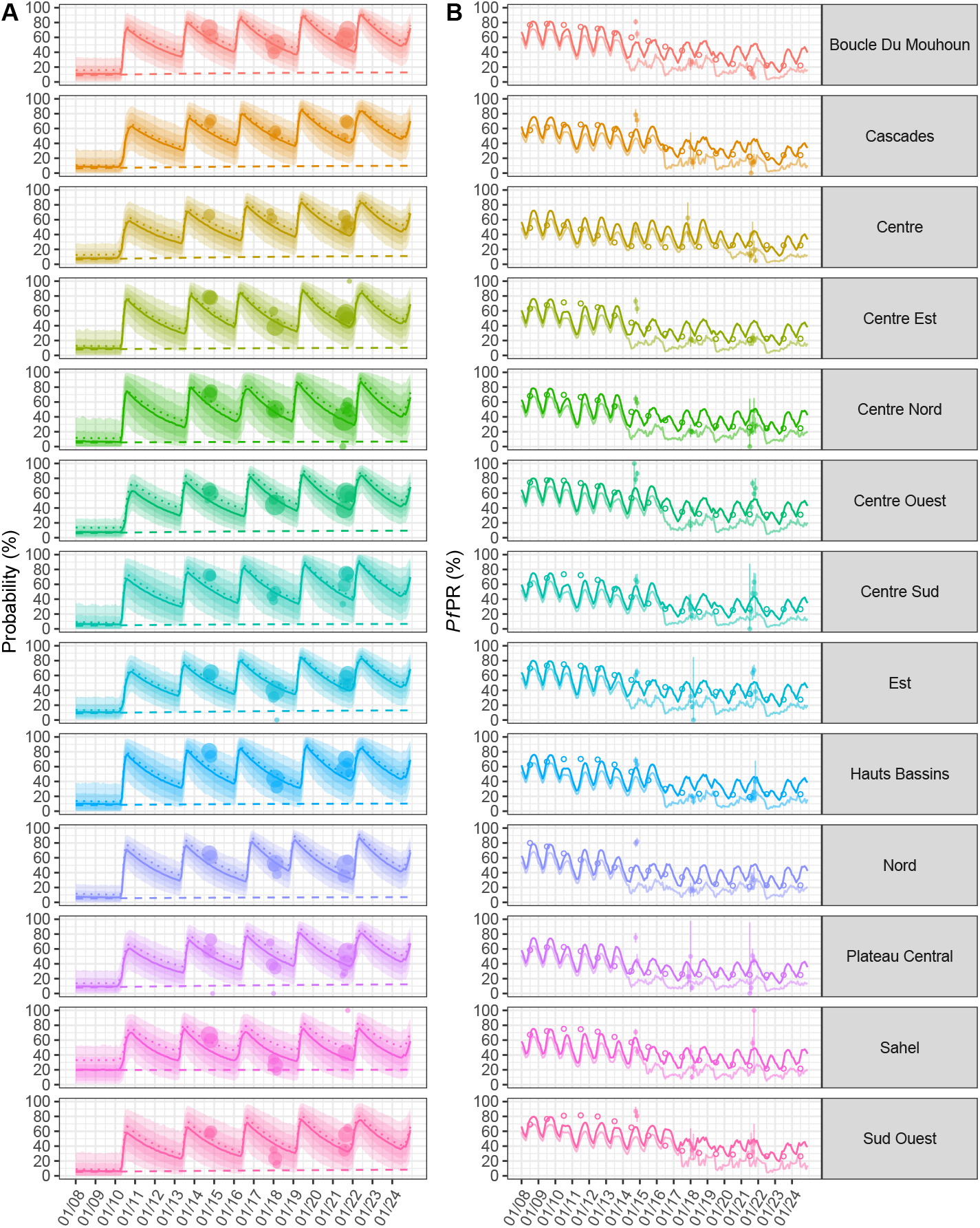
ITN use, access and *Pf* PR over time in rural Burkina Faso. **A**: Survey estimates of ITN use (probability someone slept under an ITN the previous night, points sized proportionally to sample size) and model mean estimates of the probability of use over time (solid lines) for rural portions of subnational regions in Burkina Faso. Mean estimates are shown for the probability of access (dotted lines) and the probability of using an ITN sourced from a continuous channel (dashed lines).The model accounts for differing probabilities of use between individuals within a region; this results in notable uncertainty around the probability an individual selected at random uses a ITN. This uncertainty is illustrated in the shaded region, with the 50%, 80% and 95% credible intervals as indicated by progressively lighter shaded regions. **B**: *Pf* PR_2−10_ estimates from the transmission dynamics model (darker lines) were fitted to annual estimates of *Pf* PR_2−10_ from the ***Malaria Atlas Project (2024***) (hollow points); model estimates of *Pf* PR_6−59mo_ are also shown (lighter lines), in addition to observed DHS *Pf* PR_6−59mo_ and associated 95% credible intervals due to measurement uncertainty (solid points and vertical lines). **Figure 2—figure supplement 1.** Use, access and *Pf* PR over time in rural Senegal. **Figure 2—figure supplement 2.** Use of ITNs from continuous channels.

### Universal coverage: was it achievable under triennial mass campaigns?

Access and use of ITNs are estimated to have varied substantially over time, both within and between countries (figure 2.A and 2 - figure supplement 1.A). Inferring changes over time from observational data alone can give misleading results. For example, in rural Centre-Nord in Burkina Faso, DHS survey data suggests a sustained decrease in use between 2014 and 2022 (figure 2.A); however, the model which considered the timings of mass campaigns and routine ITN distribution estimates that overall ITN use immediately following mass campaigns increased over this period. The observed decrease in use and access across many regions in Burkina Faso may therefore be a by-product of DHS surveys being conducted at progressively later dates relative to the most recent campaign; this does not necessarily indicate an underlying trend in decreasing use or access over longer timescales. This highlights the potential limitations of making direct comparisons between different surveys, which may be conducted at different intervals since the preceding campaign (figure 2.A). This waxing and waning dynamics in use over time is also observed in *P. falciparum* prevalence (figure 2.B). In many countries peak ITN use and access following mass campaigns has increased over time; for example in rural Thiès in Senegal, the overall use immediately following the first 2009 mass campaign was estimated to reach 18%, yet peak levels of use above 80% were estimated to be achieved following all mass campaigns since 2016 (figure 2 - figure supplement 1.A). Similar trends can be seen in the context of access (dotted lines, figure 2.A and figure 2 - figure supplement 1.A).

Mean ITN use over the three years following a campaign is estimated to be modest, with a population-weighted mean of 45.4% (95% CrI: 45.2, 45.5) across all regions studied. Overall access to an ITN was estimated to be greater over the three year period, with 58.1% (95% CrI: 58.0, 58.2) of people having access to an ITN, on average. The population-weighted mean use given access across all regions was estimated to be 75.7% (95% CrI: 75.5, 76.0). While no regions were estimated to have a mean use or access greater than 80% over either the three-year or two-year periods following a mass campaign, in the context of use given access, 46% of subnational regions had median estimates greater than 80% over a three-year period following a mass campaign, increasing to 70% of regions when only the first two years were considered (figure 3 and figure 3 – figure supplements 1–3). Nevertheless, some regional differences are observable, with southern Malawi having a low overall ITN use despite the region having a similar level of access to the rest of the country. Across all regions, mass campaigns are estimated to increase ITN use in their immediate aftermath to greater than 80% in 76.8% of regions (bottom row, figure 3 and figure 3 – figure supplements 1–3), with considerable variability within and between countries. For example, for rural areas in Burkina Faso, the northern Sahel region is predicted to never achieve ITN use above 80%, whereas use is predicted to be maintained above 80% for 6 months following a mass campaign in the south-western region of Cascades.

**Figure 3.**
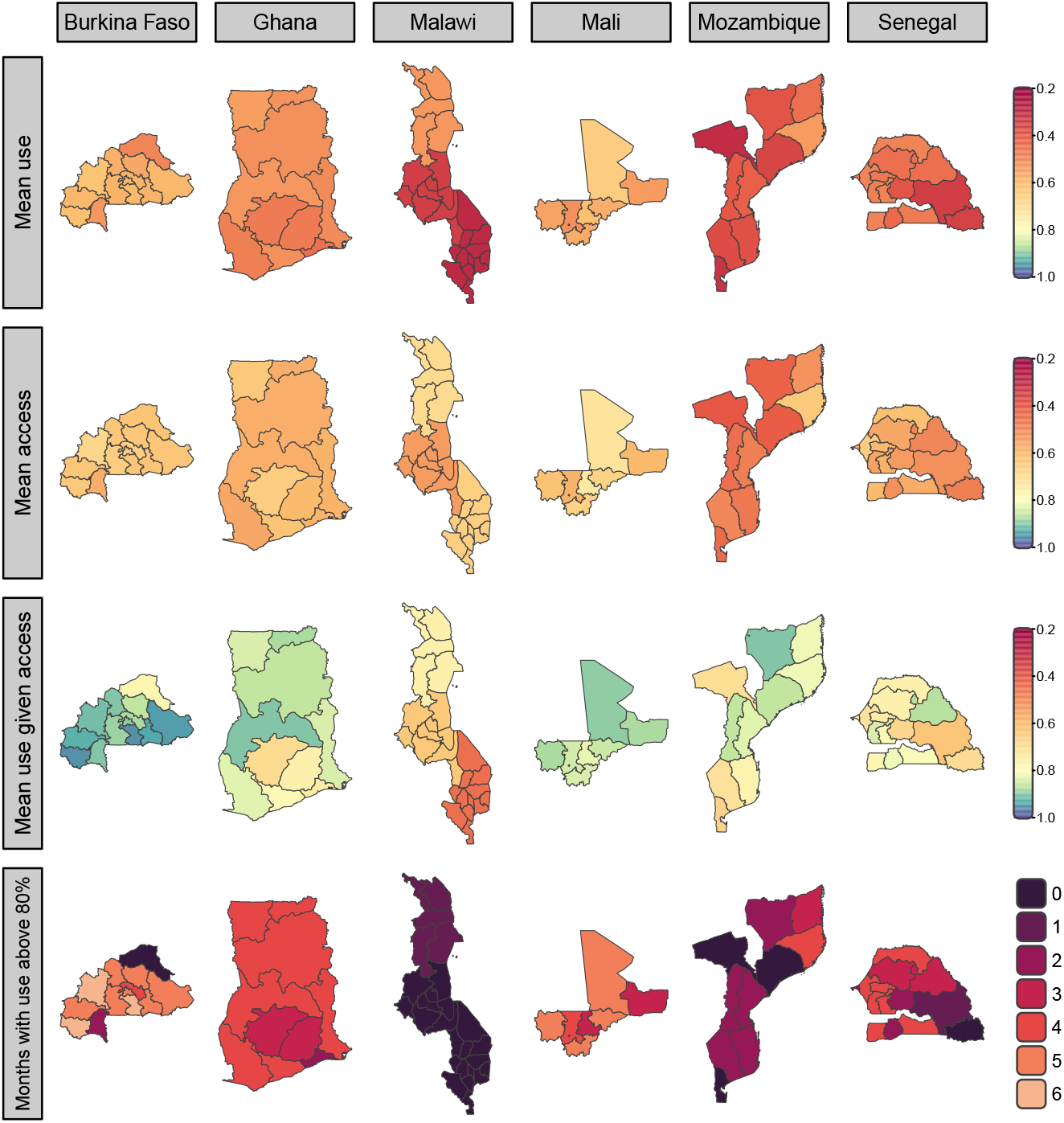
Mean use and access with three-year campaigns in subnational rural areas. Central estimates of mean overall proportion of people using an ITN the previous night, access to an ITN, and use given access for three-year mass campaign intervals are shown in the top three rows. The number of months following a mass campaign where overall ITN use exceeds 80% are shown in the bottom row. **Figure 3—figure supplement 1. Mean use and access with three-year campaigns in subnational urban areas.** **Figure 3—figure supplement 2. Mean rural use and access with two-year campaigns.** **Figure 3—figure supplement 3. Mean urban use and access with two-year campaigns.**

The contribution of ITNs from continuous channels also differed between and within countries; for example, in rural regions of Thiès (figure 2 figure supplement 1) the use of ITNs from continuous channels was estimated to be 15.8% (95% CrI: 14.7, 17.0) at the end of 2024, in comparison to 4.1% (95% CrI: 3.8,4.5) in Sédhiou. Differences in the use of continuously-distributed ITNs were also notable between countries, with 19.8% (95% CI: 17.4, 22.2) of person-nights of ITN use (one person using an ITN for one night) across Burkina Faso estimated to be attributable to continuouslydistributed ITNs over a three-year period, compared to Malawi at 61.2% (95% CI: 58.3, 64.1) (figure 2 - figure supplement 2.B).

There is considerable variability in observed mean access and use within a region, with notable uncertainty between communities, as indicated by the high uncertainty around the probability an individual randomly selected from the region will use an ITN (figure 1.A). The effect of this is illustrated by rural Thiès and Ziguinchor, which both have a mean overall ITN use of 58% when averaged over three years following the last mass campaign. Though the average is the same across both regions, more people in rural areas of Ziguinchor either have a very low or very high probability of using an ITN, indicating greater inequity in ITN use in rural areas of Ziguinchor than in Thiès (figure 4). A similar trend is also apparent in the context of access and in other settings (figure 4 - figure supplement 1 and 2).

**Figure 4.**
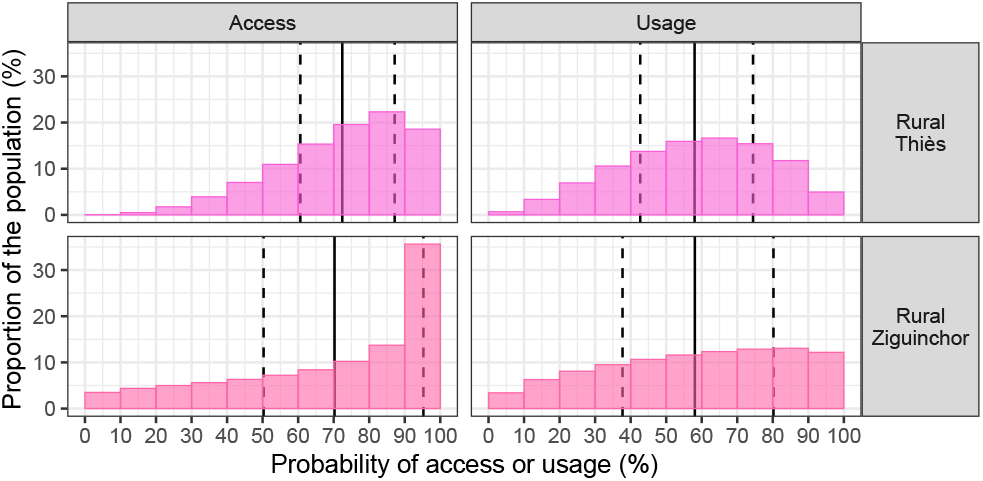
Equity of use and access in rural Thiès and Ziguinchor, Senegal. Solid vertical lines denote overall ITN use and access averaged over three years following the last mass campaign. Coloured bars indicate the proportion of the population with different probabilities of using or having access to an ITN. For example, 10% of the population have a probability of access between 70% and 80% in rural Ziguinchor, in comparison to 20% in rural Thiès. Vertical dashed lines denote 50% credible intervals for the probability of an individual uses or has access to an ITN, indicating that half of the population are expected to have a probability of use or access within this range. **Figure 4—figure supplement 1. Subnational equity of use in Senegal.** **Figure 4—figure supplement 2. Subnational equity of access in Senegal.**

### Improving ITN use

The framework can identify regions of low ITN use and make suggestions about how this could be improved (figure 5.A,B). ITNs will be most effective in areas where people have access to nets for a long period of time and are more likely to use them (quadrant 1, figure 5.B). In areas where people keep ITNs but have low use given access (quadrant 2, figure 5.B) NMPs may wish to consider social and behaviour change (SBC) interventions such as ‘hang-up’ campaigns. Conversely, in areas with lower ITN retention times but high use given access (quadrant 4, figure 5.B) NMPs may wish to consider more regular mass campaigns to improve equality of access between different regions. Both of these policy changes may be warranted in regions with both low retention and use given access to improve overall ITN use (quadrant 3, figure 5.B). Results varied by country (figure 5 – figure supplements 1-5).

**Figure 5.**
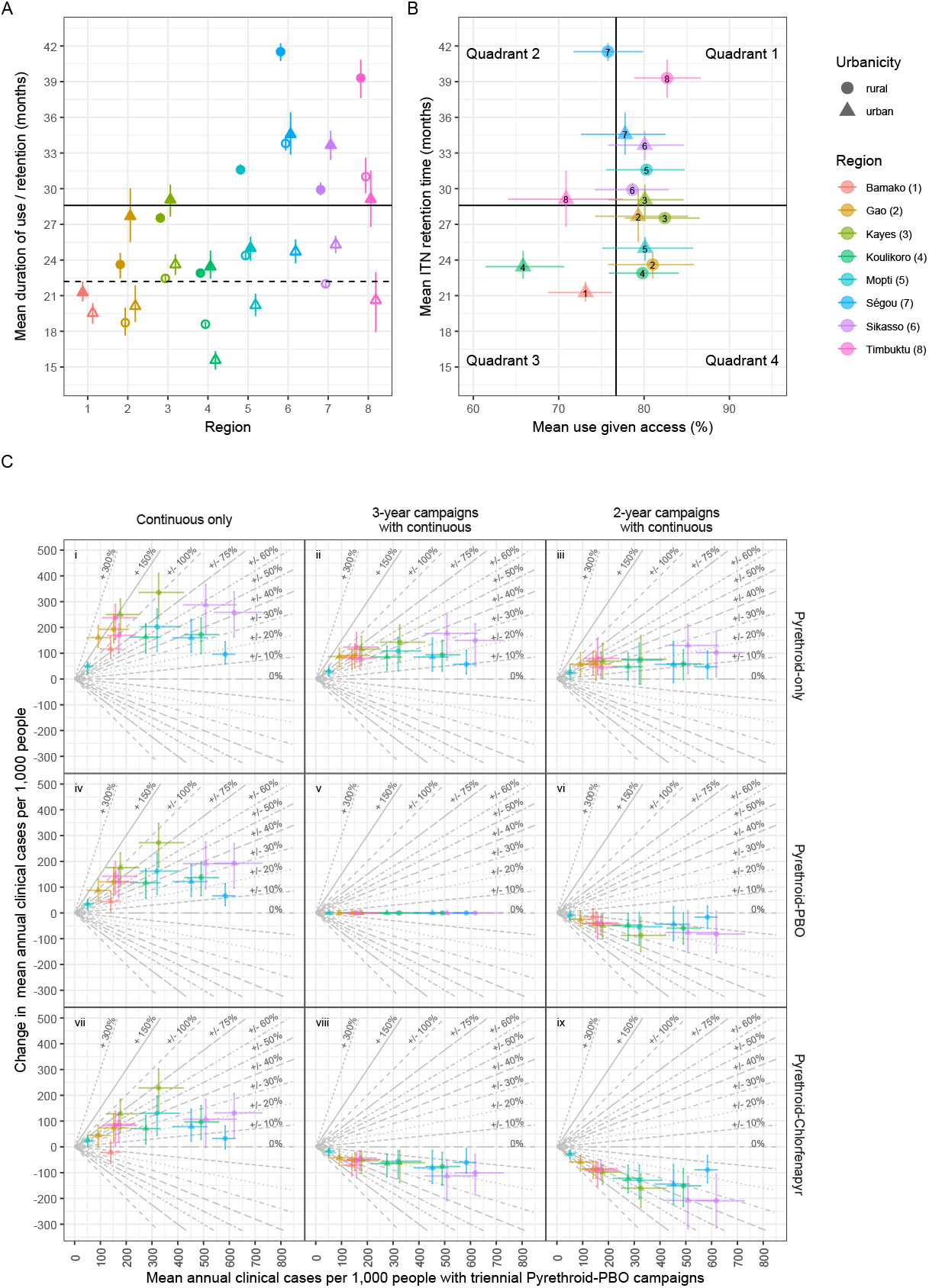
Retention time, use given access and changes in cases for Mali. All coloured points indicate median estimates and associated 95% credible intervals for subnational regions in Mali. **A**: Mean durations of ITN use (hollow) and retention (filled) are shown in addition to national median estimates (black solid and dashed lines, respectively). **B**: Mean ITN retention and use given access under a triennial campaign strategy are shown, with black lines indicating national median estimates. **C**: Points indicate mean annual clinical cases under a triennial pyrethroid–PBO campaign strategy, and the projected change in cases for alternative intervention strategies with pyrethroid-only (**C.i–iii**) pyrethroid-PBO (**C.iv–vi**) or pyrethroid–chlorfenapyr (**C.vii–ix**) ITNs, with continuous-only distribution (**C.i, iv, vii**), or in conjunction with triennial (**C.ii, v, viii**) or biennial (**C.iii, vi, ix**) campaigns. The change in clinical cases is equal to zero for equivalent comparator and intervention strategies (**C.v**). Labeled diagonal reference lines with positive gradients indicate percentage increases in clinical cases relative to mean annual estimates under the comparator strategy; lines of the same style with negative gradients indicate equivalent percentage decreases. Urban areas were estimated to broadly have lower use given access than in rural settings, while the capital city, Bamako, was estimated to have the lowest ITN retention time of all regions. **Figure 5—figure supplement 1. Retention time, use given access and changes in cases for Burkina Faso.** **Figure 5—figure supplement 2. Retention time, use given access, and changes in cases for Ghana.** **Figure 5—figure supplement 3. Retention time and use given access for Malawi.** **Figure 5—figure supplement 4. Retention time, use given access and changes in cases for Mozambique.** **Figure 5—figure supplement 5. Retention time, use given access and changes in cases for Senegal.**

The statistical model can be used to predict the increase in use caused by increasing the frequency of mass campaigns from three to two years, assuming the contribution of continuous distribution channels and the decay in use given access following mass campaigns remains the same. Increases in access are estimated to be modest, with percentage-point population-weighted mean increases across all settings of 8.15% (95% CrI: 8.13, 8.17), from 58.1% (95% CrI: 58.0, 58.2) to 66.3 (95% CrI: 66.2, 66.4)%. Greater increases are predicted to be seen for ITN use, with the population-weighted mean use across all settings increasing from 45.4% (95% CrI: 45.3, 45.5) to 53.9% (95% CrI: 53.7, 54.1), an increase of 8.50% (95% CrI: 8.47, 8.54). An increase in use given access is also projected, with the population-weighted mean increasing by 4.13% (95% CrI: 4.07, 4.20) in absolute terms from 75.7% (95% CrI: 75.5, 76.0) to 79.9% (95% CrI: 79.6, 80.2). Subnational variation is also predicted in changes of access, use and use given access following adoption of biennial mass campaigns (figure 3 and corresponding figure supplements 1-3).

### Improving ITN effectiveness

Policy-makers may wish to optimise ITN distribution to maximise epidemiological impact. We generate estimates of clinical cases under different distribution strategies by simulating halting mass campaigns or changing the campaign interval from three to two years. Fifty eight percent of ITNs distributed in sub-Saharan Africa in 2023 were reported to have been pyrethroid-PBO nets (***World Health Organization, 2024c***); we therefore estimate changes in clinical cases for alternative strategies compared to the current triennial pyrethroid-PBO campaigns with supplementary continuous distribution, with triennial pyrethroid-chlorfenapyr strategies as a comparator also presented (figure 6 - figure supplement 1).

**Figure 6.**
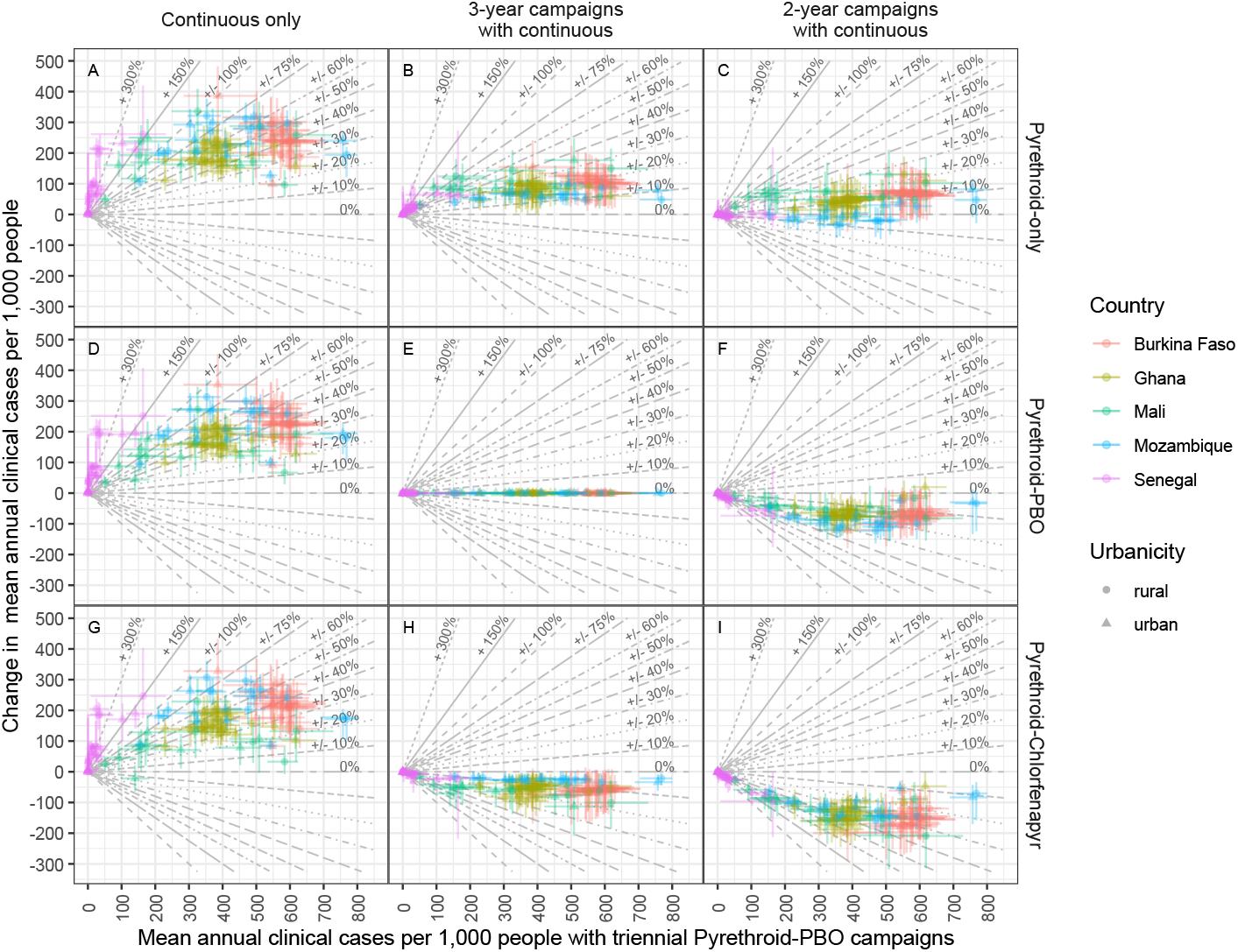
Change in cases vs. cases with triennial pyrethroid-PBO distribution. Points represent urban and rural areas within each subnational region, are colored by country, and are sized proportionally to the mean annual number of ITNs distributed under each strategy. Each point shows median estimates of the change in clinical cases following a switch from triennial pyrethroid-PBO distribution to alternative strategies against the projected clinical cases under the triennial pyrethroid-PBO strategy. Concurrent distribution of continuous ITNs of the same class used in mass campaigns in assumed throughout. Vertical and horizontal lines indicate 95% credible intervals. Labeled diagonal reference lines with positive gradients indicate percentage increases in clinical cases relative to mean annual estimates under the comparator strategy; corresponding lines with negative gradients represent equivalent percentage decreases and share the same line style. **Figure 6—figure supplement 1. Change in cases vs. cases with triennial pyrethroid-chlorfenapyr distribution.** **Figure 6—figure supplement 2. Cases averted under different ITN distribution strategies.** **Figure 6—figure supplement 3. Change in cases vs *Pf* PR with triennial pyrethroid-PBO distribution.** **Figure 6—figure supplement 4. Change in cases vs pyrethroid resistance.** **Figure 6—figure supplement 5. Change in cases vs duration of use.** **Figure 6—figure supplement 6. Change in cases vs retention time.** **Figure 6—figure supplement 7. Change in cases vs use.** **Figure 6—figure supplement 8. Change in cases vs access.** **Figure 6—figure supplement 9. Change in cases vs use given access.**

The number of clinical cases averted by the different strategies varies substantially, both within (figure 5.C and figure 5 - figure supplement 1-5, panel C) and between countries (figure 6). For example, in Mali switching from a triennial to biennial pyrethroid-PBO mass campaign is estimated to avert fewer than 50 additional cases per 1,000 people in Gao, but over 100 in Sikasso (figure 5.C.vi). The benefit of the different strategies is projected to differ in each setting, though broad trends are observable. Generally, fewer cases are projected to be averted under triennial pyrethroid-chlofenapyr campaigns than with biennial pyrethroid-PBO distribution, with 74.6% of regions having fewer clinical cases predicted when you increase the campaign frequency than when the net class is changed (figure 6 - figure supplement 2.F vs. H and regions with negative y-axis values in figure 6 - figure supplement 1.F). Triennial distribution of pyrethroid-chlorfenapyr ITNs was predicted to avert more cases than biennial distribution of pyrethroid-only ITNs for 94.3% of regions (figure 6 - figure supplement 2.H vs. C and regions with positive y-axis values in figure 6 - figure supplement 1.C). This was more consistent for policy decision-making than the differences between triennial pyrethroid-PBO distribution and biennial pyrethroid-only distribution, with distributing pyrethroid-PBO ITNs averting more cases in 65.6% of regions (figure 6 - figure supplement 2.E vs. C and regions with positive y-axis values in figure 6.C).

It is unlikely that bespoke simulation modeling will be available for all policy decisions, but the models can indicate trends that can support sub-national tailoring of ITNs. The models indicate current clinical incidence is the best predictor of the number of cases averted for the different strategies. For example, for both pyrethroid-PBO (figure 6.F) and pyrethroid-chlofenapyr ITNs (figure 6 - figure supplement 1.I), the projected additional cases averted by switching to biennial distribution is estimated to gradually increase as clinical cases increase up to approximately 300 mean annual clinical cases per 1,000 people, though above this point trends are less obvious. Similar trends are seen in countries with a broad range of transmission intensities, such as Mali (figure 5.C.vi) and Mozambique (figure 5 - figure supplement 4.B.vi), though it is less clear in countries where are no low transmission regions, such as in Burkina Faso (for example, figure 5 - figure supplement 1.C.vi). Similar trends can be observed by using all-age malaria prevalence as an indicator to select regions which will have the greatest benefit of greater investment in ITNs (figure 6 - figure supplement 3). Selecting areas to target using the level of pyrethroid resistance is less reliable (figure 6 - figure supplement 4), in part because there is little variability within countries and differences between countries are confounded by transmission intensity in this analysis. Surprisingly, there was no discernible association between either ITN retention times or the mean duration that nets remain in use on the impact of increasing campaign frequencies (figure 6 - figure supplements 6.F, 5.F). This is because ITNs have a bigger epidemiological impact in areas where they are used for longer, though policy-makers may wish to consider equity in disease burden in their tailoring process which is not considered here. There were, however, clearer relationships that indicated more cases may be averted following a switch from pyrethroid-PBO to pyrethroid-chlofenapyr triennial distribution in regions with greater mean access, use and use given access (figure 6 - figure supplements 8.H, 7.H, 9.H), though these associations are weaker than those seen for clinical cases or prevalence.

The cessation of mass campaigns is projected to lead to notable resurgences in clinical cases in many regions; an additional 150 to 300 clinical cases per 1,000 people are estimated for moderate to high transmission intensity regions (those with more than 150 mean annual clinical cases per 1,000 people under triennial pyrethroid-PBO distribution strategies). It has been suggested that the potential increase in cases caused by halting mass campaigns could be mitigated by concurrently switching to using pyrethroid-chlofenapyr ITNs in continuous distribution channels (figure 6.G vs. D). This modeling exercise indicates such mitigation is unlikely, with only 1 of the 146 regions showing a reduction of cases when switching from combined continuous distribution and triennial mass campaigns with pyrethroid-PBO to continuous distribution alone with pyrethroid-chlorfenapyr. This was for the capital city of Mali, Bamako (figure 5.C.vii), which has moderately low transmission, while Mali was estimated to distribute more ITNs through continuous channels than most other countries considered in this analysis (figure 2 - figure supplement 2).

## Discussion

International guidelines on the use of ITNs have changed over the past two decades, moving from a universal target of 80% overall use in regions at risk of malaria to more nuanced guidance of “optimal” coverage in areas with resource limitations (***World Health Organization, 2024b***). This was an important policy change as it allowed NMPs to consider the use of alternative interventions which may be more cost-effective than pushing for very high population use in areas where this might be unachievable. Funding constraints have also increased the need for consideration of subnational tailoring, with many recommendations being made on the basis of transmission intensity in the ***World Health Organization (2025***) Subnational Tailoring Reference Manual. However, a key uncertainty in assessing the potential impact of different ITN interventions has been how long nets remain in use rather than how long they are retained, and how this varies between regions. Here, to our best knowledge, we present the first estimates of subnational variation in ITN retention and the duration that ITNs remain in use, and also quantify for the first time how ITN use, access and retention vary between subnational regions across multiple African countries. Our work supports the change in guidance to optimal coverage as it highlights ITN interventions have notable differences in impact between settings, and that distributing fewer but more effective ITNs, particularly pyrethroid-chlorphenapyr products, is likely to be more impactful than maximising long-term coverage through increased campaign frequencies with pyrethroid-only ITNs. Our work also broadly supports ***World Health Organization (2025***) recommendations for subnational tailoring, particularly the consideration of deprioritisation of ITN distribution in very low transmission settings. However, our results provide new indications that deprioritisation of areas with higher ITN use given access may lead to greater resurgences in cases, highlighting that subnational tailoring decisions could be optimised further by considering additional factors to transmission intensity alone. The modelling results indicate that the overall 80% use and access threshold was seldom consistently achieved across many countries in sub-Saharan Africa, and though this threshold appears relatively arbitrary, it is unclear how the new guideline of ‘optimal’ ITN use should be assessed. Mean use of ITNs under triennial mass campaigns with supplementary continuous distribution was estimated to be, on average, 45.4% (95% CrI: 45.3, 45.5) over all regions included in our analysis. This highlights that status quo approaches are typically yielding mean levels of ITN use that are roughly half of minimum universal coverage targets. Increasing mass campaign frequency to every two years is estimated to increase mean use by 8.50% (95% CrI: 8.47, 8.54), and is therefore unlikely to close this gap. Larger increases in mean use could plausibly be predicted under alternative ITN loss functions, such as the sigmoidal loss function used by ***Bhatt et al. (2015)*** and ***Bertozzi-Villa et al. (2021)***. However, inspection of ITN age distributions from DHS surveys provided limited evidence against the exponential decay function assumed here (appendix 2.2). Nevertheless, our estimates of mean use are consistent with previous modelling across a wider range of malaria-endemic countries by ***Bertozzi-Villa et al. (2021)***, where use was estimated to have broadly plateaued between 40% and 50% from approximately 2015 onwards, despite the use of a different loss function. Unless more nets are distributed or people keep their nets for significantly longer, universal coverage has potentially always been an unobtainable goal between mass campaigns.

The benefit of ITNs depends on how long people keep and use them. We therefore aimed to identify systematic differences in subnational use of and access to ITNs. Our central estimates indicated that 86.3% of subnational regions had a mean duration of access less than three years. Although the subnational regions included in this study are from a subset of countries in sub-Saharan Africa, our findings are broadly consistent with previous model-derived estimates of national median retention times, which have been estimated to be less than three years for 87% (***Bertozzi-Villa et al., 2021***) to 100% (***Bhatt et al., 2015***) of sub-Saharan African countries. Since we assume the duration of ITN use and access is exponentially distributed, our models incorporate a greater probability that a net will be lost, or cease to be used, in the immediate period after receipt. Therefore, we would argue our mean estimates are more comparable to the median estimates by ***Bertozzi-Villa et al***. (***2021***) and ***Bhatt et al***. (***2015***), which both assume a sigmoidal decay function. Although some field studies have reported ITN retention times greater than three years (***Obi et al., 2020; Diouf et al., 2022***), results differ by location, with the majority of field studies (for exmaple, ***Gnanguenon et al., 2014; Solomon et al., 2018; Lorenz et al., 2020***) estimating that ITN retention fails to exceed three years in most settings. Even in isolation, our projections of additional cases averted under biennial over triennial campaigns raise further questions on whether the status quo of three-year intervals is sufficient to maintain adequate or “optimal” coverage; especially given its original basis stems from the duration of pyrethroid-class insecticidal activity estimated two decades ago (***World Health Organization, 2005, 2013, 2023***).

While previous work on net retention has largely focused on access, here we have also estimated the mean duration for which ITNs remain in use, which has greater epidemiological importance. Median estimates of the mean duration of use were less than three years in all but one of the subnational regions investigated. The work highlights that there is a faster rate of loss of use than access. Although the causes of this are unclear, it could be driven by poor quality ITNs, with people with older nets potentially not using them if degraded ITNs are perceived to provide lower protection. Indeed, reduced ITN integrity has been associated with reduced probability of use (***Hiruy et al., 2021***). ITN age is thought to be a better predictor of infection risk than durability alone (***Andronescu et al., 2019***). Therefore, even if duration of use is lower in some regions due to reduced ITN integrity, SBC campaigns either to promote use or to help with net care may still be beneficial.

We characterised historical trends in overall ITN use and access over time in different subnational regions. These estimated a gradual increase in both overall use and access over time, with waxing and waning dynamics driven by timings of mass campaigns; similar national patterns of declining use following mass campaigns have also been estimated in models by (***Bhatt et al., 2015***) and recorded in longitudinal field surveys (***Hamre et al., 2020***) and cluster randomised control trials (***Protopopoff et al., 2018***). Furthermore, surveillance data in multiple settings have also highlighted notable increases in malaria cases in the second year following mass campaigns (***Zhou et al., 2016; Wotodjo et al., 2017; Girond et al., 2018; Topazian et al., 2021***), providing further, albeit indirect, indications that overall ITN use declines notably in real-world settings by the second year following mass campaigns.

Although neither overall ITN use nor access in any subnational regions were projected to be maintained above the universal coverage threshold of 80%, mean use given access under present triennial strategies was estimated to be greater than 80% for almost half of subnational regions. This highlights that two decades after their widespread introduction, ITNs remain a valued commodity and key weapon in the expanding arsenal of malaria interventions when individuals have access to them. Moreover, if DHS surveys were primarily conducted in dry seasons in some locations (which is often the case), when ITN use given access is typically lower, our estimates in turn may be underestimated in some settings, especially in Senegal and regions of Mali, which are thought to have more seasonal fluctuations in use (***Koenker et al., 2019b***). However, to achieve consistent attainment of high population use targets, alternative distribution strategies, such as enhanced continuous distribution channels may be needed (***Koenker et al., 2023b***).

By identifying subnational regions with systematic low use given access and short durations of access, we highlight geographical areas where ITNs are likely to perform less effectively, irrespective of the local transmission intensity. This may aid NMPs in tailoring subnational strategies aside from those related to ITN distribution. For example, in regions where local knowledge indicates low use given access is driven by social and/or behavioural factors, SBC communication may be beneficial. Local understanding of the barriers to ITN use will be instrumental in assessing the cost-effectiveness of SBC strategies. While SBC communications, such as ‘hang-up’ campaigns have led to reported 250% relative increases in use in Zambia (***Macintyre et al., 2012***) and absolute increases between 11% and 17% in Togo (***Desrochers et al., 2014***) and Nigeria (***Kilian et al., 2016***), in other settings, such as in Uganda (***Kilian et al., 2015***), significant increases in use was not observed. We have not investigated the underlying factors that may have led to poor utilisation of ITNs in some subnational regions. We assume that use, access, and use given access immediately following a campaign, as well as their decay rates, remain unchanged when shifting from triennial to biennial strategies, and further work is needed to understand how use may change for different distribution strategies. Importantly, poor utilisation may arise due to ITN degradation (***Mboma et al., 2021***). An assessment of ITN durability studies across seven sub-Saharan African countries by ***Briet et al***. (***2020***) found greater variability in durability between locations than between different net brands. Damaged ITNs may contribute not only to reduced utilisation, but also in reduced efficacy, particularly in areas with pyrethroid-resistant mosquitoes (***Wheldrake et al., 2024***).

Subnational tailoring of ITN interventions is increasingly recommended, with some ***World Health Organization (2024b***,a, 2025) guidelines currently advising lower transmission settings to be considered first for deprioritisation from mass campaigns. Optimal subnational tailoring may not solely rely on deprioritisation, but may also require repriotisation of resources to other regions. Increasing the frequency of mass campaigns or procuring more effective, but likely more expensive, ITNs will inevitably increase the costs of conducting mass campaigns if the same level of coverage is achieved. If budget constraints necessitate the deprioritisation of campaigns, our results highlight that this should be avoided, if possible, in regions with moderate to high transmission intensity, particularly those with mean annual incidence exceeding 100–150 clinical cases per 1,000 people. However, current incidence should be considered alongside the factors that gave rise to that transmission intensity, with caution exercised when deprioritising mass campaigns in areas where historically higher transmission may currently be suppressed by high ITN access, high use given access, or other interventions. Shortening campaign intervals from three to two years in regions with current moderateor high-transmission is projected to avert more cases than the additional cases that may arise from ceasing campaigns in some lower-transmission settings. Additionally, although pyrethroid–chlorfenapyr ITNs are more costly, the additional cases projected to be averted by them relative to pyrethroid-only and pyrethroid–PBO ITNs are substantial. In certain national contexts it may be more cost-effective for biennial pyrethroid-chlorfenapyr campaigns to be conducted in fewer subnational regions even under reduced budgets. However, more thorough economic analyses will be needed to understand this fully. Moreover, as ITNs remain one of the most cost-effective malaria control interventions, improving the impact of them could still be more costeffective than the introduction of new untested interventions (***Topazian et al., 2023; Schmit et al., 2024***).

In the context of finite procurement budgets, our framework identified subnational regions where increasing the frequency of mass campaigns or switching to more effective ITNs may have the greatest impact. Although we only included six countries in our analysis, they not only cover a wide geographical range, but also a broad spectrum of *P. falciparum* transmission intensities and estimated levels of pyrethroid resistance (***Hancock et al., 2020***). For all countries considered, transmission intensity was positively correlated with increased benefit of both switching to two-year mass campaign cycles or more efficacious ITN classes. The framework can be extended to other countries as DHS data becomes more available. In the absence of this, our results indicate areas with greater transmission intensity are likely to see the greatest impact from either increasing mass campaign frequencies or switching to pyrethroid-PBO or pyrethroid-chlorfenapyr ITNs, supporting WHO guidelines ***World Health Organization (2024a***). Ultimately, decisions need to be made by NMPs whether they want to target the most cases, or whether decisions should be made with respect to equity of resources (does everyone have similar access to a net) or equity of impact (does everyone have a similar chance of getting malaria), both within and between regions.

As the toolbox for malaria control continues to expand, there is increasing need for ongoing monitoring and evaluation to facilitate evidence-based decision making and optimal deployment of interventions (***Wilson et al., 2020***). Longitudinal standardised surveys, such as the DHS, have the potential to be instrumental in assessing the effectiveness of interventions in real world settings, and have the potential to be used to refine distribution strategies to maximise impact. Such punctuated surveys can aid in monitoring the use of different public health products, such as spatial repellents or traps, and could independently assess the success of drug and vaccine campaigns. However, the timing of DHS surveys every few years creates challenges for assessing the effectiveness of both ITNs and other commodities. For example, if two DHS surveys were conducted at different times in relation to the last mass campaign, underlying trends in ITN use or access may fail to be captured. Similarly, if data are collected irregularly in relation to other future punctuated interventions, such as intermittent rapid diagnostic testing following mass vaccination campaigns, the methodology presented here provides a framework where potential biases can be controlled for, provided estimates of the duration of vaccine-induced immunity and the timings of vaccination campaigns are known. With appropriate adaptation to our model, this framework can facilitate more reliable inferences on the effectiveness of interventions beyond malaria than would be possible from survey data alone.

In conclusion, the work indicates that universal coverage targets of 80% are unlikely to be consistently met due to waning overall ITN use in the intervening years between triennial mass campaigns. Improved coverage can be achieved through more frequent biennial distributions, though this is unlikely to be feasible at scale given the current funding landscape. Indeed, when resources are constrained, deprioritisation of ITN mass campaigns in certain settings is being increasingly considered through subnational tailoring of malaria control interventions. Our work highlights that the relationship between transmission intensity (whether measured in terms of prevalence or clinical cases) and intervention impact is non-linear, and notable resurgences in cases may follow when campaigns are deprioritised in all but very low transmission settings. This broadly supports WHO subnational tailoring guidance, which suggests consideration of deprioritising distribution of ITNs in regions with *Pf*PR_2−10_ < 1% (***World Health Organization, 2025***). However, while the ***World Health Organization (2025***) Subnational Tailoring Reference Manual proposes that the withdrawal of ITNs in favour of indoor residual spraying should be considered in areas with low ITN use, here we estimate that ITN use alone appears to be a notably poorer predictor of the impact of ceasing mass campaigns than use given access. Our findings suggest that regions with higher use given access may experience disproportionately greater resurgences in cases following deprioritisation. This implies that regions with low use given access may warrant consideration for cessation of ITN distribution, rather than decisions being based solely on low overall ITN use irrespective of whether communities have sufficient ITN access. However, subnational differences in ITN use, access and retention are key knowledge gaps in many settings, and when estimated from infrequent surveys they are highly sensitive to bias arising from the timing of surveys relative to when campaigns were conducted. To our knowledge, this study is the first to estimate subnational variation in ITN retention and the first to estimate the duration that ITNs remain in use, which is of greater epidemiological relevance than retention time. It also provides a novel framework to correct for biases in estimates of ITN use and access arising from when campaigns were conducted. Although campaigns have historically aided increasing ITN use and access over time, we estimate the mean duration of ITN use is consistently shorter than mean retention times in all regions. This raises questions about whether punctuated distribution of ITNs through campaigns is the optimal mechanism for maximising their effectiveness and cost-effectiveness. Maximising the cost-effectiveness of interventions has become increasingly pertinent in the current funding context, and consideration of alternative distribution strategies, such as increased distribution through continuous distribution channels, including school- or community-based distribution, may be warranted. Frameworks such as the one presented here, which take into account the potential for impact from different net types and the high variability of ITN duration and use, could support NMP decision-making on how best to maximise impact from available funds. Whilst such frameworks may be a useful tool, local knowledge of factors impacting ITN access and use as well as operational decision making will be paramount for NMP-led tailoring of subnational strategies.

## Methods and Materials

### Inclusion criteria and data sources

The majority of data were sourced from Demographic and Health Surveys (DHS) and Malaria Indicator Surveys (MIS) (***ICF, 2025***). AIDS Indicator Surveys (AIDS) were also included where additional malaria questions were included (***ICF, 2025***). Subsequent references to DHS surveys refer to all surveys included. Data on the number of ITNs distributed nationally were also sourced from (***The Alliance for Malaria Prevention, 2024***).

Here we assume each ITN is used every night for a continuous period after receipt (duration of use), but may remain accessible for a different length of time following receipt (duration of access/retention time). It is important to distinguish between people who have access to an ITN but don’t sleep beneath it and those who don’t have access to one. We generate subnational estimates of both the population mean duration of use and retention time and assume these are constant over time. Single estimates of both these quantities are estimated separately for rural and urban areas of each region to allow the identification of systematic differences between locations that could allow the geographical targeting of resources.

Retrospective analyses were conducted from 2008, to align with the introduction of mass distribution campaigns, until 2024. Analyses were conducted at an administrative-one level stratified by urban and rural settings, as recorded in survey responses. To account for inconsistencies in administrative-one naming conventions, such as alternative French and English spelling, best efforts were made to unify recorded administrative-one entries with the naming conventions recorded by GADM v4.0 (***Global Administrative Areas, 2021***). Note that while administrative-one is referred to as a region in Senegal, for example, terminology differs between countries; here, the units of analysis are consistently referred to as (subnational) regions. The initial inclusion criteria were restricted to countries where at least three DHS/MIS surveys had been conducted between 2010 and 2022. This was to ensure there were sufficient data for all countries after mass campaigns had been established. Ten countries initially met this criteria: Burkina Faso, Ghana, Liberia, Malawi, Mali, Mozambique, Nigeria, Senegal, Tanzania and Uganda.

All mass campaigns were assumed to be conducted nationally, although their timings were not restricted to occur in synchrony across subnational regions. Instead, the timing of each mass campaign was allowed to vary subnationally within time intervals that were defined at a national level. Due to this assumption, Liberia was excluded from our analysis due to rolling campaigns being conducted between 2008 and 2012 (***Koenker et al., 2019a***). We also excluded Nigeria where mass campaigns follow different distribution schedules between states (***Koenker et al., 2019a***). Tanzania was also excluded as a mixture of annual school-based distributions and five-yearly mass campaigns are conducted within different subnational regions (***Koenker et al., 2023a***). We finally excluded Uganda since there have been notable changes to its administrative-one level boundaries over the assessed period; this has resulted in significant discrepancies between the administrative-one level regions recorded in DHS surveys and the naming conventions used in GADM v4.0. Although Ghana was included in all analyses, the sub-division of the regions of Brong-Ahafo, Northern and Volta in 2019 was not accounted for in the retrospective analyses of historical use and access; the new regions were however accounted for in the transmission dynamics models for future projections. For Malawi, we conducted retrospective analyses of use and access at an administrative-one level (termed regions in the country); however, as data we sourced for other covariates, such as rainfall patterns and non-ITN interventions, were stratified at an administrative-two level (districts) for Malawi (***Winskill, 2024b***), we excluded the country from the transmission dynamics analyses. These other covariates were stratified at the administrative-one level for all other countries (***Winskill, 2024b***).

All subsequent analyses described are conducted independently for use and access and were conducted in R (v4.3.2, ***R Core Team, 2023***). The code for models used in this study can be found at https://github.com/andrewcglover/itn_campaigns. Unless otherwise stated, median central estimates are quoted in the main text.

### Historical use, access and retention times

Due to the sparsity and irregularity of DHS and MIS surveys, we were unable to investigate seasonal fluctuations in either access or use; we therefore assume that nets provide access or are used continuously over some period of time. Although seasonal differences are not identifiable under our model, overdispersion in individual-level use and access may partially absorb unexplained variation between observations, including variation arising from unmodelled seasonal differences (equation 7). However, this does not allow us to quantify seasonal variation in ITN use or access. If ITN use were systematically higher during high-transmission rainy seasons, our assumption of continuous use may underestimate the protective impact of ITNs during these periods. We do not assume these periods are equal, and so differentiate between the duration of time that a net is slept under (duration of use) and the period for which it provides access before being discarded or repurposed (retention time). Using a Bayesian hierarchical model fitted to pooled ITN age distribution data across DHS surveys, we separately generated initial subnational estimates of the mean duration of use and mean retention times of ITNs from DHS survey data using the age distributions of ITNs in surveyed households. When inferring mean retention times under our hierarchical model, we implicitly assume ITNs are only distributed to individuals without them; this is a necessary methodological step, but is relaxed later in our analysis. Although the methodology is applied analogously in the context of use, the remainder of this section focuses on access.

Under the assumption that nets are lost at a constant rate, the long-term age distribution of nets providing access follows an exponential distribution, allowing retention times to be inferred from observed net ages. The hierarchical structure pools information across regional, national, and continental spatial scales to reduce bias arising from survey timing relative to mass campaigns, particularly in regions with sparse survey data. Region-specific mean retention times were modelled with country-level random effects, and country-level means were in turn modelled with a continent-level random effect. Right-censoring of reported net ages at 36 months and survey sampling weights were explicitly accounted for in the likelihood when fitting the model in Stan via the R interface RStan (v2.32.6, ***Stan Development Team (2024***)). Full specification of the hierarchical model and prior distributions are provided in appendix 2.

We then sought to estimate the timings of mass campaigns in different regions. We use annual estimates of the number of ITNs delivered nationally from the AMP Net Mapping Project (***The Alliance for Malaria Prevention, 2024***) to estimate time intervals within which each mass campaign was likely to occur for each country. We do however allow for the timings of campaigns to vary subnationally within these intervals. For each region, the timing of each mass campaign is estimated probabilistically by constructing distributions over plausible months using DHS data on the ages of ITNs recorded in surveys. As time elapses since a mass campaign, ITNs are less likely to be recorded in DHS surveys. Our initial ITN retention estimates help correct for this bias (see appendix 3.1 for further details). We can then probabilistically infer the timing of the previous campaign (*ϕ*_*j*_) relative to some time *t*_*j*_ in a given region (see appendix 3.2 for further details), noting that in contrast to appendix 3.2, we do not explicitly indicate subscript *i* notation to indicate the region of interest for notational ease. In turn, this allows us to infer the time since the previous campaign (*m*_*j*_) relative to all calendar times considered in our model:

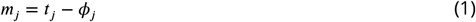

We then utilise the following discrete-time model of the proportion of the population with access (*p*_*j*_) for a given region at some time *t*_*j*_ in the observed survey period:

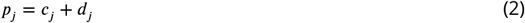

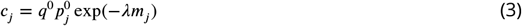

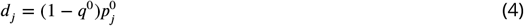

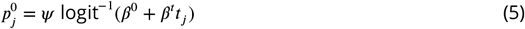

where superscripts 0 and *t* are used for notational purposes and do not indicate exponentiation. An in-depth explanation of the model can be found in appendix 4.1, but in brief, we firstly assume access can be attributable to ITNs received from campaign (*c*_*j*_) or continuous distribution (*d*_*j*_) channels (equation (2)). Secondly, we use *q*^0^ to denote the proportion of the population with access to an ITN from a campaign relative to access to any ITN immediately post-campaign, which is assumed to be the same following all campaigns for a given region (equation (3)); the relative proportion with access to an ITN from continuous distribution channels at these times is therefore given by 1 − *q*^0^ (equation (4)). Thirdly, we assume logistic-type growth in access achieved immediately following each subsequent campaign, denoted by 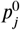, this growth is characterised by the parameters 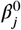 and 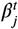 (inverse-logit function in equation (5)), and is bounded by an upper limit, *ψ*. We therefore also implicitly assume logistic-type growth in access to ITNs from continuous channels over time (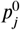 in equation (4)) and ITNs distributed through campaigns over time (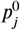 in equation (3)). Finally, we assume access to campaign nets decays following mass campaigns (exponential term in equation (3)); the rate of this decay is governed by the decay parameter, *λ*, which is related to the mean retention time (Λ^−1^) such that:

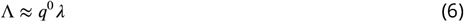

where retention times are assumed to be the same for all nets irrespective of their distribution channel. In contrast to the earlier hierarchical model, the discrete-time model and subsequent intervention impact projections assumes random allocation of ITNs in deriving the updated estimates of mean retention time (equation (6)). Under this assumption, at the point of distribution, an individual already with access to an ITN has the same probability of receiving a new ITN as an individual currenlty without access. Further details on how retention times are inferred from the approximation in equation (6) can be found in appendix 4.4.

We do however use our regional posterior estimates of mean ITN retention times from our previous hierarchical model (which assumed ITNs are only distributed to those without access) as informative priors for mean retention times under random allocation to aid parameter identifiability. Either non-informative or regularising priors are used for other region-specific parameters (see appendix 4.2 for further details).

When fitting our model in Stan with the R interface RStan (v2.32.6, ***Stan Development Team (2024***)), we accounted for the potential of individual-level access (and indeed use, in that context) to vary subregionally; for example, between different wards or districts. Indeed, examination of the raw data (for example, figure 2 figure supplement 1.A) indicates substantial variability in the recorded proportions of use and access in adjacent months. This may reflect substantial subregional variability, and there are likely other covariates that our model does not account for that will create additional variability in the probability of access (and use) within an administrative-one unit at any given point in time. Our discrete-time model thus far describes the proportion of the population with access (*p*_*j*_ in equation (2)). However, we also consider the probability of individuals having access at each time *t*_*j*_, which is denoted by 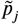. We do not assume the probability of access is the same for all individuals in a region at a given point in time. Instead, we assume the probability any given individual has access to an ITN at time *t*_*j*_ can be described by a Beta distribution:

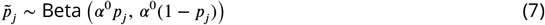

Under this assumption, the mean probability of access across all individuals at a given time is equal to the overall proportion of the population with access. Meanwhile, for a given proportion of the population with access, the overdispersion parameter, denoted by *α*^0^, is approximately inversely proportional to the variance in the probability of individual-level access. This overdispersion parameter, which is fitted for each region and assumed to remain constant for each over time, therefore controls the estimated level of subregional heterogeneity in access. DHS data on the total number of surveyed individuals by month, in addition to the number with access to ITNs from different sources, are accounted for in our likelihood specification for region-specific access over time and the relative contribution towards this from campaign and continuous distribution channels. Further details can be found in appendix 4.3.

With the exception of panels B and C respectively in figures 2 and 5 (and associated supplementary figures), figures 1 - 5 are generated using predictive posterior estimates from our discrete time model. Current mean estimates under triennial (or biennial) campaigns (figure 3) are inferred from equations (2) - (5) over a three- (or two-) year period had a hypothetical campaign been conducted in every region at the start of January-2025, using posterior parameter estimates for December-2024 (the end of the period of included surveys). Estimates of the proportion of the population with different probabilities of access (and use) are then inferred from this using equation (7) (figure 4). This provides a measure of equity in use and access of nets within a region which will have important policy implications for distribution strategies.

### Simulating future distribution strategies

The future scenarios considered were no future ITN distribution, continuous distribution in isolation, in addition to biennial and triennial campaigns with supplementary continuous distribution. Distribution of pyrethroid-PBO ITNs is assumed in all regions prior to December-2024. Distribution of different ITN classes (pyrethroid-only, pyrethroid-PBO or pyrethroid-chlorfenapyr) is simulated from January-2025 onwards for each distribution strategy. Biennial and triennial distribution strategies are respectively simulated to commence either 2or 3-years following the median estimate of the last estimated campaign prior to 2025. For strategies with campaigns, clinical cases are recorded over a 6-year interval commencing on the first simulated campaign after December-2024. Clinical case estimates for continuous-only distribution and no future distribution are recorded over a 6-year period commencing on the mid-point between the start of the recorded 6-year biennial and triennial scenarios, should these differ for a region. For each subnational region, 100 stochastic realisations were simulated for a population of 100,000 individuals, and cases averted were calculated relative to the baseline scenario with no future ITN distribution. For each realisation, parameters are drawn from our joint posterior distribution of fitted parameters from our discrete-time model of use, however ongoing logistic growth in use of ITNs from continuous channels and immediately following campaigns is not assumed beyond December-2024. Future campaigns, whether conducted every two or three years, are therefore assumed to achieve a consistent initial level of use. Since epidemiological dynamics are only dependent on whether ITNs are ultimately used, our simulations are agnostic of our access estimates. For each realisation and distribution event, parameter draws are also made from previously generated posterior distributions of the probability of repellency and mortality at different pyrethroid-resistance levels; these were fitted to data from systematic reviews of experimental hut trials across Africa (***Sherrard-Smith et al., 2022; Churcher et al., 2024***). Region-specific uncertainty in ITN efficacy, use, retention, and the relative contributions of continuous and campaign channels is therefore propagated through to our estimates of cases averted.

Regional differences in transmission intensities are accounted by calibration of historical baseline estimates of the entomological inoculation rate (EIR) prior to the introduction of any interventions. Baseline EIR estimates were calibrated by fitting our modelled annual mean *Plasmodium falciparum* prevalence in children between the ages of 2 and 10 (*Pf*PR_2−10_) to those estimated by the ***Malaria Atlas Project (2024***) (figure 2.B). Forecasted increases in pyrethroid resistance were incorporated, while coverage of other interventions (including IRS, SMC, drug treatment and vaccination) was assumed to be maintained at 2024 levels in each region; these were estimated using the *site* (v0.2.2) R package (***Winskill, 2024b***) package

Future projections were generated using *malariasimulation* (v1.6.0) (***Charles et al., 2024***), an individual-based malaria transmission model that incorporates typical seasonal patterns in transmission. The transmission dynamics model, which incorporates typical seasonal patterns in each region, is broadly able to reproduce the observed changes in *Pf*PR_6−59mo_, as estimated by DHS surveys, providing some confidence in the resulting epidemiological predictions. However, while the model captures typical seasonal dynamics, it does not account for inter-annual variation or specific weather events. These factors may have contributed to larger deviations from DHS *Pf*PR_6−59mo_ point estimates at certain times; estimates that themselves often have wide 95% credible intervals (figure 2.B). Full details of model calibration, ITN efficacy parameters, and simulation procedures are provided in appendix 5.

## Data Availability

All data used in this study are publicly available. ITN access and use data were obtained from Demographic and Health Surveys (DHS), Malaria Indicator Surveys (MIS), and AIDS Indicator Surveys (AIS), accessible via https://dhsprogram.com upon registration. National-level ITN distribution data were sourced from the Alliance for Malaria Prevention (AMP) Net Mapping Project: https://allianceformalariaprevention.com. All other covariate data were accessed using the 'site' R package (Winskill et al., 2024), which is publicly available at https://mrc-ide.github.io/site/. Country-specific datasets used by the package are accessible upon request to the package maintainers, with instructions provided at the same site. All analyses were conducted in R (v4.3.2). The code for all models and analyses used in this study is publicly available at: https://github.com/andrewcglover/itn_campaigns

https://mrc-ide.github.io/site/

https://allianceformalariaprevention.com/itn-dashboards/net-mapping-project/

https://dhsprogram.com/data

## Acknowledgments

The work was funded by the Global Fund under the Net Transition Initiative. ACG and TSC acknowledge funding from the Medical Research Council (MRC) Centre for Global Infectious Disease Analysis (reference number MR/R015600/1), jointly funded by the MRC and the UK Foreign, Commonwealth and Development Office (FCDO), under the MRC/FCDO Concordat agreement, and is also part of the EDCTP2 program supported by the European Union. Authors would like to thank the surveyed communities and those involved in the collection of the Demographic Health System data.

## Appendix 1

### 1 Methodological summary

The methods used here can be divided into 4 main steps. Firstly, the mean durations of use and access (retention time) are initially estimated from a hierarchical model fitted to the pooled age distributions of ITNs that were used or provided access (Appendix 1 - figure 1.A, with further details in appendix 2). ITN use is inferred from DHS data (***ICF, 2025***) on whether individuals slept under an ITN the previous night. While all individuals who used an ITN are assumed to have access, when fewer than two individuals used an ITN, that ITN is assumed to be able to provide access to up to two individuals in total within a household. Under this assumption, when ITNs in a household can provide access to more individuals than the number of users, access is assigned at random to non-users within each household under our framework. DHS data were obtained throughout with the *RDHS* R package (v0.8.4) (***Watson et al., 2019***).

Secondly, the timings of campaigns are estimated at a monthly resolution for each subnational region (Appendix 1 - figure 1.B, with further details in appendix 3). This step utilities annual ITN deliveries by country from the ***The Alliance for Malaria Prevention (2024***) Net Mapping Project and the monthly empirical distribution of delivery dates from DHS surveys (***ICF, 2025***). Initial estimates of retention times are used to correct for sampling bias due to older ITNs being less likely to be observed in DHS surveys.

Thirdly, inferred use and access of ITNs from different channels from DHS surveys are used to fit discrete-time models of historical use and access for each subnational region (Appendix 1 - figure 1.C, with further details in appendix 4). This utilises previously generated estimates of when campaigns occurred in each region. ITN retention estimates are refitted during this process, with previously generated estimates used as informative priors.

Finally, clinical case estimates are generated for different ITN distribution strategies using our estimates of the mean duration of use, the level of use achieved immediately following a campaign, and the relative contribution to use from campaign and continuous distribution channels for each subnational region (Appendix 1 - figure 1.D, with further details in appendix 5). Simulations are parameterised for different ITN types using estimates of the probability of repellency and mortality ***Sherrard-Smith et al***. (***2022***); ***Churcher et al***. (***2024***) for region-specific estimates of pyrethroid resistance from the *site* R package (v0.2.2) (***Win-skill, 2024b***). Simulations also account for other region-specific characteristics, including, demography, seasonality and non-ITN interventions using the the *site* package. Historical estimates of ITN use and region-specific characteristics from the *site* package, are used alongside mean annual *Pf* PR_2−10_ estimates from the ***Malaria Atlas Project (2024***) to calibrate historical entomological inoculation rates (EIR) prior to the introduction of interventions using the *cali* R package (v1.0.8) (***Winskill, 2024a***). Clinical case projections are then estimated using the individual-based transmission dynamics model *malariasimulation* (v1.6.0) (***Charles et al., 2024***) for different future ITN distribution strategies.

**Appendix 1—figure 1.**
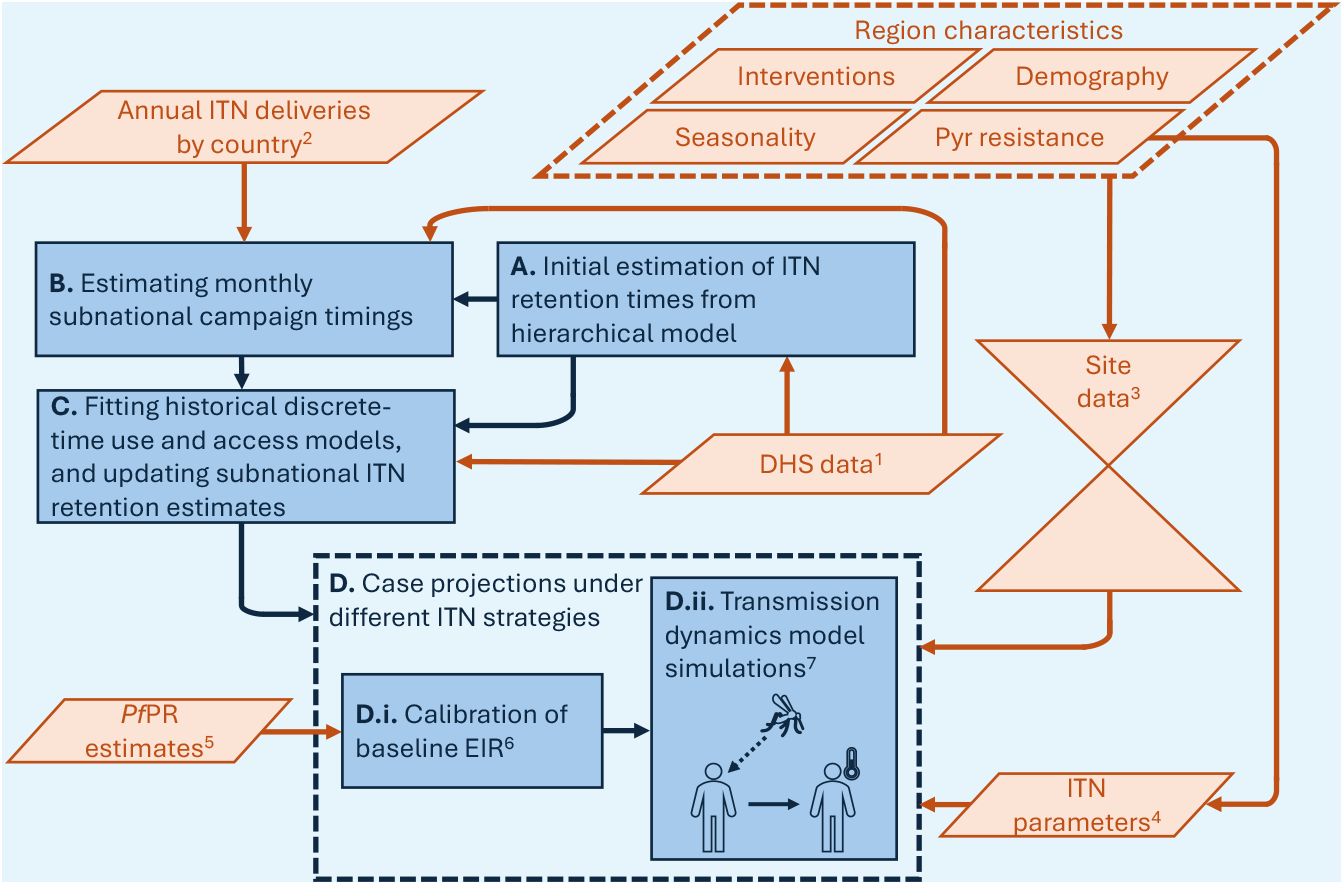
Flowchart of methods. Data inputs and methodological steps are respectively summarised in orange and blue. ^1^DHS data (***ICF, 2025***) were sourced using the *RDHS* R package (v0.8.4) (***Watson et al., 2019***). ^2^Annual estimates of the number of ITNs delivered by country were obtained from the ***The Alliance for Malaria Prevention (2024***) Net Mapping Project. ^3^Region-specific characteristics were sourced from the *site* R package (v0.2.2) (***Winskill, 2024b***). ^4^Parameter values that describe the probability of repellency and mortality as different ITN types age were sourced from ***Sherrard-Smith et al***. (***2022***); ***Churcher et al***. (***2024***) conditioned on annual pyrethroid (Pyr) resistance estimates for each subnational region from *site*. ^5^Mean annual *Pf*PR_2−10_ were sourced from the ***Malaria Atlas Project (2024***); ^6^these were used to calibrate baseline entomological innoculation rates (EIR) with the *cali* (v1.0.8) R package (***Winskill, 2024a***) given historical ITN and non-ITN interventions, with coverage estimates of the latter sourced from *site*. ^7^Case projections were simulated for different ITN distribution strategies (D) using the *malariasimulation* R package (v1.6.0) (***Charles et al., 2024***) following characterisation of subnational differences in ITN use, access and retention (A-C).

Additionally, in relation to uncertainty estimates, credible intervals are shown for all sub-national quantities that are directly estimated in our models. National and continental values are reported as population-weighted summaries of the median subnational estimates generated from the discrete-time models (appendix 4) and therefore do not correspond to explicitly estimated model parameters, so credible intervals are not shown for these aggregated estimates.

#### 1.1. Tables of parameters

Here, core notation used in the main text and throughout all appendices is shown in appendix 1 - table 1. Parameters used in appendices 2-5 beyond those defined in appendix 1 - table 1 are shown respectively in appendix 1 1 - tables 2-5. Superscript and subscript notation in the tables may be simplified or omitted elsewhere in the text for notational ease; for example, the age (*α*_*il*_) of a given ITN *l* in region *i* may be denoted elsewhere as *α*_*l*_. For brevity, notation for counting variables (e.g. total numbers of regions or surveys), functions and interval bounds that are defined locally are not always listed in the notation tables. Finally, although some parameters in the tables are used in both the context of ITN use and access, as indicated elsewhere in the text, this does not necessarily mean they will be equal for both contexts.

**Appendix 1—table 1.** Core notation used throughout the main text and all appendices.

| Symbol | Definition |
| --- | --- |
| $i$ | Subnational region index. |
| $j$ | Time point index. |
| $k$ | Mass campaign index. |
| $l$ | ITN or individual index, depending on context. |
| $t$ | Continuous time variable. |
| $t_j$ | Discrete time point corresponding to month $j$. |
| $\zeta(i)$ | Country to which region $i$ belongs. |
| $a_{ij}$ | Proportion of the population with ITN access in region $i$ at time $t_j$. |
| $u_{ij}$ | Proportion of the population with ITN use in region $i$ at time $t_j$. |
| $\tau_{ik}$ | Timing of the $k^{\text{th}}$ mass campaign in region $i$. |
| $\phi_{ij}$ | Timing of the most recent mass campaign in region $i$ before time $t_j$. |
| $m_{ij}$ | Number of months since the most recent mass campaign in region $i$ at time $t_j$. |
| $p_{ij}$ | Proportion of the population with ITN use or access in region $i$ at time $t_j$. |
| $c_{ij}$ | Component of $p_{ij}$ attributable to campaign-distributed ITNs. |
| $d_{ij}$ | Component of $p_{ij}$ attributable to continuously-distributed ITNs. |
| $p_{ij}^0$ | Immediate post-campaign level of ITN use or access in region $i$ at time $t_j$. |
| $q_{ij}$ | Proportion of ITN use or access attributable to campaign-distributed ITNs in region $i$ at time $t_j$. |
| $q_i^0$ | Proportion of immediate post-campaign ITN use or access attributable to campaign-distributed ITNs in region $i$. |
| $\psi_i$ | Upper asymptote for $p_{ij}^0$ in region $i$. |
| $\beta_i^0$ | Region-specific intercept parameter governing the logistic-type trajectory in immediate post-campaign use or access. |
| $\beta_i^t$ | Region-specific slope parameter governing the logistic-type trajectory in immediate post-campaign use or access over calendar time. |
| $\lambda_i$ | Region-specific exponential decay rate for the campaign-derived component of ITN use or access. |
| $\gamma_i^{-1}$ | Mean duration of ITN use or access (retention time) in region $i$ under the assumption of non-random allocation of ITNs. |
| $\Lambda_i^{-1}$ | Mean duration of ITN use or access (retention time) in region $i$ under the assumption of random allocation of ITNs. |
| $\tilde{p}_{ij}$ | Individual-level probability of ITN use or access in region $i$ at time $t_j$. |
| $\alpha_i^0$ | Region-specific overdispersion parameter governing heterogeneity in the individual-level probability $\tilde{p}_{ij}$. |
| $\alpha_i$ | Age distribution of ITNs providing use or access in region $i$. |
| $\alpha_{il}$ | Age of ITN $l$ in region $i$ at the point of observation. |
| $w_{il}$ | DHS household survey weight for ITN $l$ in region $i$, rescaled to remove the DHS factor of $10^6$. |

**Appendix 1—table 2.**
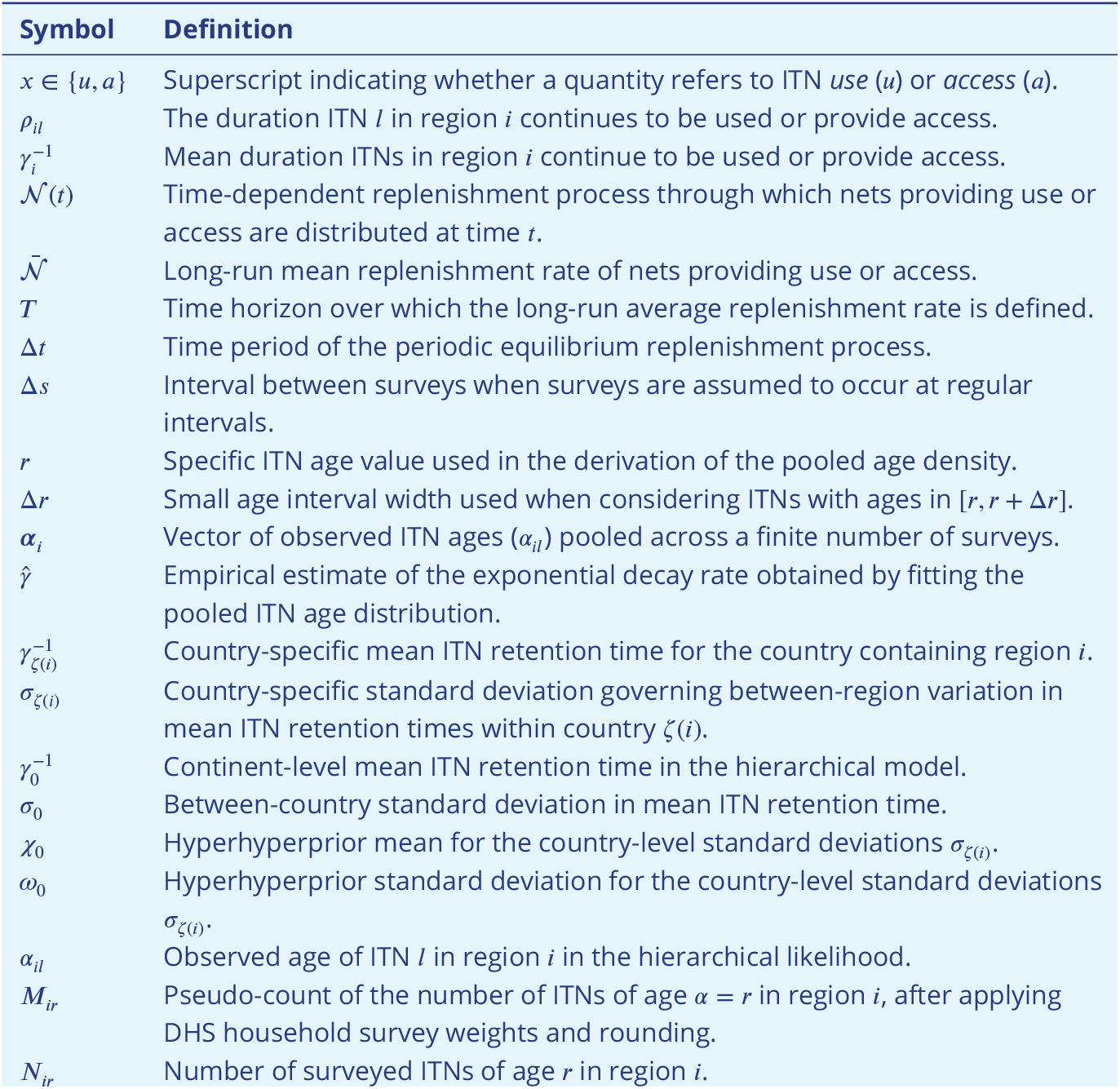
Additional notation used in appendix 2 beyond the core notation defined in appendix 1 - table 1.

**Appendix 1—table 3.**
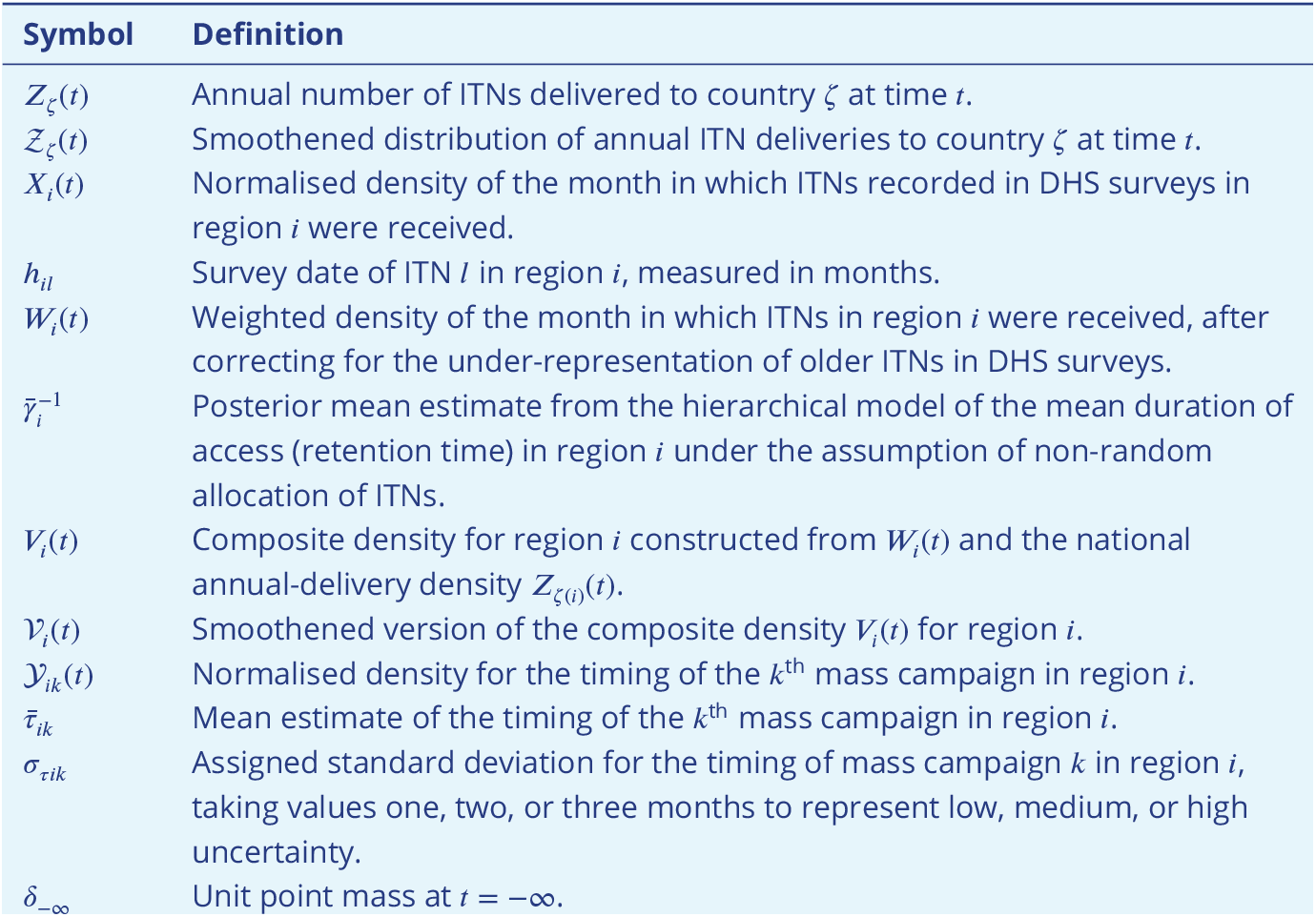
Additional notation used in appendix 3 beyond the core notation defined in appendix 1 - table 1.

**Appendix 1—table 4.**
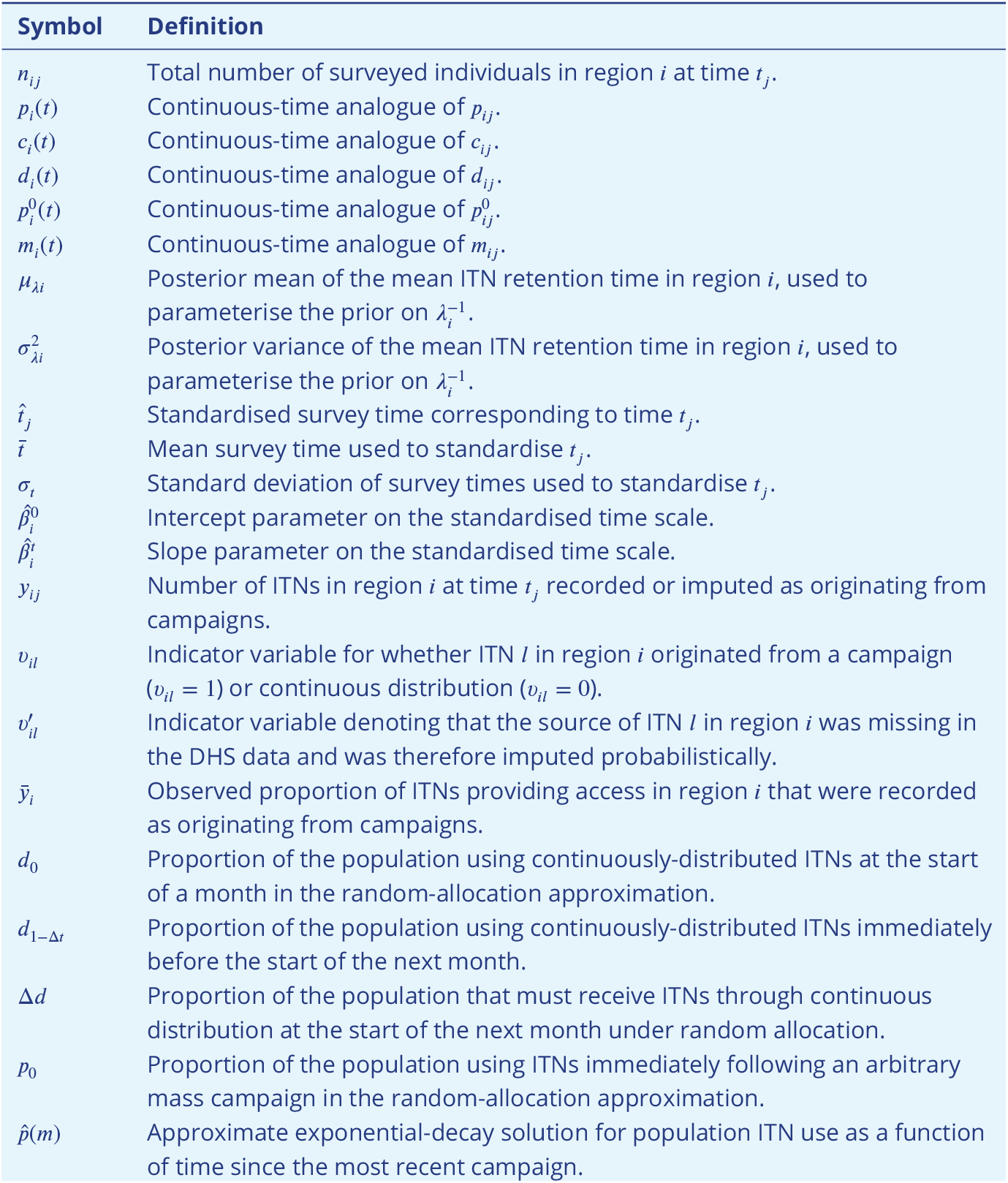
Additional notation used in appendix 4 beyond the core notation defined in appendix 1 - table 1.

**Appendix 1—table 5.**
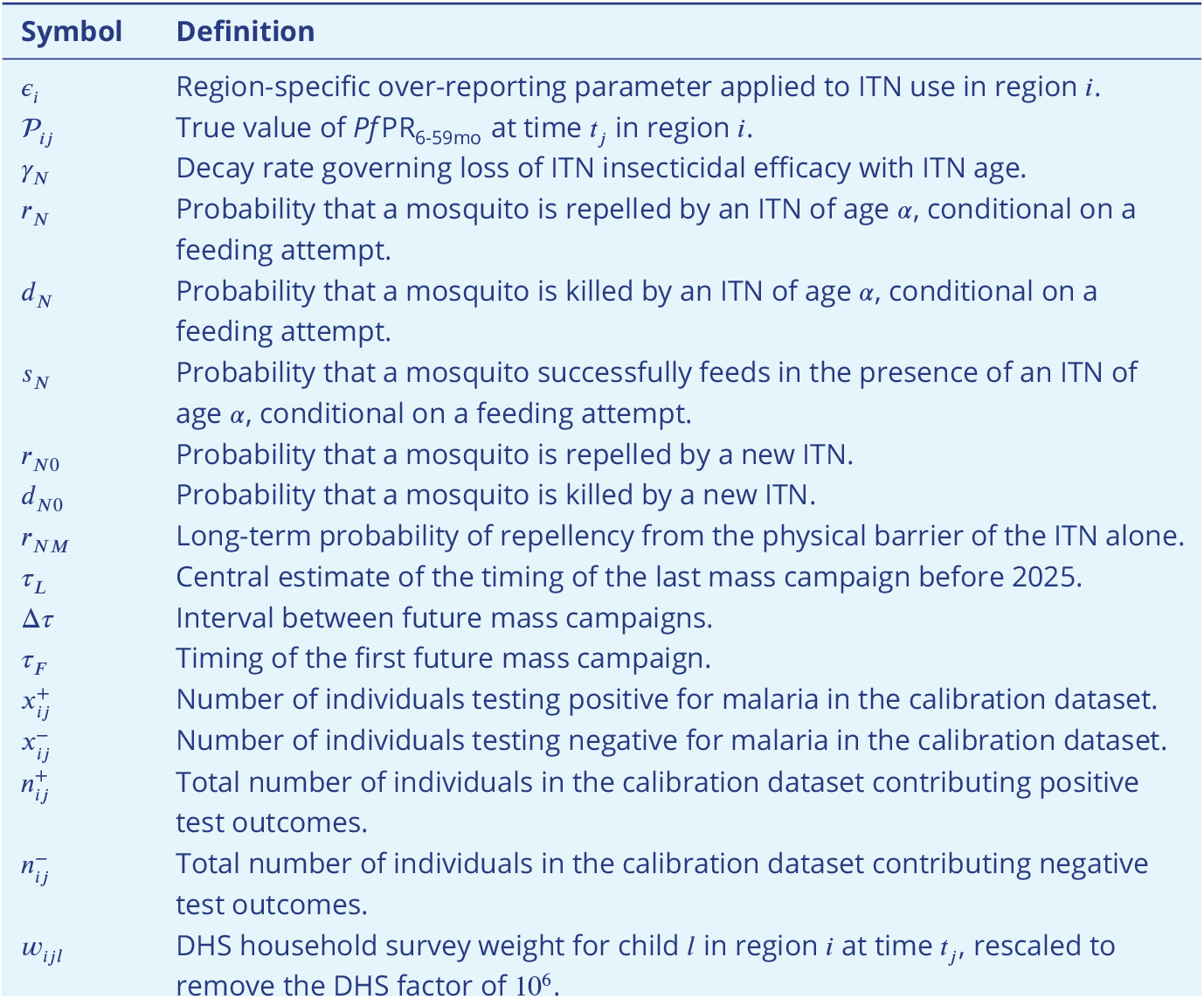
Additional notation used in appendix 5 beyond the core notation defined in appendix 1 - table 1.

| Symbol | Definition |
| --- | --- |
| $\epsilon_i$ | Region-specific over-reporting parameter applied to ITN use in region $i$. |
| $\mathcal{P}_{ij}$ | True value of $PfPR_{6-59mo}$ at time $t_j$ in region $i$. |
| $\gamma_N$ | Decay rate governing loss of ITN insecticidal efficacy with ITN age. |
| $r_N$ | Probability that a mosquito is repelled by an ITN of age $\alpha$, conditional on a feeding attempt. |
| $d_N$ | Probability that a mosquito is killed by an ITN of age $\alpha$, conditional on a feeding attempt. |
| $s_N$ | Probability that a mosquito successfully feeds in the presence of an ITN of age $\alpha$, conditional on a feeding attempt. |
| $r_{N0}$ | Probability that a mosquito is repelled by a new ITN. |
| $d_{N0}$ | Probability that a mosquito is killed by a new ITN. |
| $r_{NM}$ | Long-term probability of repellency from the physical barrier of the ITN alone. |
| $\tau_L$ | Central estimate of the timing of the last mass campaign before 2025. |
| $\Delta\tau$ | Interval between future mass campaigns. |
| $\tau_F$ | Timing of the first future mass campaign. |
| $x_{ij}^+$ | Number of individuals testing positive for malaria in the calibration dataset. |
| $x_{ij}^-$ | Number of individuals testing negative for malaria in the calibration dataset. |
| $n_{ij}^+$ | Total number of individuals in the calibration dataset contributing positive test outcomes. |
| $n_{ij}^-$ | Total number of individuals in the calibration dataset contributing negative test outcomes. |
| $w_{ijl}$ | DHS household survey weight for child $l$ in region $i$ at time $t_j$, rescaled to remove the DHS factor of $10^6$. |

## Appendix 2

### 2 Initial estimation of ITN retention times

Throughout this study, we assume that a net *l* in given region *i* is used continuously each night over some period of time, 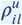, but may provide access for a different period, 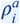. We also assume that nets in region *i* cease to be used and are disposed of by households at constant, but not necessarily equal, rates, 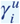 and 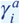, respectively, such that:

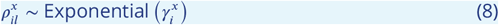

where *x* = *u* and *x* = *a* in the contexts of use and access, respectively. Unless otherwise stated, both use and access are treated as proportions of the population throughout. The expected mean duration of use and access (the retention time) of nets are assumed to be 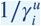 and 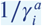, respectively. Here in section 2, ITNs are assumed to be distributed without replacement (i.e. only individuals without an ITN would receive one), though this is relaxed later when fitting our historical estimates of use and access (appendix 4), and when simulating future impact projections (appendix 5). However, throughout this study, the durations of use and retention time are always estimates of how long an individual continues to use or have access to a net in the absence of future replacement; estimates of these are therefore reflective of behaviour or ITN durability and not distribution patterns themselves.

The remainder of section 2 is presented for inference of the mean duration of access, or retention time. Aside from subsetting the data for nets that are used, instead of those that provided access, the methodology is otherwise identical for inferring the mean duration of use and is repeated independently in that context. We therefore drop superscript notation previously used to explicitly indicate access-specific parameters.

For clarity, the notation used in this appendix is summarised at the end in appendix 2 – table 2.

#### 2.1. Estimation of ITN loss from ITN age distributions

Here, we show how the pooled age distribution of ITNs providing access across DHS surveys can be used to estimate the decay rate of access 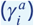 and in turn the mean retention time for a given region. If access is lost at a constant rate then the pooled age distribution of ITNs providing access over a sufficiently long period of time will be exponentially distributed, irrespective of how ITNs are distributed temporally, provided the rate at which they are replenished has a stable long-run average. As the relationship between the pooled ITN age distribution and the mean retention time is applicable for all regions, we drop subscript notation for clarity for the remainder of subsection 2.

We initially assume ITN distribution channels are long-standing such that population access follows some periodic equilibrium with period Δ*t*, which could be equal to mass campaign intervals. We assume ITNs are distributed without replacement whereby access is replenished through some time-dependent process, 𝒩 (*t*); this could be through periodic mass campaigns with supplementary continuous distribution, for example. The long-term mean replenishment rate of nets providing access is therefore:

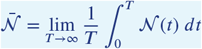

Given our assumption the process is at some periodic equilibrium, we note this is invariant to time-shifts:

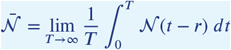

We now consider the proportion of the population at time *t* who have access to ITNs with ages, *α*, lying within some small interval. If *α* ∈ [*r, r* + Δ*r*], where Δ*r* is small, then the proportion of the population with access to ITNs of those ages is given by:

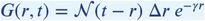

This can be inferred since approximately 𝒩 (*t* − *r*)Δ*r* of the population would have gained access from nets distributed during the interval [*t*−*r*−Δ*r, t*−*r*]. Then, due to the assumption of a constant rate of loss of access, a relative proportion of *e*^−*γr*^ would be expected to still have access to those nets at time *t*.

If we then pool all ITNs with ages between *r* and *r* + Δ*r* across all times when they may be observed, *t* ∈ [0, ∞), then the expected proportion of the population using an ITN of that age per unit time is:

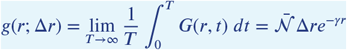

We can then infer the pooled age density of ITNs providing access with age *α* = *r* by normalising across all possible ages:

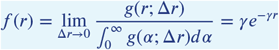

This therefore implies the long-term age-distribution (*α*) of ITNs providing access in the population follows the same distribution as the duration an ITN provides access (equation (8)):

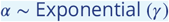

If we then consider a vector of observed ages, *α ∋ α*_*l*_, of nets that provided access pooled across a finite number of surveys, *n*_*s*_, conducted over an interval [*s*_0_, *s*_*m*_], then:

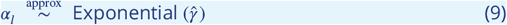

where:

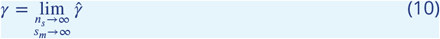

#### 2.2. Bayesian hierarchical model

Given equation (9), provided a mass campaign strategy is suitably long-established where a large number of surveys have been conducted over time, the mean retention time can be estimated from the mean of an exponential distribution fitted to the empirical age distribution of ITNs that provided access across all recorded DHS surveys. Additionally, the limit behaviour in equation (10) can be shown to hold both when surveys are conducted randomly or at regular intervals Δ*s*, provided that Δ*s* ≠ Δ*t* (Appendix 2 - figure 1). Although these methods are presented in the context of access, they are repeated independently for use.

**Appendix 2—figure 1.**
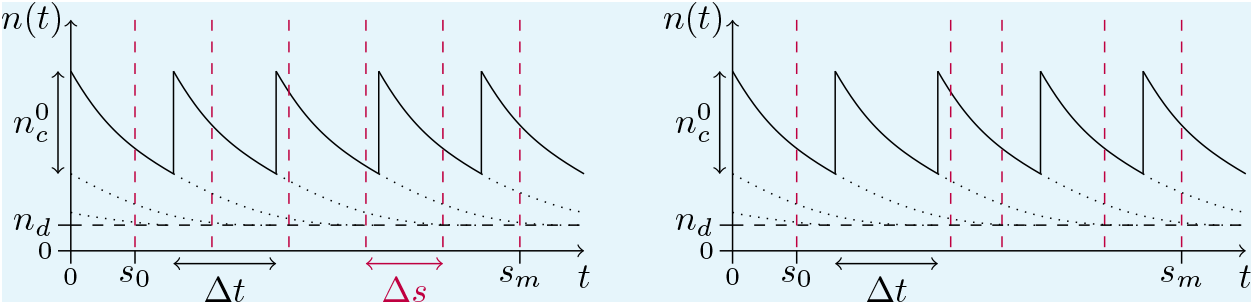
An illustration of the number of nets, *n*(*t*), in a region over time with continuous replenishment of routinely distributed nets, *n*_*d*_ , where 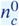 nets are distributed every Δ*t* years through regular mass campaigns (solid black lines). The contribution towards the total number of nets from routine distribution and previous mass campaigns are shown by the dashed and dotted blue lines, respectively. The timing of DHS surveys are shown by the red vertical dashed lines for both regular (left) and irregular (right) surveying.

In the case where Δ*s* = Δ*t*, the rate parameter in equation (9) will lead to biased estimates for the decay rate; if surveys are consistently conducted soon after a mass campaign, the empirical age distribution will be biased towards younger nets, such that the fitted rate parameter for the distribution in equation (9) would over-estimate the true decay rate; the converse would hold true when surveys are consistently conducted immediately prior to a mass campaign. Given the frequency of DHS surveys can vary significantly between countries, for countries with fewer surveys, there is an increased risk this may lead to biased estimates in decay rates. For example, if surveys are systematically conducted immediately after a mass distribution campaign, this will lead to over-estimates in the decay rates. Therefore, to attempt to reduce this risk of bias, we use a hierarchical model to pool information over larger spatial scales with the following postive-truncated normal (N_>0_) prior distributions:

##### Region-level prior

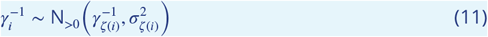

##### Country-level hyperpriors

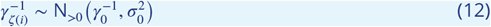

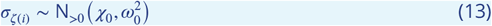

##### Continent-level hyperhyperpriors

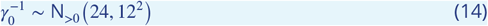

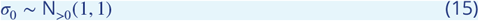

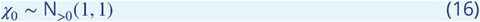

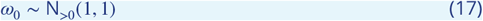

Under our hierarchical model, there is a country-level random effect on the region-specific mean ITN retention time (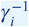, equation (11)). There is then a continent-level random effect on the country-specific mean ITN retention time (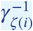, equation (12)), where *ζ* (*i*) is the country of region *i*. Standard deviations are modelled in an analogous manner using a hierarchical structure (equation (13)).

We then used a weakly-informative hyperhyperprior for the continent-wide mean retention time estimate (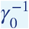, equation (14)), with a mean and standard deviation of 24 and 12 months, respectively. A prior mean of 24 months broadly reflects findings from previous studies (***Bhatt et al., 2015; Bertozzi-Villa et al., 2021***), yet equation (14) cautiously imposes a prior belief there is a 95% probability the continent-wide mean is between 3.9 and 47.6 months.

The between-country standard deviation in mean ITN retention time (*σ*_0_, equation (15)) is then assumed a priori to have a mean and standard deviation of 1 month. Since the central 95% range of a N_>0_ (1, 1) distribution is [0.08, 3.03], conditional on values of *σ*_0_ near the upper tail of this prior would allow roughly 95% of country-level means to lie within approximately six months of the continental mean a priori. This is broadly in line with previous estimates by ***Bhatt et al***. (***2015***) who estimated 85% (n=40) of sub-Saharan African countries had a median retention time between 1.5 and 2.5 years; the smooth-compact loss function assumed in that study leads to similar mean and median estimates. Imposing a marginally tighter inter-country variance than previous studies is nevertheless intentional. This is because the derivation that the age distribution of ITNs when pooled across surveys tends to the same distribution as the duration of continuous access assumes periodic distribution of ITNs is long-standing and that a sufficiently large number of surveys have been conducted without inherent bias in their timings relative to mass campaigns. While mass campaigns have been conducted for roughly an order of magnitude greater than expected ITN retention times, in countries where more Malaria Indicator Surveys (MIS), which are often conducted 6 months after a campaign, than other DHS surveys that are not timed in relation to campaigns, this may create an inherent bias that overestimates the decay rate of ITN use (and access). Assuming a random effect a priori with a slightly smaller variance than indicated in previous studies helps to mitigate bias arising from survey timings for data-sparse countries.

The mean (*χ*_0_, equation (16)) and standard deviation (*ω*_0_, equation (17)) of the inter-regional standard deviation of mean ITN retention times within each country (*σ*_*ζ*_) are also assumed a priori to have means and standard deviations of 1 month. Here *χ*_0_ and *ω*_0_ respectively govern the magnitude and variability of within-country heterogeneity in mean ITN retention times. Given the upper bound of the 95% range of these hyperhyperprior distributions is approximately 3 months, this would correspond to inter-regional standard deviations up to approximately 9 months in extreme cases where *σ*_*ζ*_ has values around *χ*_0_ + 2*ω*_0_. Conditional on such values, 95% of regional mean retention times would lie within approximately ±2*σ*_*ζ*_ ≈ ±18 months of the country mean. Although the hyperhyperpriors (which govern inter-regional variability) assumed in equations (16) and (17) are the same as the hyperprior (which governs between-country variability) assumed in equation (15), the model structure assumes random effects of greater magnitude between regions than between countries. As discussed, we have assumed a marginally narrower prior belief in inter-country variability than findings from previous studies (***Bhatt et al., 2015; Bertozzi-Villa et al., 2021***) to minimise the risk of survey timing biasing our estimates. Since DHS surveys and mass campaigns are often conducted at similar timings within a country, these biases are likely to be conserved across regions, allowing for greater scope to assume greater inter-regional variability a priori.

Under our assumption that nets are discarded at a constant rate within each subnational region *i*, the age, *α*_*il*_, of each net *l* is modelled as exponentially distributed with rate *γ*_*i*_. However, as DHS surveys do not record ages of nets greater than 36 months, but report them as being older than this threshold, we therefore treat these observations as right-censored at 36.5 months (due to whole-month rounding), giving a likelihood:

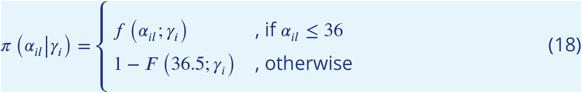

where *f* (*α*; *γ*) and *F* (*α*; *γ*) are Exponential density and cumulative distribution functions, respectively.

To correct for non-random probabilities of households being sampled in DHS surveys, we also conduct the fitting process using pseudo-counts, *M*_*ir*_, of the number of nets of a given age *α* = *r* in region *i*, where:

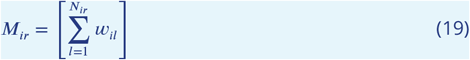

given *N*_*ir*_ is the surveyed number of nets of age *r* in region *i, w*_*il*_ is the household survey weight for net *l*, while [⋅] denotes a standard rounding function. As DHS surveys have a stratified sampling design each household has a unique probability of being sampled within each region *i*. To account for these non-random probabilities of households being surveyed, each net *l* that is recorded has an associated household weighting, *w*_*il*_. The DHS also scales these weightings by a factor of 10^6^; our notation here of *w*_*il*_ corrects for this scaling, and is therefore equal to the reported DHS household weightings divided by 10^6^.

After fitting our model in Stan with the R interface RStan (v2.32.6, ***Stan Development Team, 2024***), we generated 1,000 samples from each of the four MCMC chains after a burn-in period of 1,000 iterations. This simulated 4,000 draws from the posterior distributions of the mean duration of use and retention time at subnational, national, and continental scales.

Our subnational posterior estimates of the mean duration of use and retention time are then used in section 3 in estimating the timing of mass campaigns. They are subsequently used as informative priors and are re-fitted to observed use and access data in section 4. This subsequent re-fitting process aids in reducing potential biases arising from the relative timings of surveys and campaigns as discussed above, and further reduces the influence of prior assumptions made in the hierarchical model described in equations (11) - (13).

It should also be highlighted that while we have assumed an exponential loss function for the duration of use and access, other studies, including those by ***Bhatt et al***. (***2015***) and ***Bertozzi-Villa et al***. (***2021***), have utilised an “S-shaped” smooth-compact loss function. This was originally developed for the net procurement planning tool, NetCALC (available at: http://www.networksmalaria.org/networks/netcalc) (***Paintain et al., 2013***). This functional form was fitted to net age distribution data (*n* > 2500 nets) across 12 surveys in Uganda prior to 2013 and was selected on the basis that the pooled observed age density peaked around 2 years and was six-fold greater than the density for newly distributed nets (File S2, figure 2 in ***Paintain et al., 2013***). Meanwhile for this study, discounting peaks at 12, 24 and 36 months (due to whole year rounding), the empirical age distribution broadly appears to be monotonically decreasing from month six onwards when pooled across all DHS surveys and countries included in this study (figure 2.a). However, the slight peak at month six, which is two-fold greater than at month zero, may be an artifact of MIS surveys often being conducted six months post-campaign, which would bias the sampled data to include more nets of this age. In light of potential survey artifacts and the largely monotonic decrease in net age, there was limited evidence against the use of an assumed exponential loss function from the data used in this study.

**Appendix 2—figure 2.**
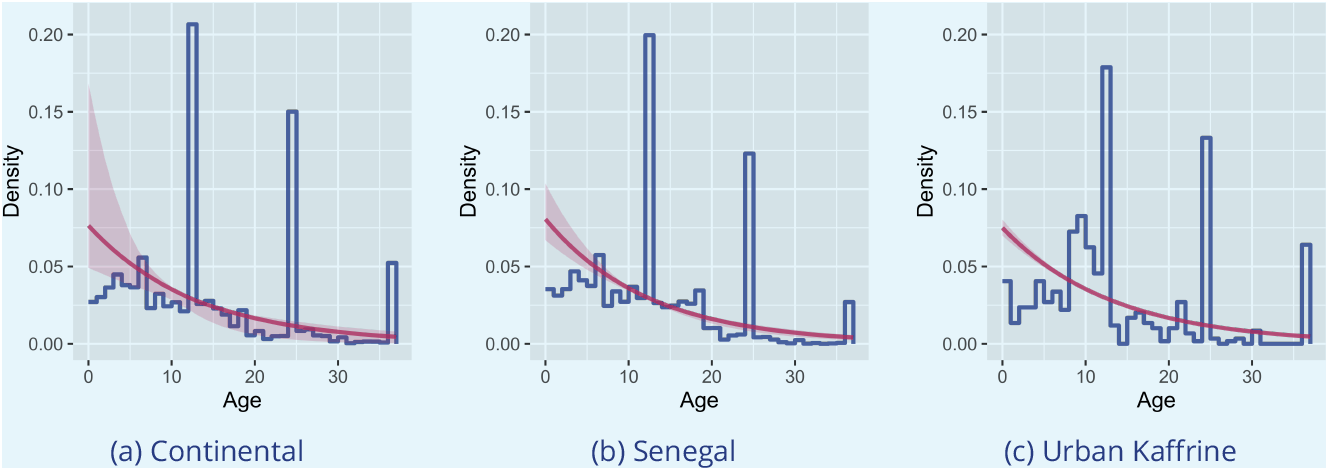
Posterior predictive distributions (red) from the hierarchical model for the age of a used net in months if one is sampled randomly over time and across either sub-Saharan Africa (a), or within an exemplar country (b) or subnational region (c). The shaded regions indicate 95% credible intervals, while the blue lines indicate the normalised empirical densities; these were generated from the pseudo-counts as described in equation (19). The peaks at 12, 24 and 36 months in the empirical densities are believed to be artifacts from the survey data due to individuals rounding reported ITN ages to the nearest whole year.

## Appendix 3

### 3 Mass campaign timings

#### 3.1 Estimating subnational timings

Prior to estimating changes in use and access over time subnationally, we first sought to estimate the timings of subnational mass distribution campaigns. We assume that for all subnational regions *i* within a country *ζ* (*i*), the *k*^th^ national mass campaign is conducted within a given time interval, [*r*_*ik*_, *s*_*ik*_] ∀ *i* where *ζ* (*i*) = *ζ* . However, we do not necessarily assume that a national mass campaign occurs in synchrony across all regions.

Using data on the annual numbers of nets delivered to a country, *Z*_*ζ*_ (*t*), that was compiled by the ***The Alliance for Malaria Prevention (2024***) Net Mapping Project, we applied Nadaraya–Watson kernel regression (***Nadaraya, 1964; Watson, 1964***) to generate smoothened distributions, Ƶ _*ζ*_(*t*). This was conducted using the ksmooth function from the R stats package (v4.3.2) for a bandwidth of 12 months due to the original resolution of the data being recorded annually (figure 1.a).

**Appendix 3—figure 1.**
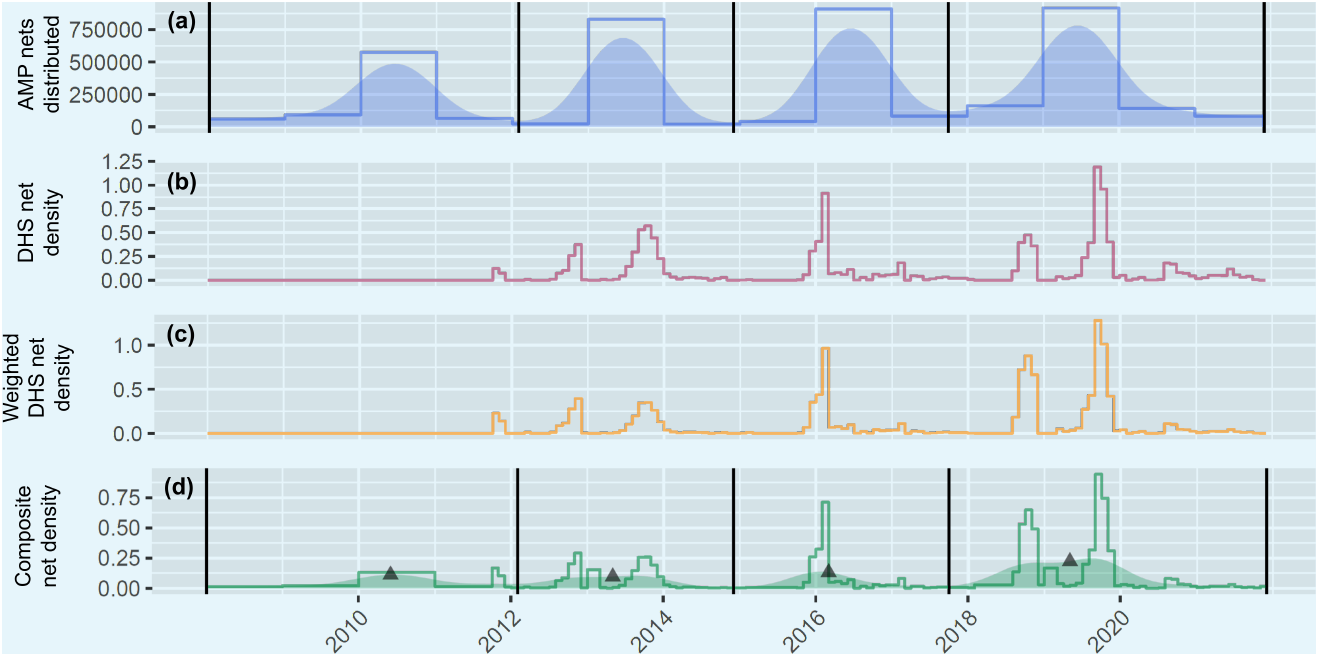
Estimates of mass campaign timings in the subnational Est region in Burkina Faso. Letting region *i* be the Est region of Burkina Faso, (a) shows the empirical, *Z*_*ζ*(*i*)_, and smoothened, Ƶ _*ζ* (*i*)_ , annual numbers of nets delivered nationally are shown by the solid blue line (as reported by the Alliance of Malaria Prevention, AMP) and shaded region. The local minima of Ƶ_*ζ*(*i*)_, which define the midpoints between net delivery dates are shown by the vertical black lines in (a) and (d) and indicate the assumed periods when mass campaigns are allowed to have occurred. The density proportional to the the number of nets received subnationally by month, *X*_*i*_, is shown in red (b). The weighted version of this density, *W*_*i*_, to account for older nets being under-represented in DHS surveys is shown in yellow (c). The composite density, *V*_*i*_, and its smoothened counterpart, 𝒱_*i*_ , which are constructed from the stepwise densities in (a) and (c) are shown by the green line and shaded region in (d). The densities, 𝒴_*ik*_ , are subsetted from 𝒱_*i*_ by the black lines. These are treated as approximations of the probability of a mass campaign occurring in each month, and the expected value for the timing of each *k*^th^ mass campaign are shown by the black triangles.

If a net were selected randomly across space and time within a country *ζ* , we treat Ƶ_*ζ*_ (*t*) as an initial approximation of the probability of the month in which that net was received after normalisation. Under our assumption that mass campaigns are conducted within defined nationwide time intervals, we therefore consider the local minima of the density Ƶ_*ζ*(*i*)_(*t*) to also be indicative of the local minima for the probability of a mass campaign occurring in any subnational region *i* of country *ζ* (*i*) at those times. If the *k*^th^ mass campaign in region *i* occurred at time *τ*_*ik*_, we therefore define the range of possible values of *τ*_*ik*_ such that Ƶ_*ζ*(*i*)_(*r*_*ik*_) and Ƶ_*ζ*(*i*)_(*s*_*ik*_) are respectively the local minima of Ƶ_*ζ*(*i*)_ immediately preceding and following mass campaign *k* given *r*_*ik*_ ≤ *τ*_*ik*_ < *s*_*ik*_ and *s*_*ik*_ = *r*_*i*(*k*+1)_.

To refine our estimates of when mass campaigns were likely to have occurred subnationally, we initially consider the recorded number of nets received by month in DHS surveys in subnational region *i* and normalise this to generate another density, *X*_*i*_(*t*) (figure 1.b). Given a total *N*_*i*_ nets were recorded across all surveys conducted in region *i*, we generated each density given the age, *α*_*il*_, and survey date, *h*_*il*_, in months for each net *l* ∈ {1, …, *N*_*i*_}:

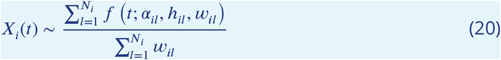

Where:

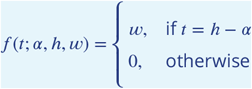

such that *w*_*il*_ is the DHS household survey weight for net *l* to correct for non-random probabilities of households being surveyed. However, since the density in equation (20) is generated from nets that were observed in households, this density will be systematically biased by younger nets, since there will be a greater probability that an older nets would have been discarded before it was recorded in a survey than a younger one. Under our assumption that ITNs are discarded at a constant rate, for every *w* ITNs surveyed of age *α*, an expected 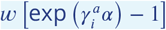 ITNs will have been lost between the date they were received and that of the survey. Therefore, to generate an unbiased density, *W*_*i*_(*t*) (figure 1.c), we use our previously generated a posteriori mean estimates of mean retention times, 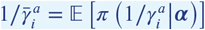, to scale the DHS survey weights by the expected number of ITNs that would have been received for each ITN that was surveyed:

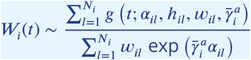

Where:

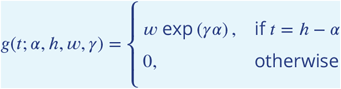

However, to account for months where no nets were recorded to have been distributed, we then generated a composite density with the stepwise normalised density, *Z*_*ζ*(*i*)_(*t*), that was proportional to the national number of nets distributed annually:

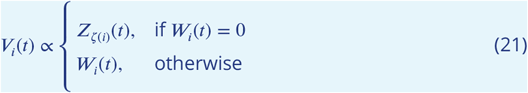

We again conduct Nadaraya–Watson kernel regression on this composite density for each subnational region using the ksmooth function in R with a bandwidth of 12 months to generate smoothened densities, 𝒱_*i*_ (*t*) (figure 1.d). Our rationale for this is two-fold; firstly, the composite density in equation (21) is proportional to the number of annual nets distributed for some months; secondly, as the age distributions of nets for most subnational regions has notable peaks at 12, 24 and 36 months (figure 2), it is probable that responses to this survey question were rounded to the nearest whole year by some participants.

After normalising the density 𝒱_*i*_ (*t*), such that ∑ _*t*_𝒱_*i*_ (*t*) = 1, this represents our best effort at capturing the probability of the month of distribution if a net is selected at random both in space and time within a subnational region. We then consider the probability density, 𝒱 _*i*_ (*t*), of the distribution date of a net selected randomly conditional on that date being in the interval [*r*_*ik*_, *s*_*ik*_] in which the *k*^th^ mass distribution campaign occurred:

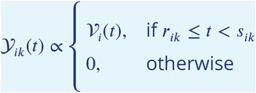

After discretising this density by month and by normalising such that ∑_*t*_ 𝒴_*ik*_(*t*) = 1, we then treat 𝒴_*ik*_(*t*) as a proxy for the probability of mass campaign *k* occurring at time *τ*_*ik*_ in region

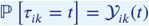

The expected value of the timing of a mass campaign is therefore estimated from:

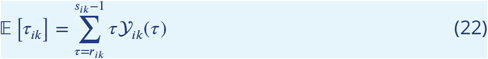

To account for uncertainty in the timing of mass campaigns, we then calculate the variance around the timing of each mass campaign:

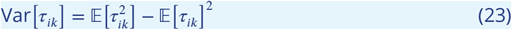

Given:

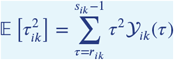

While this approach represented our best effort at capturing the central tendency of the subnational timings of mass campaigns, the variances calculated from equation (23) are likely to overestimate the degree of uncertainty in campaign timing. For example, for variance around the period of the 2019 mass campaign in Est, Burkina Faso, is far greater than in 2016 (figure 1.d). However, this is potentially driven by a DHS survey begin conducted at the end of 2021, which resulted in a notable number of ITNs being reported as 24 and 36 months old, due to rounding to the nearest whole year. We therefore propose that the peaks in the composite density in figure 1.d are primarily driven by whole year rounding, which has resulted in a large variance in the smoothened density that potentially overinflates the uncertainty in the timing of the 2019 mass campaign.

Due to the potential of over estimating the uncertainty in the timings of mass campaings due to whole year rounding in survey responses, we rank the variances calculated from equation (23) for all mass campaigns in all regions. We then split these into tertiles of equal size based on the variances calculated from equation (23) and assign standard deviations for the timings of mass campaign *k* in region *i, σ*_*τik*_ of one, two and three to represent mass campaigns where there is low, medium and high uncertainty in the timing of mass campaigns. We then approximate the uncertainty around our central estimates with a truncated-normal (TN) density with upper and lower bounds set at three standard deviations from the expected value:

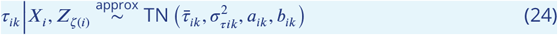

Where:

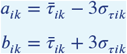

and 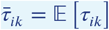 from equation (22). This gives a range of proposed dates for the area which is desirable as campaigns are likely to be spread out over a period given the geographical scale of the area and the considerable logistics involved. The proposed timings of mass campaigns were verified where possible though discussion with interested parties nationally and internationally, though further work to record the subnational timings of mass campaigns is encouraged.

#### 3.2. Time since the most recent campaign

If the time of the last campaign (*ϕ*_*ij*_) in region *i* before a survey at time *t*_*j*_ was known, then we could directly infer the months since the last campaign:

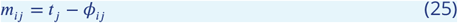

In contexts where national malaria programmes or other stakeholders have knowledge of the timings of mass campaigns (i.e. when there is no uncertainty in *ϕ*_*ij*_), the methodology can be adapted by deterministically evaluating the time since the last campaign (equation (25)) for each time point. However, the timings (*τ*_*ik*_) of each *k*^th^ campaign in each region *i* are unknown random variables. Although we do not know their exact timings, we captured our uncertainty of them previously through equation (24). Before we can probabilistically represent the months since the last mass campaign, we need to probabilistically estimate the timing of the last campaign relative to each time point *t*_*j*_ for each region *i*. Before proceeding, we note that for all regions, the supports [*a*_*ik*_, *b*_*ik*_] of the probability distribution (equation (24)) for the timing of each *k*^th^ campaign are non-overlapping:

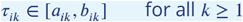

where:

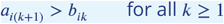

For times *t*_*j*_ < *a*_*i*1_ that precede the support [*a*_*i*1_, *b*_*i*1_] of the distribution for the first campaign, we therefore will have assumed a 100% probability that there was no preceding campaign. Therefore, the probability of using any net here is equal to the probability of using a continuously-distributed net, *p*_*ij*_ = *d*_*ij*_ , as per equation (31). We can also represent this as a unit point mass (*δ*_−∞_) for a campaign occurring at *t* = −∞, since 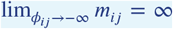 from equation (25) and 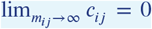 from equation (35). The distribution of the timing of the last campaign conditional on a 100% probability of there being no preceding campaign to *t*_*j*_ is therefore given by:

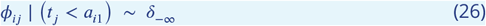

For the remainder of this section, given some time *t*_*j*_, we specifically use *k* to denote the index of the campaign in region *i* with an uncertainty window [*a*_*ik*_, *b*_*ik*_] that either contains (*a*_*ik*_ ≤ *t*_*j*_ < *b*_*ik*_) or immediately precedes (*b*_*ik*_ ≤ *t*_*j*_ < *a*_*i*(*k*+1)_) time *t*_*j*_. For times between the supports of the distribution (equation (24)) describing the timing of the last *k*^th^ campaign and the next (*k* + 1)^th^ campaign (i.e. when *b*_*ik*_ ≤ *t*_*j*_ < *a*_*i*(*k*+1)_), we will have assumed a 100% probability that the last campaign relative to *t*_*j*_ will be the *k*^th^ campaign. In this case, the probability distribution of the timing of the last campaign is the same as the timing of the *k*^th^ campaign:

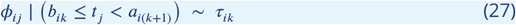

Now, let *F*_*ik*_ be the cumulative distribution function of the truncated normal distribution for the timing of the *k*^th^ campaign (equation (24)). For times within the uncertainty window of the first campaign (when *a*_*i*1_ ≤ *t*_*j*_ < *b*_*i*1_), there will be some probability *F*_*i*1_(*t*_*j*_) that the first campaign was the most recent; the remaining probability would be that there was no preceding campaign. In this case, the probability distribution of the timing of the last campaign will be:

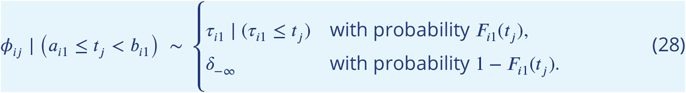

Extending this to the general case, for times within the uncertainty window of any other campaign (*k* ≥ 2), there will be some probability *F*_*ik*_(*t*_*j*_) that campaign was the most recent, with the remaining probability that the earlier campaign was the most recent:

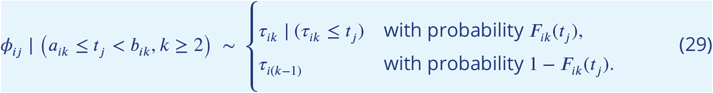

From equations (26)-(29), we can therefore describe the probability distribution of the timing of the last campaign relative to any time *t*_*j*_ in region *i* as follows:

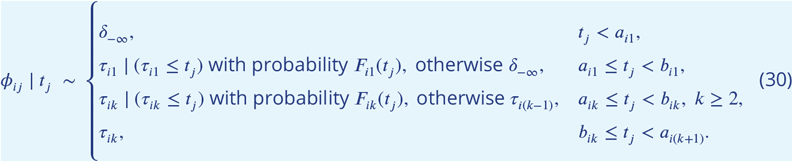

Then, given equation (30), the months since the last campaign for any general *t*_*j*_ can also be treated as a random variable and inferred directly from equation (25).

## Appendix 4

### 4. Historical use and access

The model presented in this section describes how we estimated historical trends in access. Aside from fitting the model to the number of individuals who used nets, *u*_*ij*_ , instead of those who had access, *a*_*ij*_ , in region *i* at time *t*_*j*_, the methodology is otherwise identical to how we estimated historical trends in use.

#### 4.1. Core model structure

An ITN is assumed to provide access for up to two people within the same household, except where more than two individuals were recorded as having slept under the same net; in such cases, an ITN is assumed to provide access to all individuals who slept under it the previous night. When fewer than two people sleep under an ITN, access is allocated at random among the remaining household members who did not use a net. When more than one ITN can provide access to an individual who did not sleep under one, the net deemed to provide access is also assigned at random.

Given the first mass campaign in region *i* occurs at time *τ*_*i*1_ (equation (24)), we compartmentalise the proportion of the population with access (*p*_*ij*_) into access provided by campaign-distributed ITNs (*c*_*ij*_) and continuously-distributed ITNs (*d*_*ij*_):

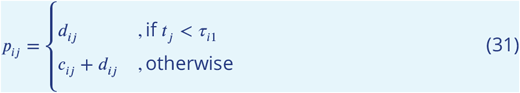

We then denote 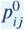 as the proportion with access immediately following a mass campaign. We assume this exhibits logistic-type growth over time towards some saturation threshold, *ψ*_*i*_:

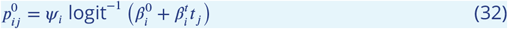

Immediately following any campaign, when 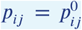, we assume the proportion of access attributable to campaign-distributed ITNs, relative to access from any ITN, is constant and given by 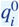. The proportions of the population with access to campaign-distributed and continuously-distributed ITNs immediately following a campaign are, respectively:

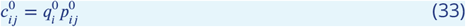

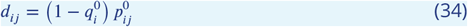

Since 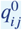 is assumed to be constant, we also implicitly assume access to continuously-distributed ITNs also exhibits logistic-type growth.

Where 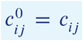 when the number of months following a mass campaign, *m*_*ij*_ = 0. We have also implicitly assumed continually-distributed nets are replenished in the population such that the proportion with access to them grows logistically over time since 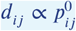.

However, as mass campaigns are conducted periodically, given we assume ITNs are discarded with a constant hazard rate, the proportion with access to a campaign-sourced ITN will decay over time following the last campaign:

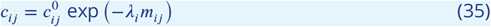

If ITNs are distributed without replacement (an assumption which is later relaxed), then the inverse of the decay parameter in equation (35), *λ*^−1^, is equal to the mean retention time. Given the proportion of the overall population with access to campaign ITNs decays over time, it follows that the proportion with access to a campaign ITN relative to the proportion with access to any ITN (*q*_*ij*_) will decrease over time following the last mass campaign:

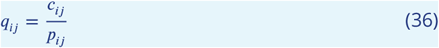

#### 4.2. Prior distributions

To fit our model, we use our previous posterior estimates of mean retention times given observed net ages from to inform strongly informative priors (equation (37)) of the mean retention time, with either non-informative or regularising priors for other parameters:

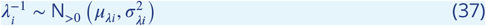

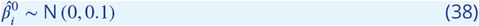

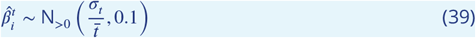

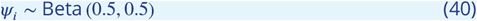

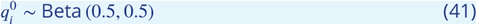

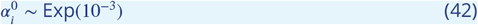

Given the posterior mean and variance of the mean ITN retention time conditional on observed ITN ages:

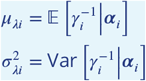

which are estimated from the sampled posterior distribution of mean ITN retention from the hierarchical model (equations (11) and (18)) described in section 2:

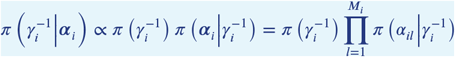

where 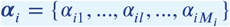 is the vector of ages of ITNs providing access in region *i*. Here, *M*_*i*_ denotes the total observed number of ITNs providing access of any age *r* after correcting for DHS sample weights such that *M*_*i*_ = ∑_*r*_ *M*_*ir*_ (equation (19)).

Meanwhile, survey times were standardised such that 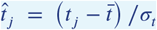 with 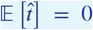 and 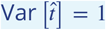. Therefore, on the logit scale of the probability of access immediately following a mass campaign relative to its maximum value (i.e. logit 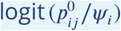, the intercept and slope parameters on the original time scale are therefore given by:

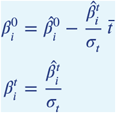

Priors on the standardized coefficients (equations (38) and (39)) were chosen to provide regularisation against implausibly abrupt changes on the standardized scale, while non-informative Beta (0.5, 0.5) priors were used for the maximum probability of access (equation (40)) and the conditional probability access is provided by a campaign-sourced ITN immediately following a campaign (equation (41)).

Finally, we use a regularising Exponential(10^−3^) prior for the overdispersion parameter 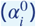, which controls subregional heterogeneity in access and is discussed further in Section 4. A flat or more diffuse prior was avoided, as it would assign disproportionate weight to extremely large values of 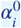. Our choice centres prior belief on low overdispersion 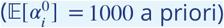, this corresponds to a maximum subregional standard deviation in access of approximately 1.6%. However, it retains support for substantially lower values of 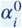, which correspond to higher overdispersion and greater subnational variability in access, where suggested by the data.

#### 4.3. Likelihood specification

Before formulating our likelihood model, we note there are two sources of data from the DHS that can be used, in addition to the previous posterior inference made on retention times given the surveyed ages of nets.

Firstly, given *n*_*ij*_ surveyed individuals in region *i* at time *t*_*j*_, we can construct the likelihood from the probability of observing the number of individuals with access, *a*_*ij*_ , given our parameters:

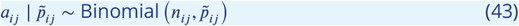

where the probability of having access to an ITN at time *t*_*j*_ is denoted by 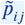. However, we do not assume this is equal to the proportion of the population (*p*_*ij*_) with access to ITNs for all individuals. There is likely to be greater variability in access to, and indeed use of, ITNs both within subnational regions and over time; for example, differing access between districts within subnational regions or differing use between rainy and dry seasons. These potential additional sources of uncertainty are not directly accounted for in our discrete-time model of historical access (and use) (equations (31) and (36)). Focusing again on access, to account for differences in the probability of access between individuals in the same region, we assume the probability of access at a given point in time can be described by a Beta distribution:

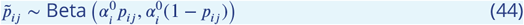

equations (43) and (44) can then be decomposed to a Beta-Binomial likelihood. The mean probability of individual access across a region at a given point in time is then equal to the overall proportion with access:

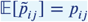

Meanwhile, the overdispersion parameter, 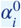 (unrelated to the notation for ITN age), controls the variability of the probability of individual access around the mean:

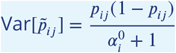

Such that smaller values of 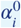 imply greater variability in the probability of individual access. While this variance will change as a function of the proportion with access, we assume the overdispersion parameter is constant for each region over time.

Secondly, given those individuals with access, we can construct the likelihood that nets providing access were distributed through campaigns, given the total number of ITNs that were recorded, or imputed, as having originated from campaigns, *y*_*ij*_ :

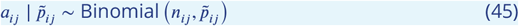

For each ITN *l* that provided access in region *i*, we assign an indicator variable, *υ*_*il*_, depending on whether the net originated from a campaign (*υ*_*il*_ = 1) or from continuous distribution (*υ*_*il*_ = 0). For the purposes of this study, we treat an ITN with any recorded source other than a campaign as having originated from continuous distribution channels. Sources either recorded as missing or ‘NA’ in DHS surveys are treated as missing data, and are denoted with 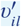. Letting 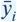 be the observed proportion of ITNs from campaigns that provided access in region *i*, for each net where the source was not recorded, we probabilistically impute whether they originated from campaigns:

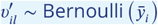

#### 4.4. ITN retention under random allocation

The model described in equations (31) - (35) describes how use and access change over time in different subnational regions. In itself, this makes no assumptions on whether probability an individual will receive a net is independent of whether an individual already has access to, or is currently using, a net. If ITNs are assumed to be distributed without replacement such they are only distributed to individuals without access to a net, then the inverse of the decay parameter *λ* (equation (35)) will be equal to the mean retention time of a net (or the mean duration of use when the model is applied in that context). However, for simulating transmission dynamics, we make the conservative assumption that ITNs are distributed randomly to the population. Therefore, we assume individuals are equally likely to start using, or gain access to, a net following a campaign or continuous distribution event irrespective of whether they are currently using of have access to one. As before, we will describe the remainder of this section in the context of access, though the same also applies for use.

While our use and access model (equations (31) (35)) is discretised by month to facilitate fitting to DHS data, our transmission dynamics model is simulated with the *malariasimulation* (v1.6.0) R package (***Charles et al., 2024***), which discretises a continuous-time model over daily timesteps. The transmission dynamics model only explicitly models use, and so the remainder of this section is described in that context.

Dropping region-specific subscripts for notational ease, the continuous-time model for the proportion of the population with use, which we wish to replicate in our transmission dynamics model is:

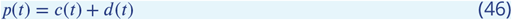

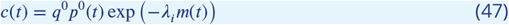

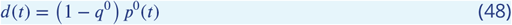

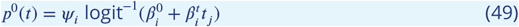

where all variables and parameters are as defined under the discrete-time model for the period January 2008 to December 2024, with time dependence explicitly indicated.

To reduce computational overheads, we simulate the distribution of ITNs at the start of every month in our transmission dynamics model. If we consider a region with no mass campaign distribution (when *q*^0^ = 0), then if there is a proportion *d*_0_ = *d* of the population using ITNs at the start of some month, then some small Δ*t* immediately prior to the start of the next month the proportion of the population using ITNs will be:

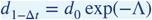

where Λ^−1^months is the mean duration of use (under our assumption of random allocation of ITNs) and the underlying logistic-type growth in continuous use of nets is assumed to be sufficiently small over one month (i.e.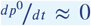). If we then assume random allocation of nets at the start of the next month, for the expected use of ITNs to reach the same level as the preceding month such that 𝔼[*d*_1_] = *d*_0_, we require ITNs to be distributed to a proportion Δ*d* of the population where:

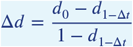

We now consider the counterfactual with mass campaigns occurring in the same region. If a campaign is conducted at some month (*m* = 0) such that use immediately after it is *p*_0_, then by the end of that month (*m* = 1 − Δ*t*) it follows that use would be:

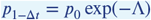

This can alternatively be described by the following initial value problem, again assuming underlying logistic-type growth occurs over much longer timescales than the decay in use following a campaign:

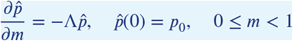

Then, at the start of the next month (*m* = 1), following a top-up distribution of ITNs through continuous channels that are randomly allocated to the population, the expected population use is:

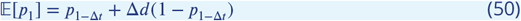

Following substitution and some algebra, it can be shown that if the mean duration of use is assumed to be the same when ITNs are distributed with replacement as without (i.e. *λ*^−1^ = Λ^−1^), then the expected population use (equation (50)) following a continuous distribution top-up will underestimate the target value (equation (46)) since 𝔼[*p*_1_] < *p*(1). An improved approximation can instead be found by approximating the change in use from equation (46) as an exponential decay given:

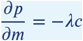

and so letting:

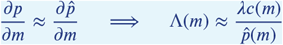

Then, applying this approximation immediately following a campaign:

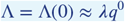

Therefore, the mean duration of use (under the assumption of random allocation of use with replacement) can be estimated from:

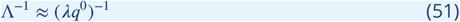

Although this is still an approximation, it can be shown to be a closer one than Λ^−1^ ≈ *λ*^−1^ provided the following condition is satisfied:

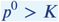

where:

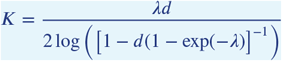

and for small *λ*:

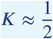

which is approximately true when *λ* is restricted to realistic values; for example, it can be shown 0.5051 < *K* < 0.5105 for all possible values of *d* when *λ* = ^1^/24 month^−1^. Over the recent period of interest (i.e. since December 2024), use immediately following a campaign is likely to be notably greater than 50% in all regions, and so we report our estimated mean durations of ITN use for different regions using the approximation given by equation (51). The same approach is also repeated in the context of access to give estimates of the mean ITN retention time.

## Appendix 5

### 5. Simulating different strategies

#### 5.1. Over-reporting of ITN use

While we fitted the discrete-time model of historical access and use directly to observed DHS data, hereafter we accounted for the potential of self-reported ITN use through DHS surveys to be greater than objectively measured use. A meta-analysis by ***Krezanoski et al***. (***2018***) estimated self-reported ITN use is typically 8% (95% CI: 3, 13) higher than the true value, or a 13.6% relative overestimation. As the majority of regions were estimated to have levels of use greater than 90% immediately following a campaign by 2024 under our model, and since use following mass campaigns is estimated to have meaningfully increased over 2018-2024 for most regions, we take a conservative approach in accounting for over-reporting and purposefully treat the absolute values estimated by ***Krezanoski et al***. (***2018***) as relative values to the reported use. The scaled probability of use, 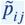 is herein assumed to be:

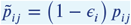

Where:

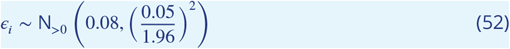

This over-reporting assumption is applied for both campaign-distributed and continuously-distributed ITNs. While we do not assume a similar systematic discrepancy between surveyed access and true access, the over-reporting assumption applied to use is accounted for in our calculations of use given access.

#### 5.2 Region-specific calibration

Future projections were conducted using *malariasimulation* (v1.6.0) (***Charles et al., 2024***), which encodes an individual-based *P. falciparum* transmission dynamics model. The core structure and parameterisation of the model are described in ***Griffin et al***. (***2010***, 2014) and are detailed in full at https://mrc-ide.github.io/malariasimulation/index.html. Aside from ITN-interventions, transmission dynamics models were calibrated for rural and urban settings in each subnational unit using the *site* (v0.2.2) (https://mrc-ide.github.io/site/) (***Winskill, 2024b***) R package, which has compiled location specific data from multiple sources for 2000 to 2024. *site* characterises local human population structure from WorldPop population estimates (***Tatem, 2017***), and vector abundance and distribution by species from ***Malaria Atlas Project (2024***) estimates. *site* has also compiled treatment rates and coverage of non-ITN interventions, including indoor residual spraying of insecticide (IRS), seasonal malaria chemoprevention (SMC), drug treatment and vaccination from historical estimates by ***Malaria Atlas Project (2024***) (MAP), ***DHS STATcompiler (2022***), ***ACCESS-SMC (2022***), ***SMC Alliance (2022***), and the Malaria Vaccine Implementation Programme (***Hamel, 2021***). Model seasonality is characterised (***Winskill, 2024b***) by fitting Fourier series to CHIRPS (***Funk et al., 2014***) daily rainfall data for each administrative-one unit. A linear relationship following a 30-day delay between the rainfall Fourier series and larval carrying capacity is assumed in each location following ***White et al***. (***2011***). Administrative-one level estimates of pyrethroid resististance have been estimated using spatio-temporally distributed bioassay mortality data (***Winskill, 2022***).

After accounting for population structure and non-ITN interventions, we calibrated the baseline transmission intensity (i.e. prior to the introduction of control interventions) using the *cali* (v1.0.8) R package (https://mrc-ide.github.io/cali/) (***Winskill, 2024a***). *cali* utilises an optimisation algorithm to minimise the sum of absolute differences between a desired prevalence series and one predicted by a model. For this process we fitted our historical model estimates of *Pf*PR_2−10_ to median estimates of the mean annual *Pf*PR_2−10_ from the ***Malaria Atlas Project (2024***) over 2005 - 2024. Since the relationship between baseline EIR and *Pf*PR_2−10_ here is specific to malariasimulation, MAP uncertainty estimates were not propagated through to our estimates in baseline EIR since these would not faithfully represent its true uncertainty. As a means of validation, we compared our model estimates of *Pf*PR_6-59mo_ to estimates from DHS surveys (for example, figure 2.B). We have also quantified the uncertainty in the estimate of the true *Pf*PR_6-59mo_ values (𝒫_*ij*_) attributable to the number of children tested in region *i* at time *t*_*j*_. Using a conjugate Beta-Binomial model with a non-informative Beta(0.5, 0.5) prior, the posterior distribution of 𝒫 is:

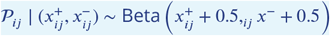

Where 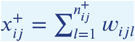, and 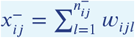, given 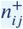 and 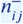 are the number of 6-59 month olds who tested positive and negative, respectively, at time *t*_*j*_ in region *i* and *w*_*ijl*_ is the scaled DHS sample weight for child *l* such that 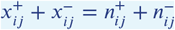.

#### 5.3. ITN efficacy parameters

The insecticidal activity of an ITN is assumed to decay at a constant rate, *γ*_*N*_ . Conditional on a feeding attempt on a human using an ITN of age *α*, the probability that a mosquito successfully feeds (*s*_*N*_), or is repelled (*r*_*N*_) or killed (*d*_*N*_) by that ITN is described by:

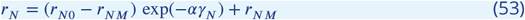

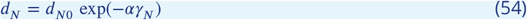

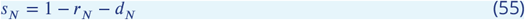

Where *r*_*N*0_ and *d*_*N*0_ are the respective probabilities of repellency and induced mortality of a new ITN, while *r*_*NM*_ is the long-term probability of repellency from the physical barrier alone. Following a systematic review by ***Nash et al***. (***2021***) that characterised the relationship between the level of resistance in wild mosquito populations, using the probability of bioassay survival as a proxy, and the probability of surviving 24 hours following exposure to a pyrethroid-only ITN, ***Sherrard-Smith et al***. (***2022***) estimated the parameters in equations (53) (55) for a range of pyrethroid-resistance levels. This was conducted for both pyrethroid-only and pyrethroid-PBO ITNs from a suite of paired Experimental Hut Trials. This work has since been updated by ***Churcher et al***. (***2024***) in the context of pyrethroid-chlorfenapyr ITNs and the models are broadly able to recreate the observed epidemiological benefit of different classes of ITNs evaluated in two cluster randomised control trials. Given estimated levels of pyrethroid resistance for each location over time, the probabilities of repellency and induced mortality for pyrethroid-only, pyrethroid-PBO and pyrethroid-chlorfenapyr nets were parameterised for our transmission dynamics model accordingly.

#### 5.4. Future case projections

For each subnational region, 100 realisations were simulated for all ITN distribution strategies considered (biennial and triennial mass campaigns with pyrethroid-only, -PBO and - chlorfenapyr ITNs) for a population of 100,000 individuals. We initialise all transmission models in January 2000 for each location, and assume continuous distribution over 2000-2007 at the same level as our historical estimates for January 2008. We then utilise our historical estimates of use from 2008 until our central estimates of the time of the last campaign, *τ*_*L*_, before 2025. Given a mass campaign interval of Δ*τ*, the timing of the first campaign of future projections, *τ*_*F*_ , is then drawn randomly from the interval *τ*_*L*_ +Δ*τ* ± 6 months. Subsequent mass campaigns occur after every Δ*τ* interval thereafter. Pyrethroid-PBO ITNs are assumed to be distributed prior to 2025 through both continuous and mass campaign channels. All ITNs distributed from *τ*_*F*_ onwards are assumed to be of the new ITN type, again in both continuous and mass campaign contexts. Continued logistic growth in the number of ITNs distributed through continuous channels or in total use following a mass campaign is no longer assumed beyond 2024, modifying equation (32) accordingly such that:

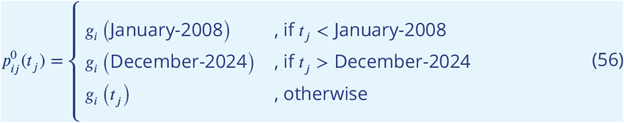

where:

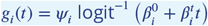

As ITN use is ultimately responsible for epidemiological impact, *malariasimulation* does not explicitly model access. For each realisation, values of 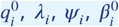 and 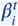 were therefore drawn from the joint posterior distribution from our discrete-time model of historical use (equation (31)), which was fitted using RStan (v2.32.6, ***Stan Development Team, 2024***). Our estimates of the subnational variability in the probability of use and access (equation (44)), are captured in our posterior distributions of the overdispersion parameters, 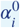. Our discrete-time model cannot distinguish between spatial and temporal subregional variability. In practice, temporal heterogeneity is likely to be more significant for use than access due to seasonal fluctuations (for example, use is likely to vary more between dry and wet seasons). To ensure compatibility with *malariasimulation*, for our transmission dynamics simulations we assume ITNs are used continuously before they cease to be used. Due to this incompatibility, subnational variability in the probability of use is therefore captured through stochastic binomial sampling in our transmission dynamics simulations rather than sampling from our posterior distribution for 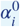. The degree of over-reporting of use is simulated from equation (52). Uncertainty in ITN repellency and mortality parameters (equations (53) and (54)) is also propagated forward to this study by simulating random draws from previous posterior distributions (***Sherrard-Smith et al., 2022; Churcher et al., 2024***) across each distribution event and realisation. Every 100 realisations for each scenario is simulated over 2000 to 2030 with the mean cases averted over the final nine years recorded. We then restrict this to a period of six years following the first mass campaign under future projections allow direct comparisons of cases averted between two triennial or three biennial campaigns; median time windows are used for future projections in the absence of mass campaigns. The maximum use estimated to be achieved by a campaign, had one been conducted at the end of 2024, is assumed to be maintained under future projections for each region. Similarly, the level of use of continuously-distributed ITNs estimated at the end of 2024 for each region is assumed to be maintained. It is assumed that for a particular subnational region, the use achieved after a biennial campaign is the same as that under triennial campaigns. We generate estimates of clinical cases under different distribution strategies by simulating campaign intervals of two and three years, in addition to the cessation of mass campaigns entirely, over the six-year projections. When switching from triennial to biennial campaigns, the model assumes those already with access are not excluded from mass campaigns, and accounts for diminishing returns in access as nets-per-capita increases (***Bertozzi-Villa et al., 2021***). It also assumes that use given access changes in the same manner to what was observed in triennial campaigns. All ITNs are assumed to be of the same type within a subnational region, and remain the same over subsequent mass campaigns. Routine distribution of ITNs, IRS, SMC and the level of drug treatment is assumed to continue as normal at the observed level. Posterior predictive estimates of cases averted are calculated from the additional cases averted over a baseline scenario where no future ITNs are distributed beyond our historical estimates (and those with nets loose them over time).

---

**Figure 2—figure supplement 1.**
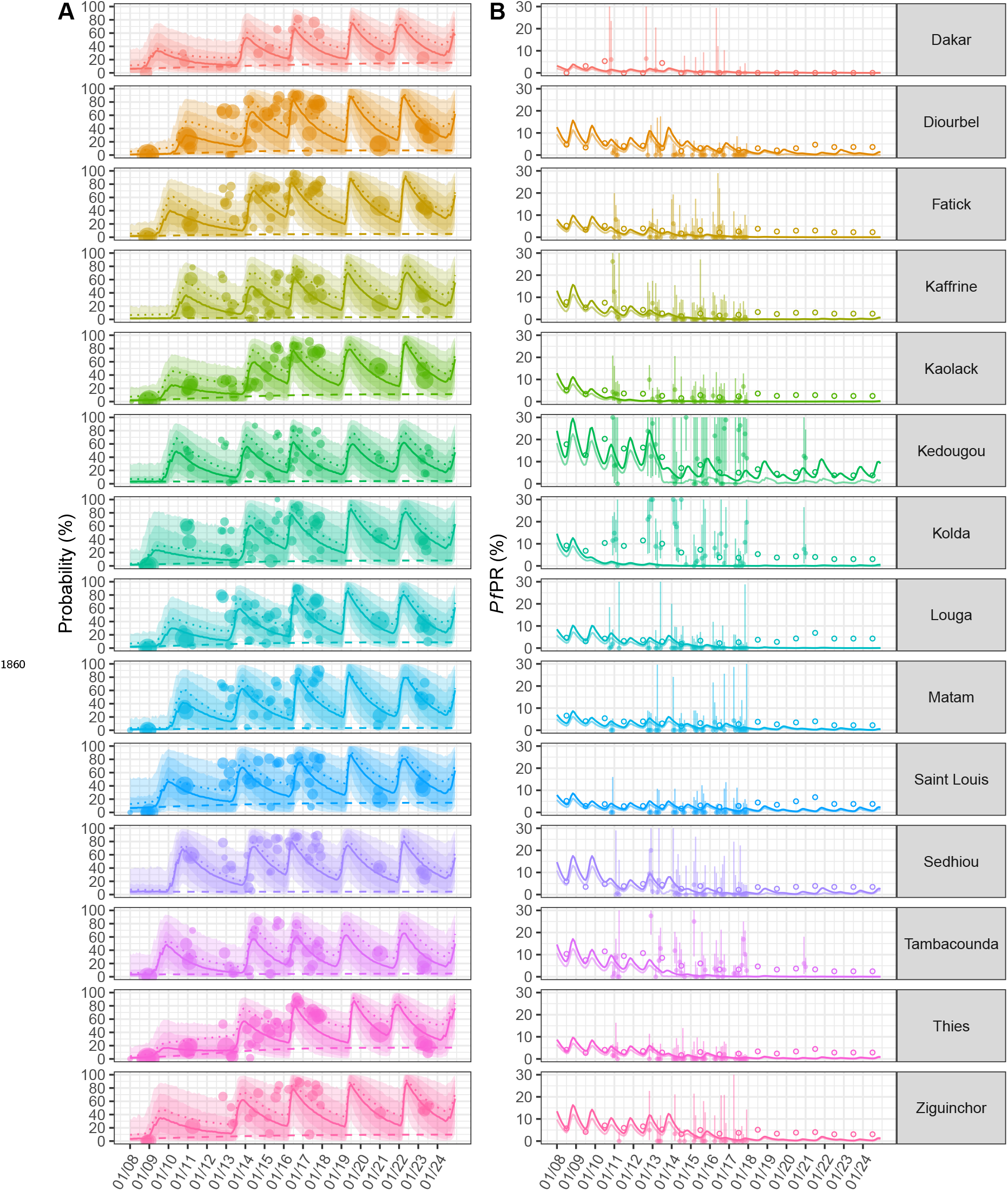
Use, access and *Pf*PR over time in rural Senegal. Model estimates in a lower transmission context in Senegal are shown. All features remain unchanged from figure 1. Note, in **B**, some credible intervals extend beyond the y-axis limits, which have been restricted to 30% to aid visibility.

**Figure 2—figure supplement 2.**
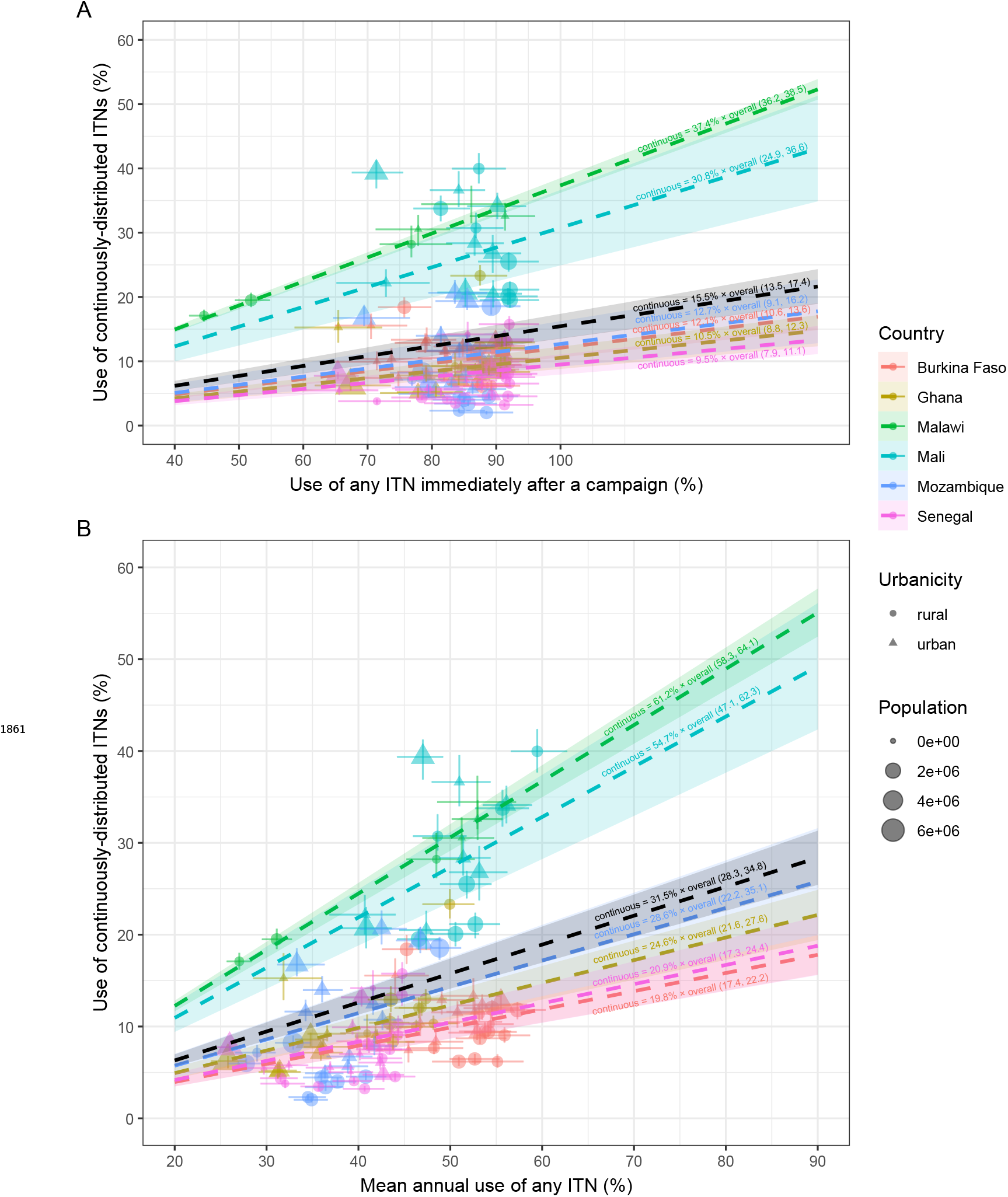
Use of ITNs from continuous channels. For each subnational region, coloured points indicate median estimates of the mean use of any ITN, and of those sourced from continuous channels, immediately following a mass campaign **(A)** and over the subsequent three years **(B)** with associated 95% credible intervals. For each country, the proportion of ITNs that were used which were sourced from continuous channels immediately following a campaign **(A)** and over the subsequent three years **(B)** can be estimated from the slopes of the coloured population-weighted linear regression lines, which assume a zero intercept. Estimates over all countries are shown by the population-weighted linear regression line in black.

**Figure 3—figure supplement 1.**
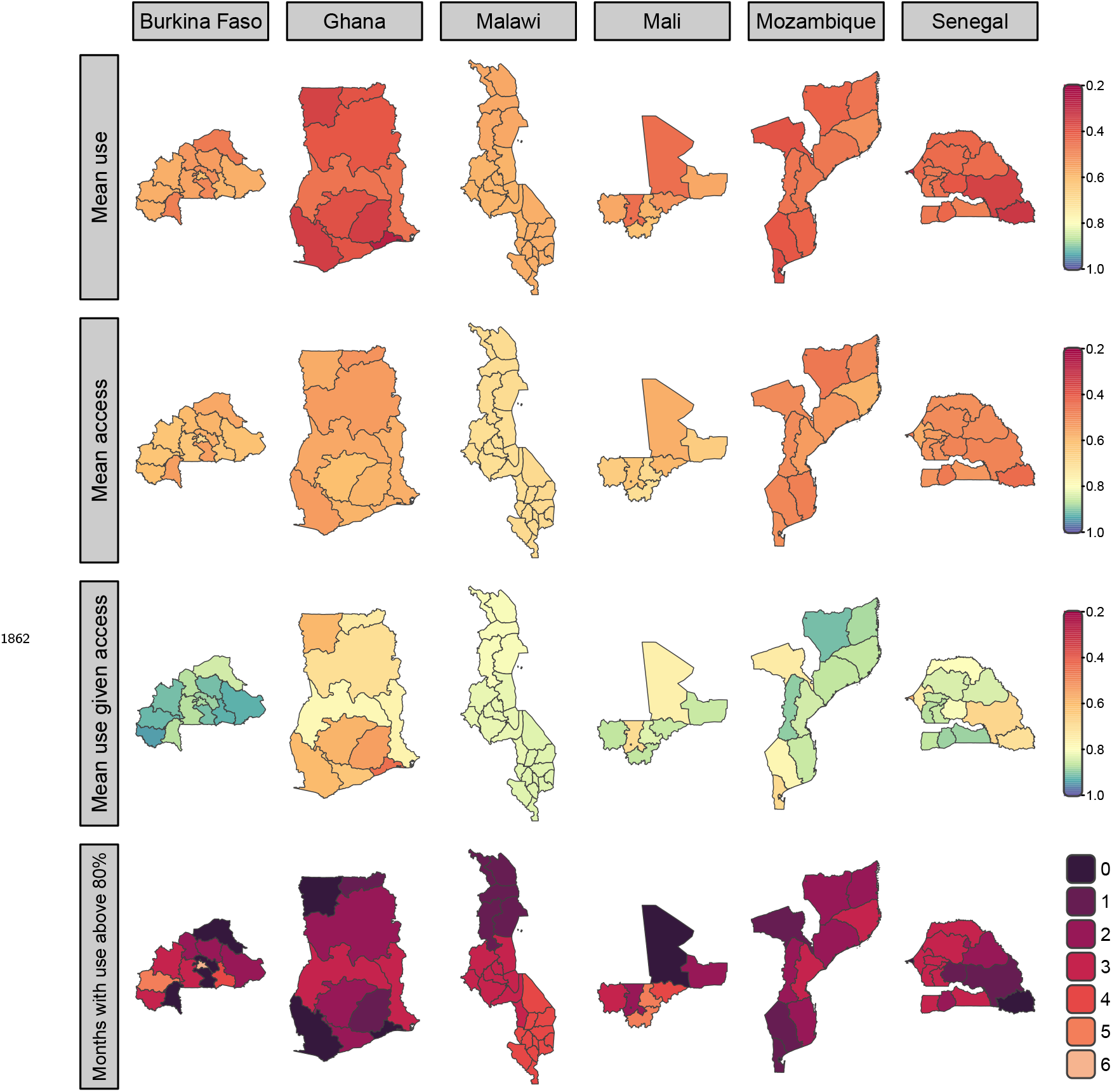
Mean use and access with three-year campaigns in subnational urban areas. Central estimates of mean overall proportion of people using an ITN the previous night, access to an ITN, and use given access for three-year mass campaign intervals are shown in the top three rows. The number of months following a mass campaign where overall ITN use exceeds 80% are shown in the bottom row.

**Figure 3—figure supplement 2.**
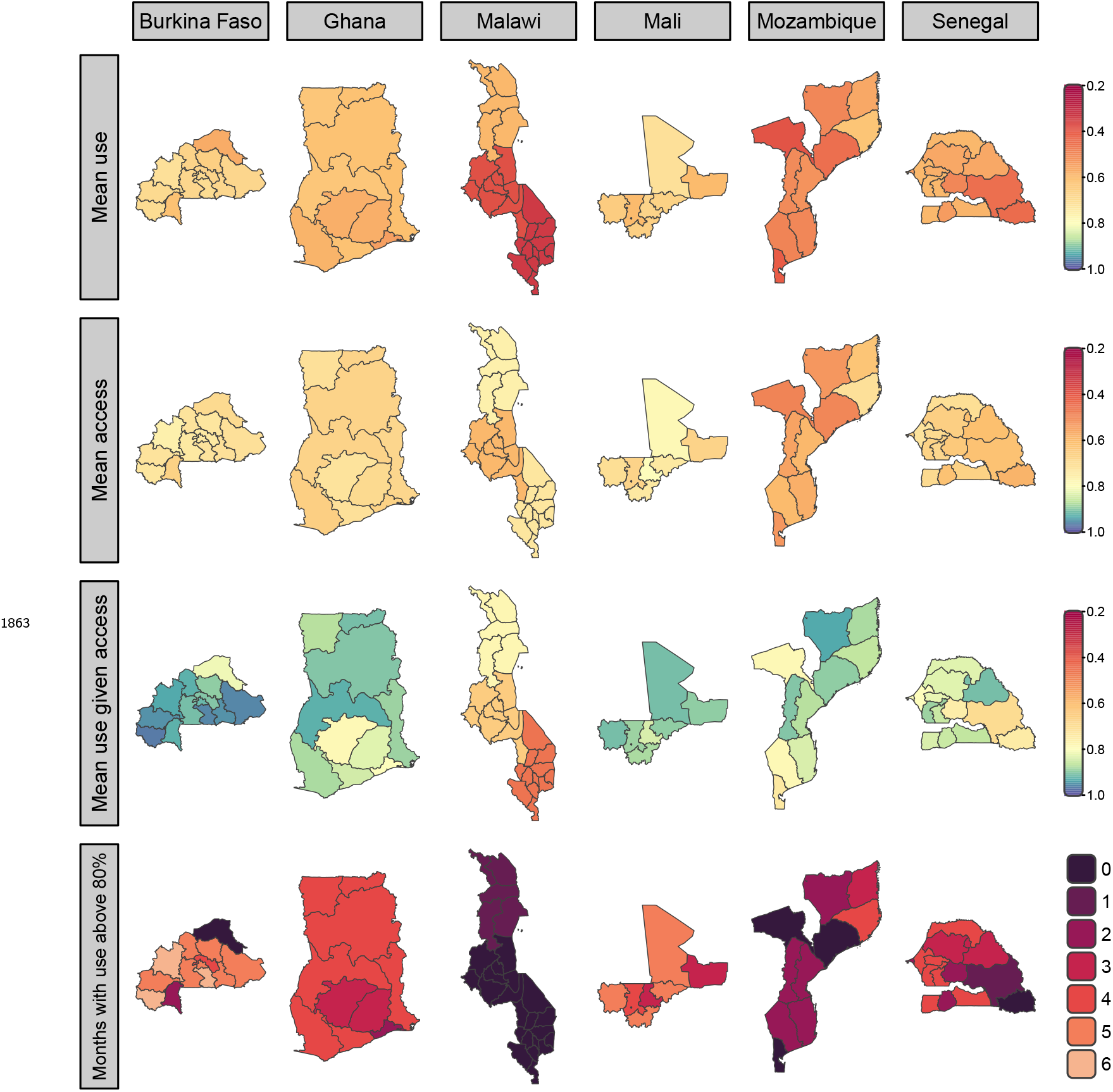
Mean rural use and access with two-year campaigns. Central estimates of mean overall subnational ITN use, access and use given access for two-year mass campaign intervals in rural areas are shown in the top three rows. The number of months following a mass campaign where overall ITN use exceeds 80% are shown in the bottom row.

**Figure 3—figure supplement 3.**
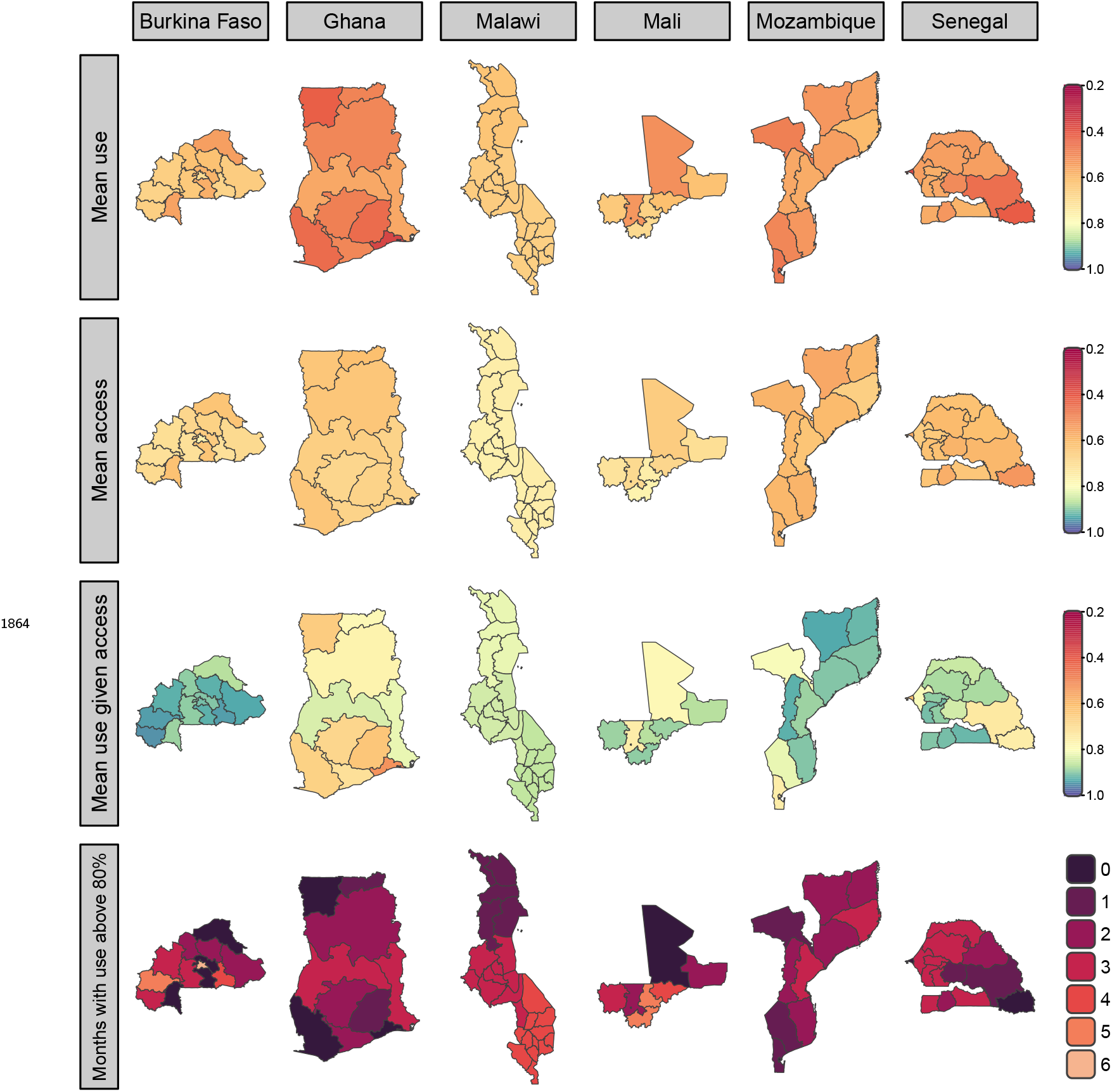
Mean urban use and access with two-year campaigns. Central estimates of mean overall subnational ITN use, access and use given access for two-year mass campaign intervals in urban areas are shown in the top three rows. The number of months following a mass campaign where overall ITN use exceeds 80% are shown in the bottom row.

**Figure 4—figure supplement 1.**
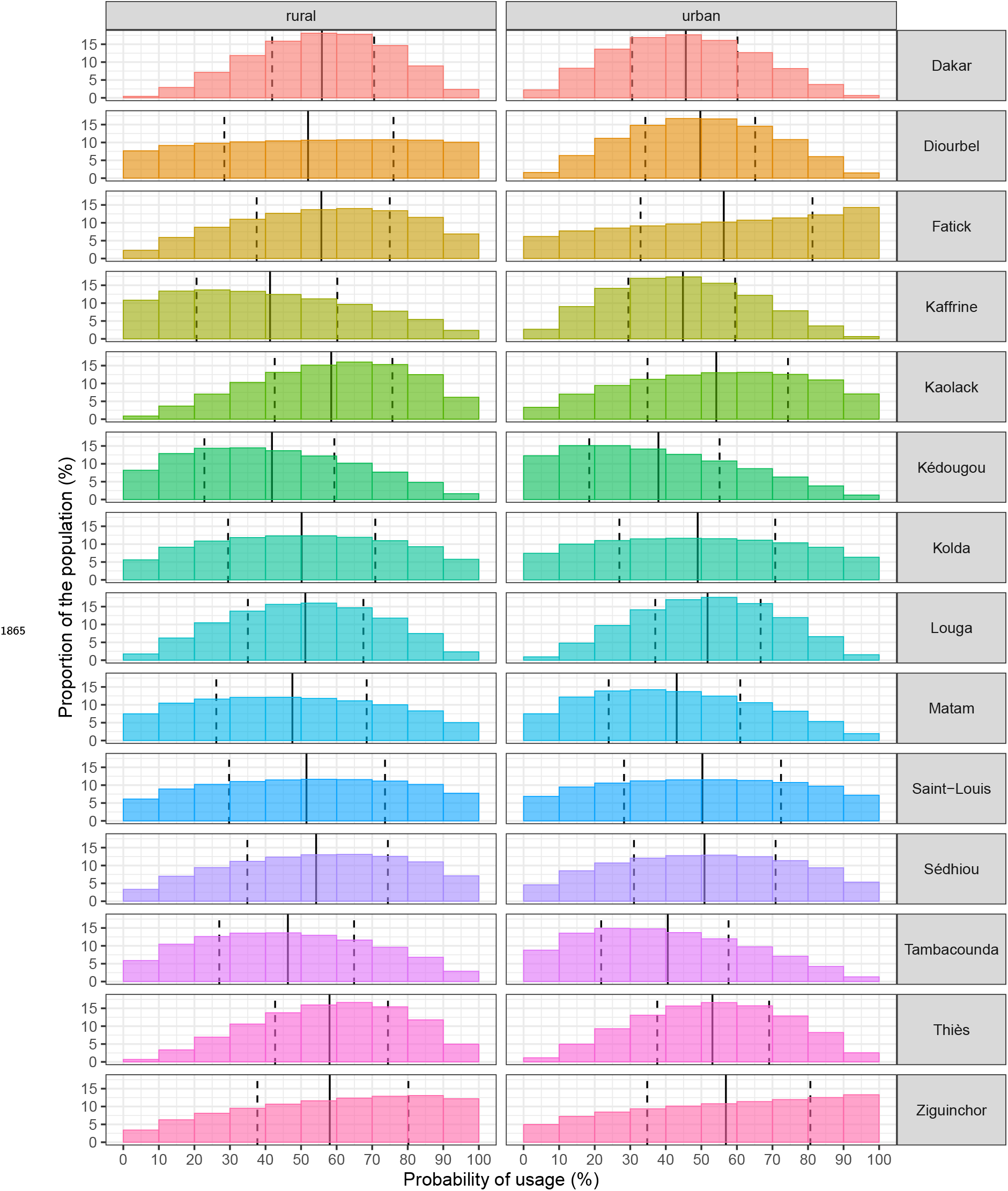
Subnational equity of use in Senegal. All features remain unchanged from figure 4

**Figure 4—figure supplement 2.**
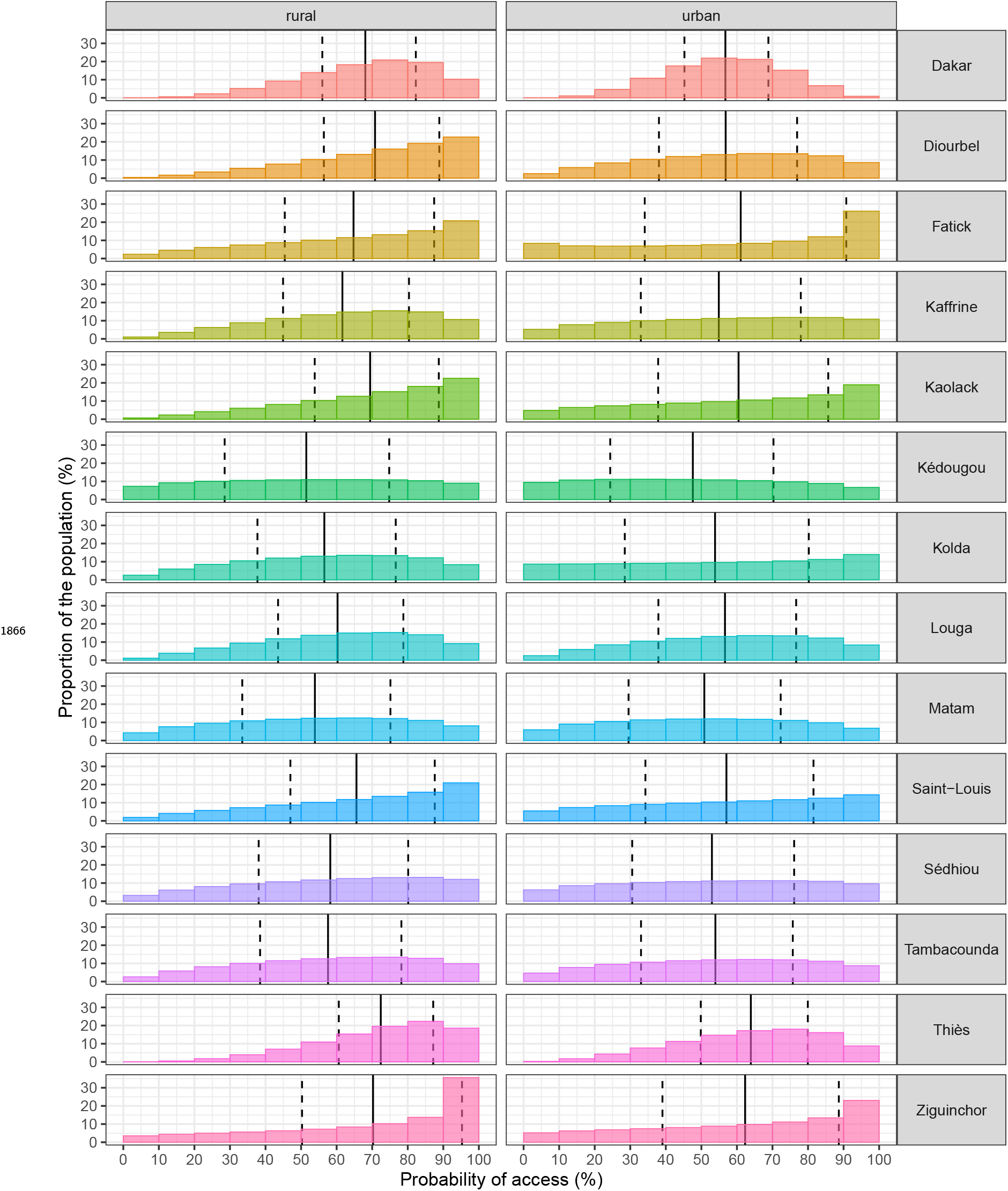
Subnational equity of access in Senegal. All features remain unchanged from figure 4

**Figure 5—figure supplement 1.**
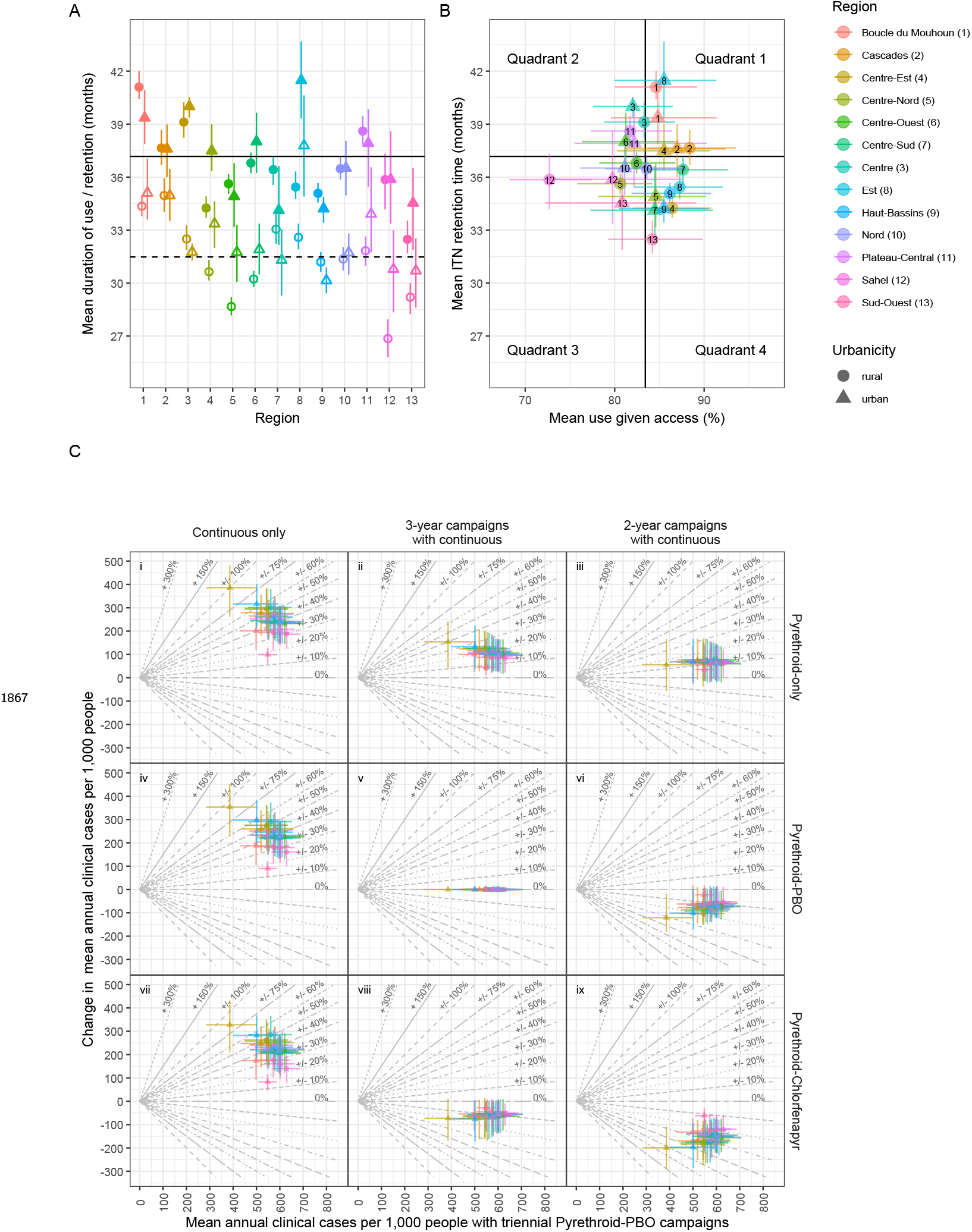
Retention time, use given access and changes in cases for Burkina Faso. All features remain unchanged from figure 5. Note the ordering of colours differs in panel B where numerical labels should be referred to.

**Figure 5—figure supplement 2.**
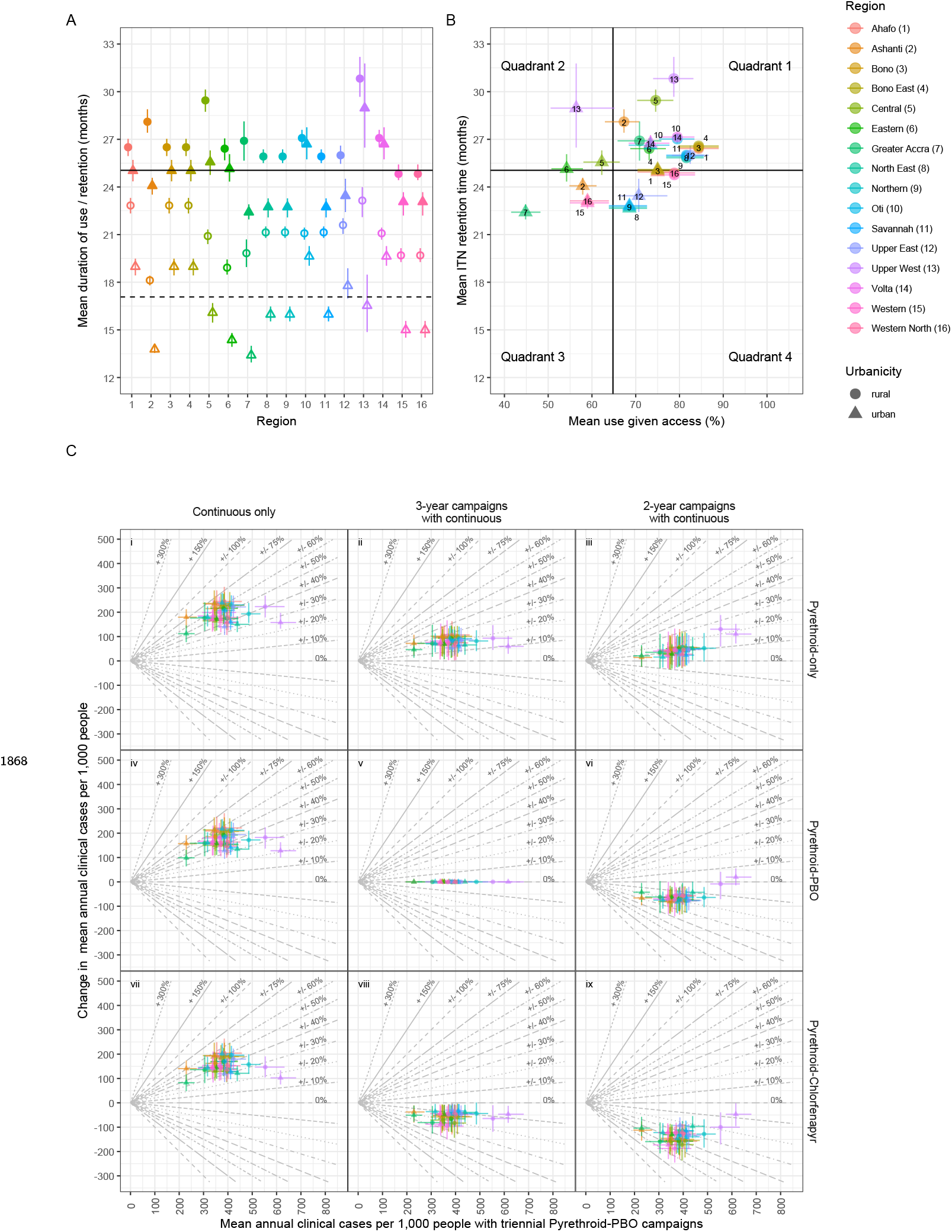
Retention time, use given access, and changes in cases for Ghana. All features remain unchanged from figure 5. In 2019, several administrative regions in Ghana were subdivided: Brong-Ahafo became Bono, Bono East, and Ahafo; the Northern Region was split into Northern, Savannah, and North East; and Volta was divided into Volta and Oti. Retrospective analyses **(A-B)** were conducted using the pre-2019 regional boundaries, while future projections **(C)** used the post-2019 subdivisions. In all subnational regions, ITN retention times and use given access were estimated to be higher in rural than in urban settings.

**Figure 5—figure supplement 3.**
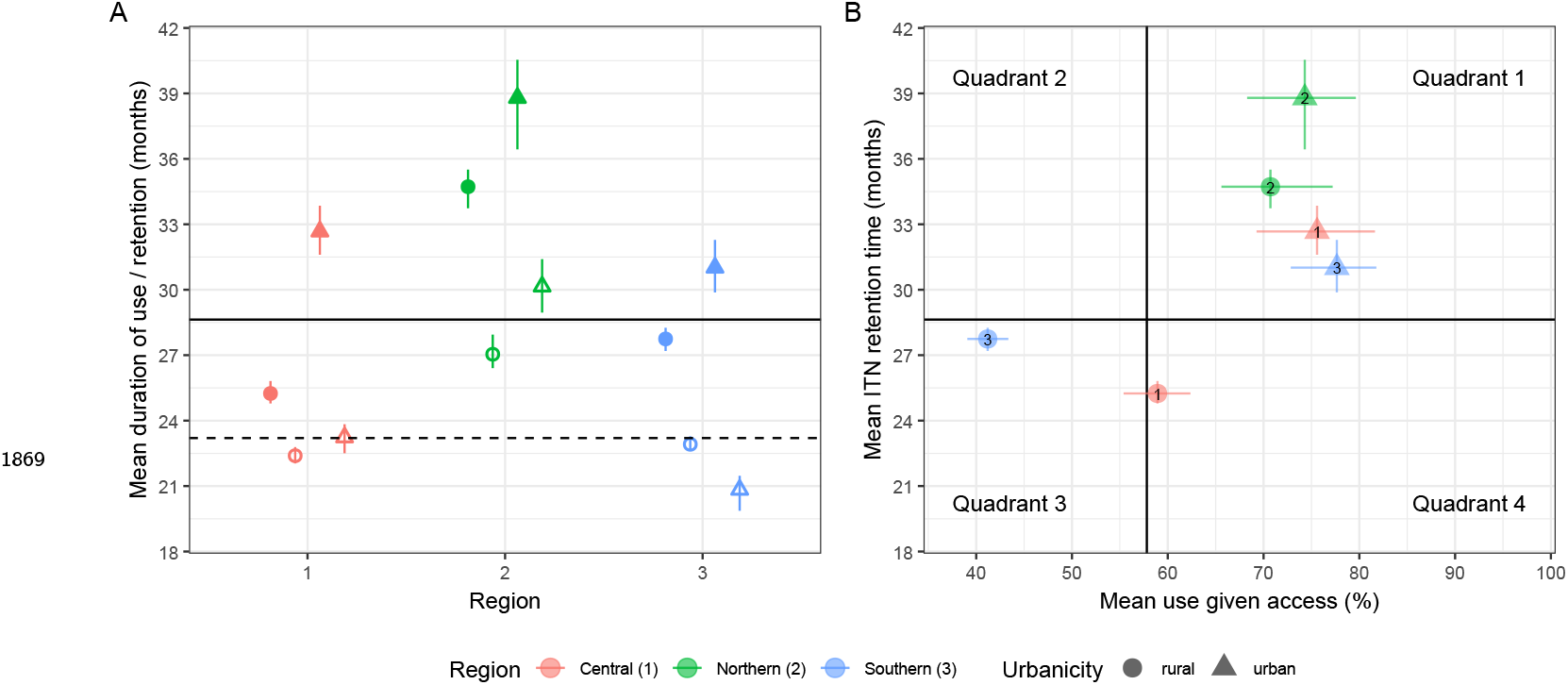
Retention time and use given access for Malawi. All features remain unchanged from figure 5.A. Clinical case projections were not conducted for Malawi due to the lack of intervention data stratified to the administrative-one level, despite DHS surveys being conducted at that scale.

**Figure 5—figure supplement 4.**
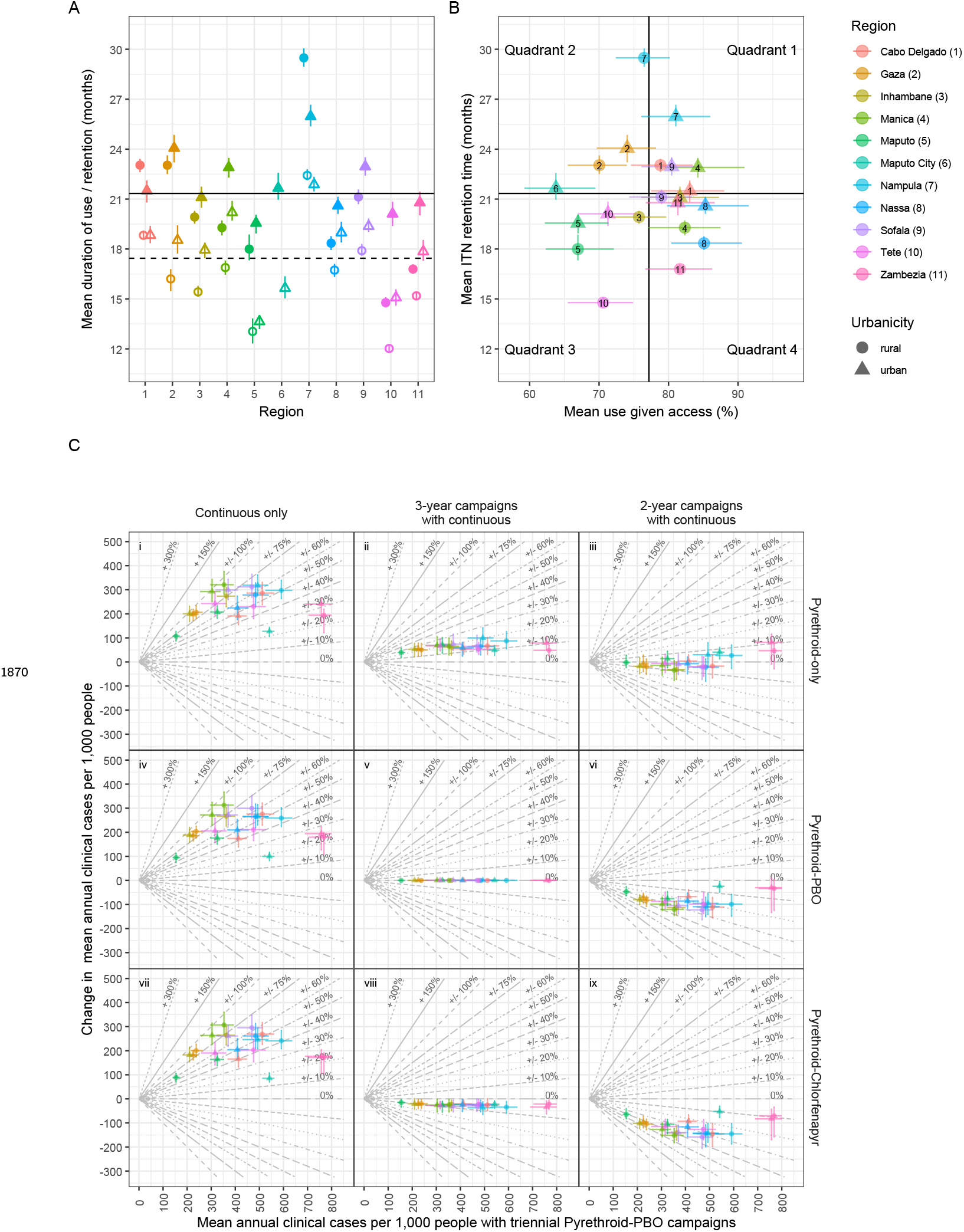
Retention time, use given access and changes in cases for Mozambique. All features remain unchanged from figure 5.

**Figure 5—figure supplement 5.**
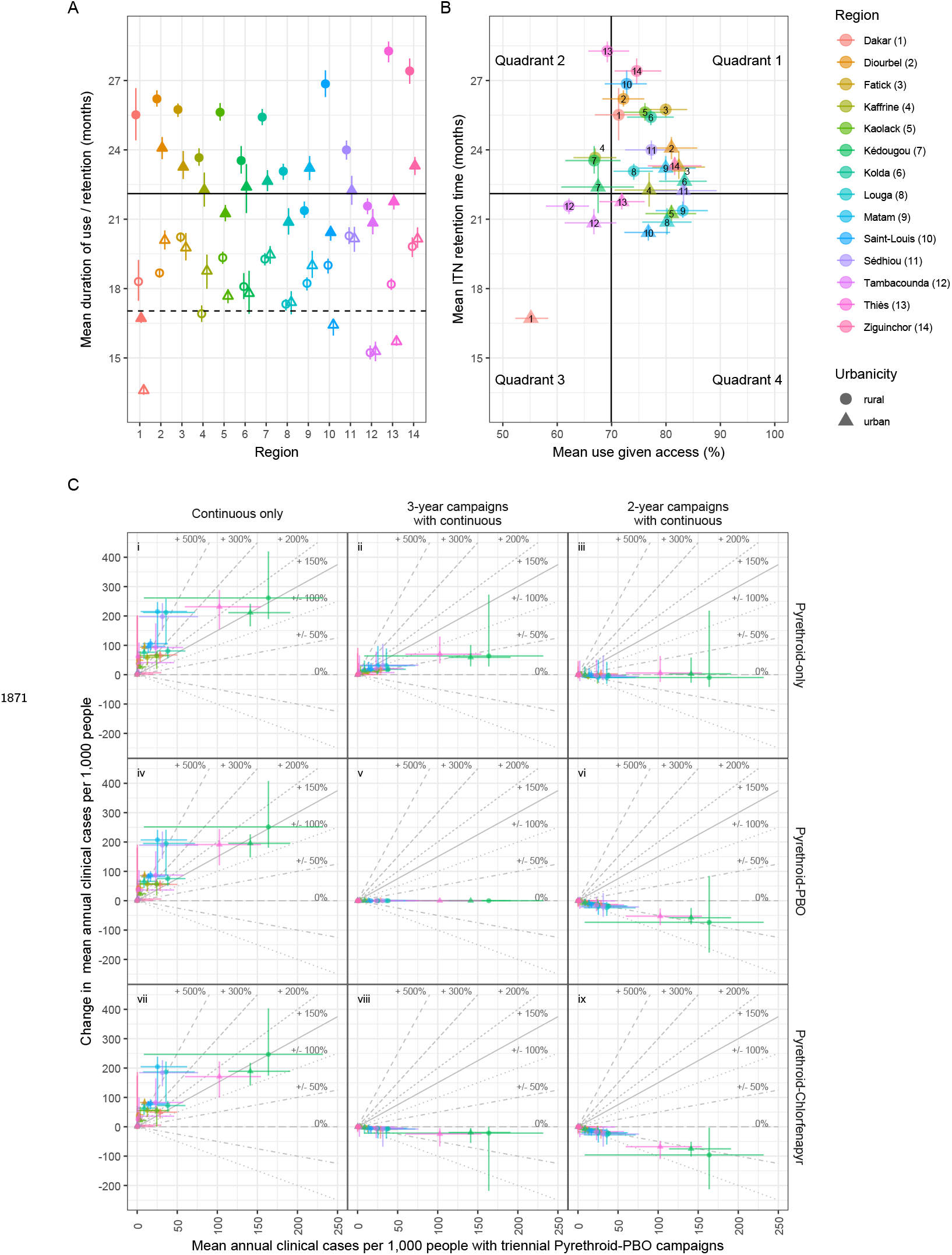
Retention time, use given access and changes in cases for Senegal. All features remain unchanged from figure 5. While use given access was estimated to be broadly similar across urban and rural settings, ITNs were generally estimated to be retained for longer in rural areas;

**Figure 6—figure supplement 1.**
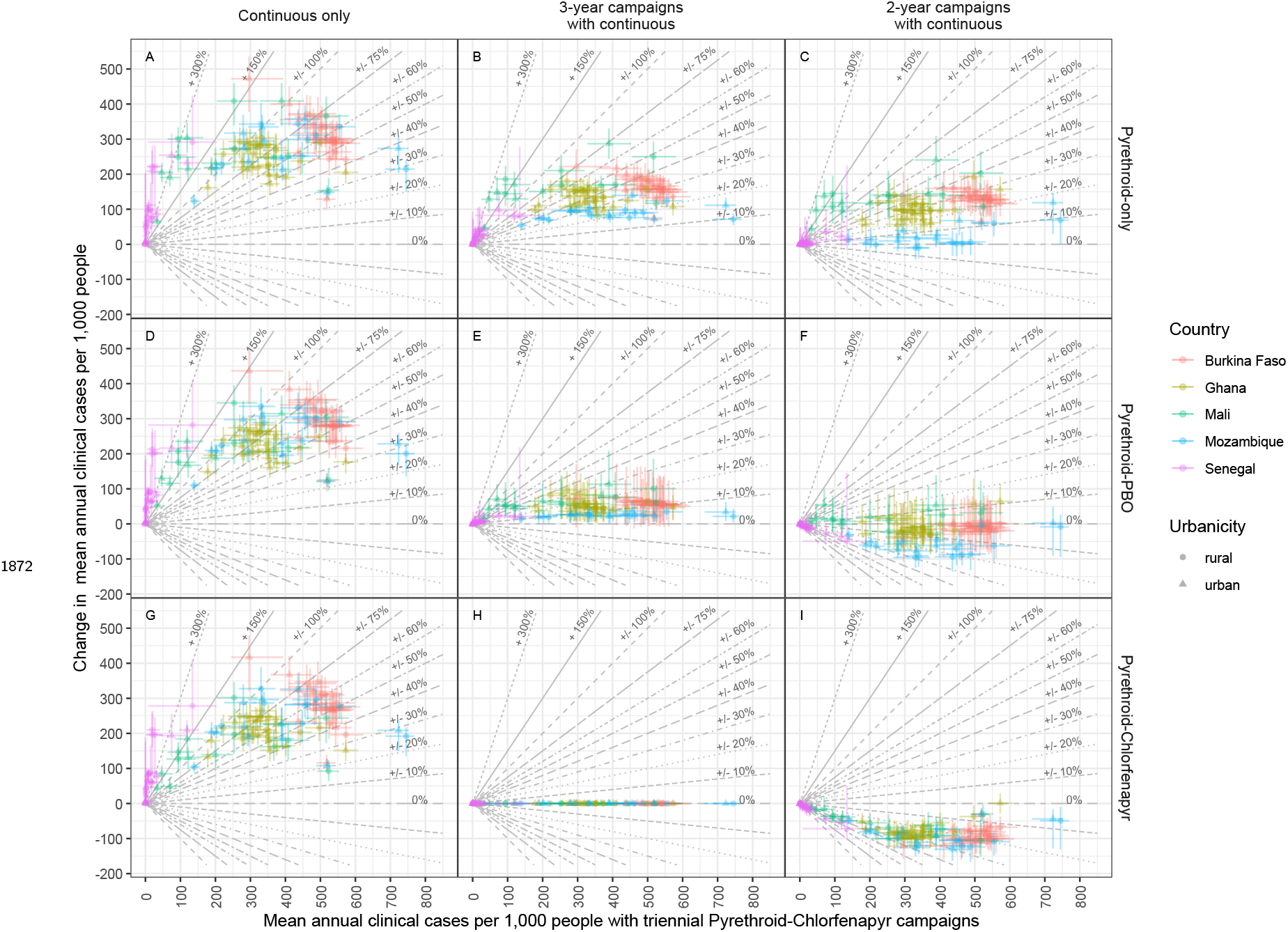
Change in cases vs. cases with triennial pyrethroid-PBO distribution. Points show median estimates of the change in clinical cases following a switch from triennial pyrethroid-chlorfenapyr distribution to alternative strategies against projected clinical cases under the triennial pyrethroid-chlorfenapyr strategy. Figure features otherwise remain unchanged from figure 6.

**Figure 6—figure supplement 2.**
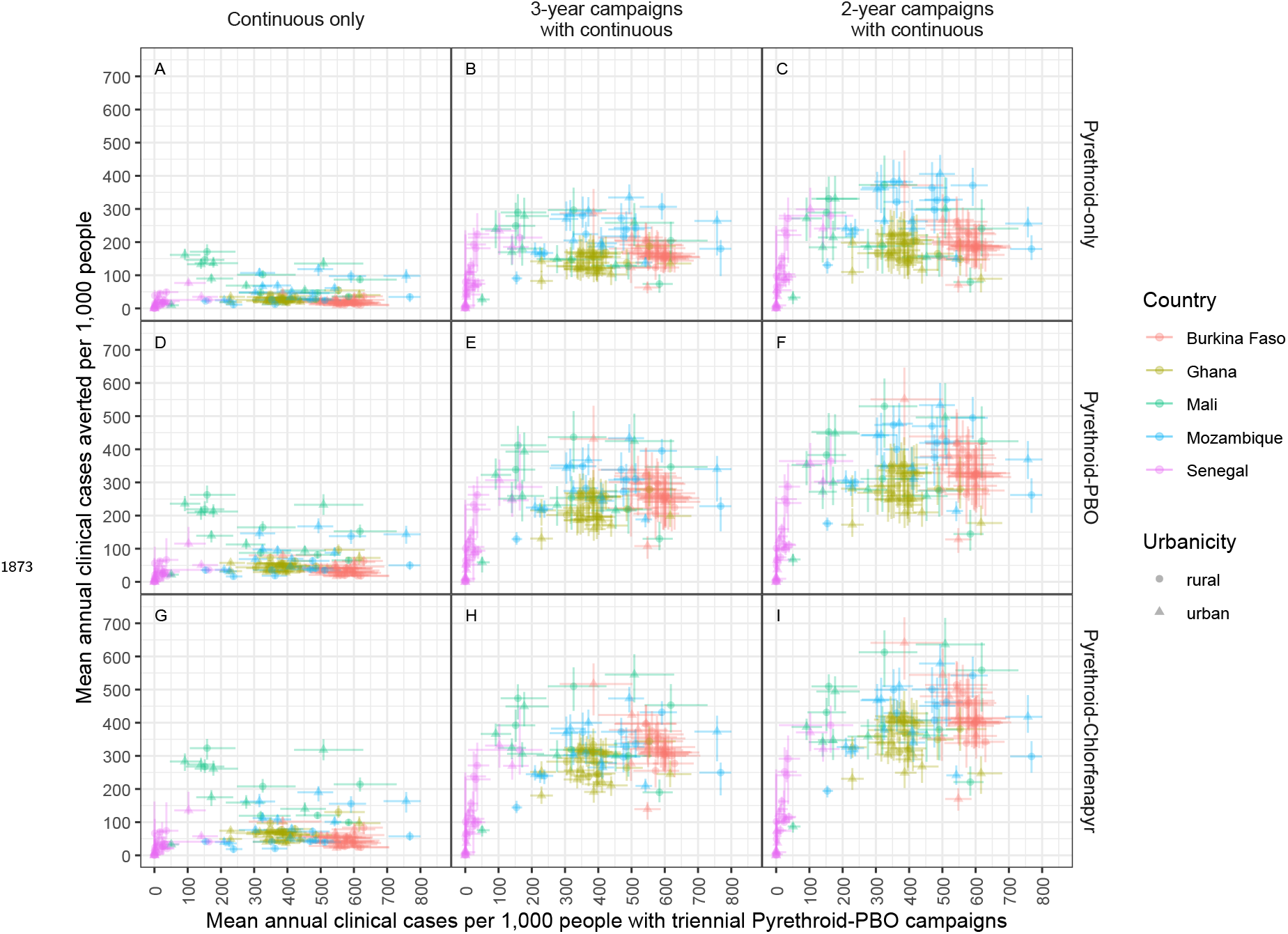
Cases averted under different ITN distribution strategies. Points indicate median estimates of the mean annual clinical cases averted by different ITN distribution strategies, in comparison to ceasing ITN distribution entirely, against clinical cases under a triennial pyrethroid–PBO distribution strategy. Figure features otherwise remain unchanged from figure 6.

**Figure 6—figure supplement 3.**
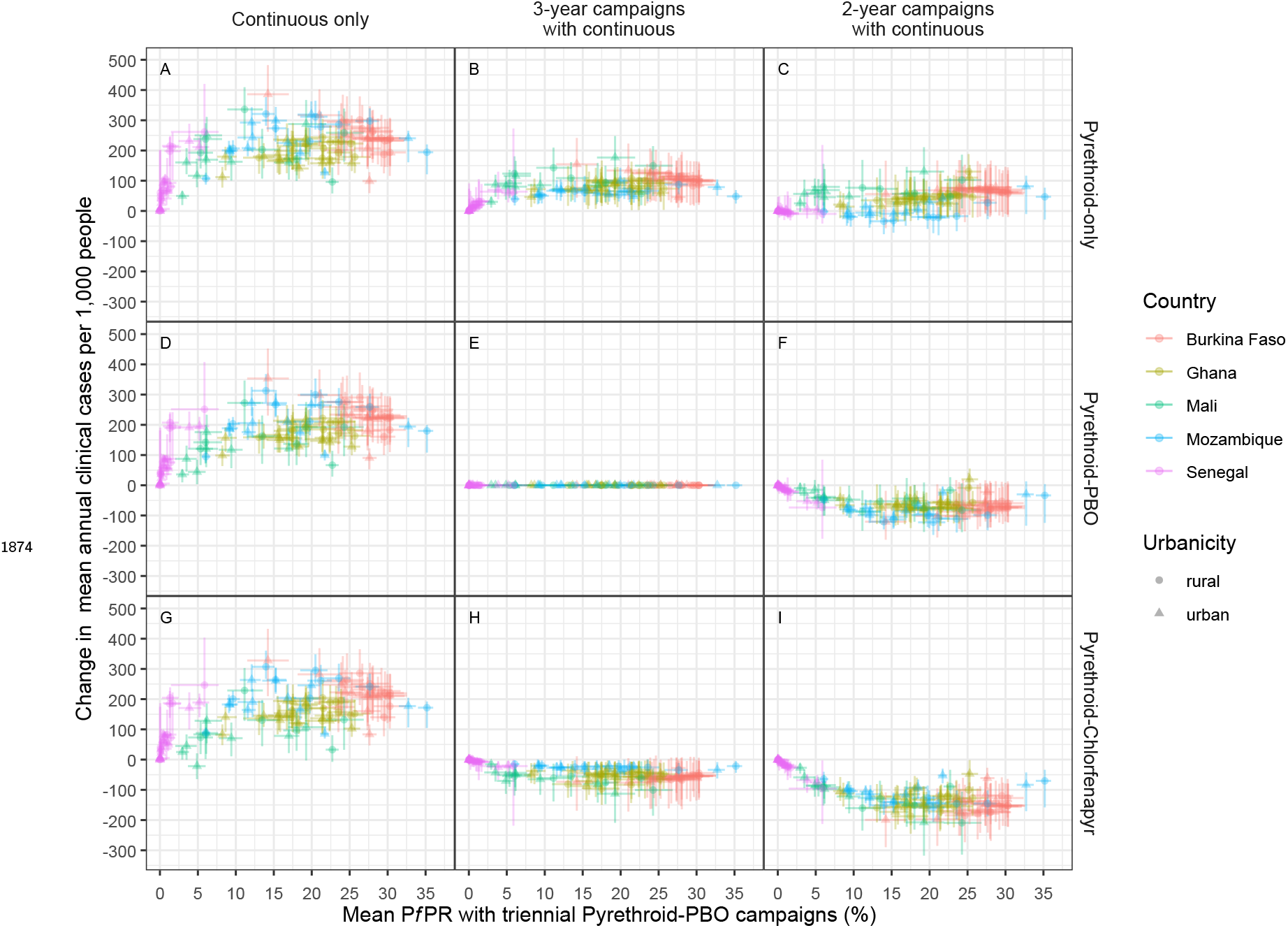
Change in cases vs *Pf* PR with triennial pyrethroid-PBO distribution. Points show median estimates of the change in clinical cases following a switch from triennial pyrethroid-PBO distribution to alternative strategies against all-age *PfPR* under the triennial pyrethroid-PBO strategy. Figure features otherwise remain unchanged from figure 6.

**Figure 6—figure supplement 4.**
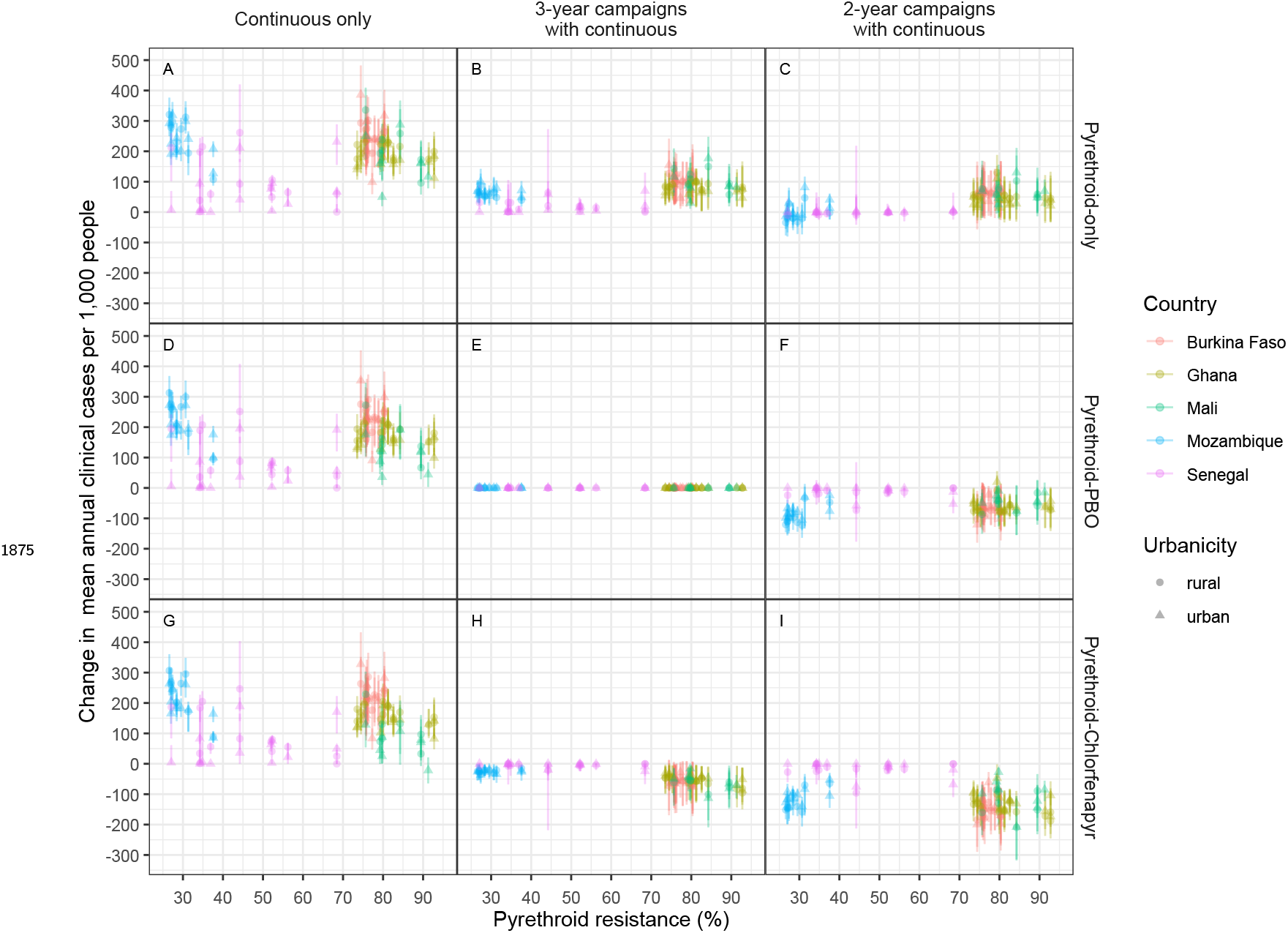
Change in cases vs pyrethroid resistance. Points show median estimates of the change in clinical cases following a switch from triennial pyrethroid-PBO distribution to alternative strategies against pyrethroid resistance. Figure features otherwise remain unchanged from figure 6.

**Figure 6—figure supplement 5.**
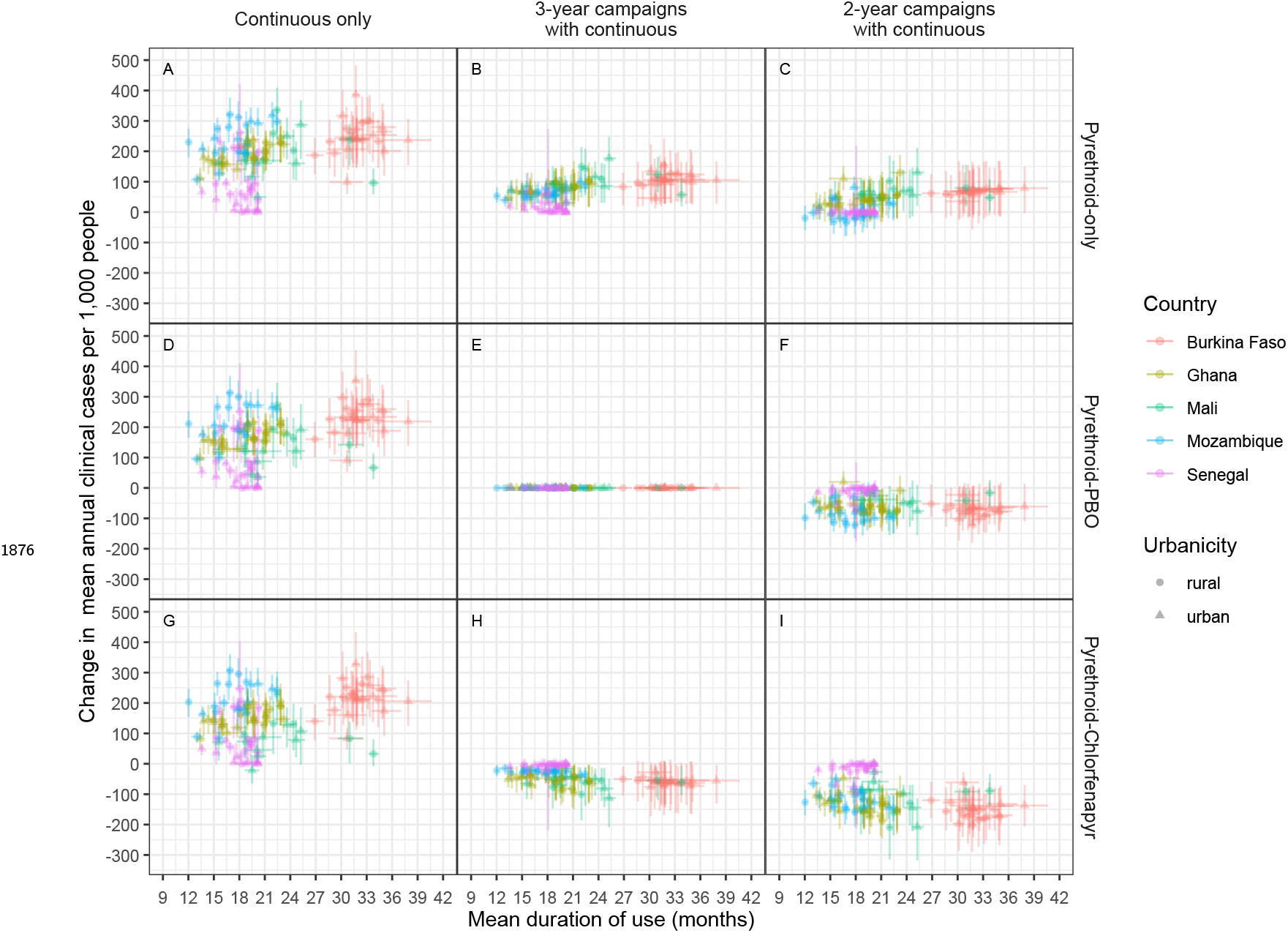
Change in cases vs duration of use. Points show median estimates of the change in clinical cases following a switch from triennial pyrethroid-PBO distribution to alternative strategies against mean duration of use. Figure features otherwise remain unchanged from figure 6.

**Figure 6—figure supplement 6.**
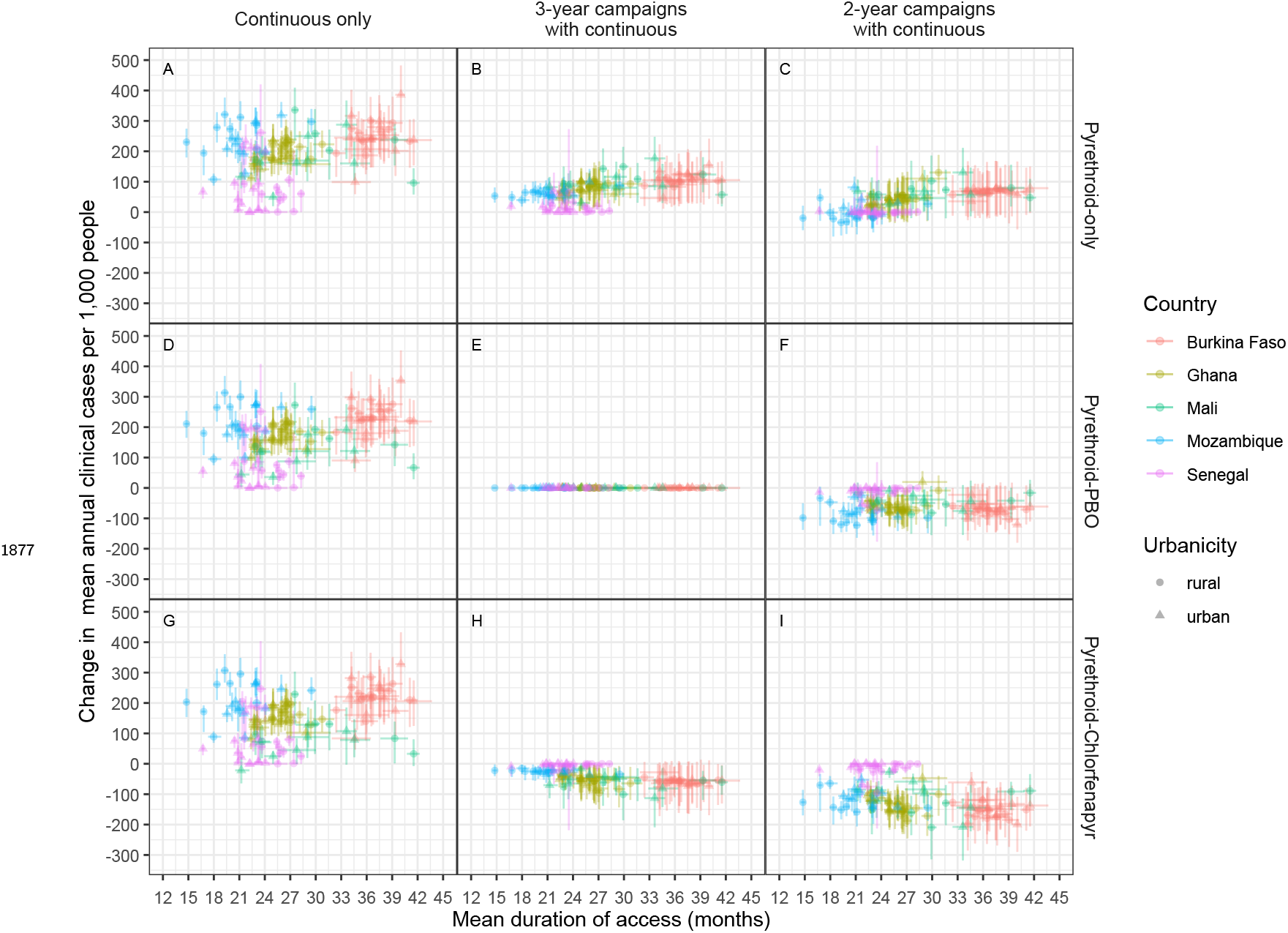
Change in cases vs retention time. Points show median estimates
of the change in clinical cases following a switch from triennial pyrethroid-PBO distribution to alternative strategies against mean retention time. Figure features otherwise remain unchanged from figure 6.

**Figure 6—figure supplement 7.**
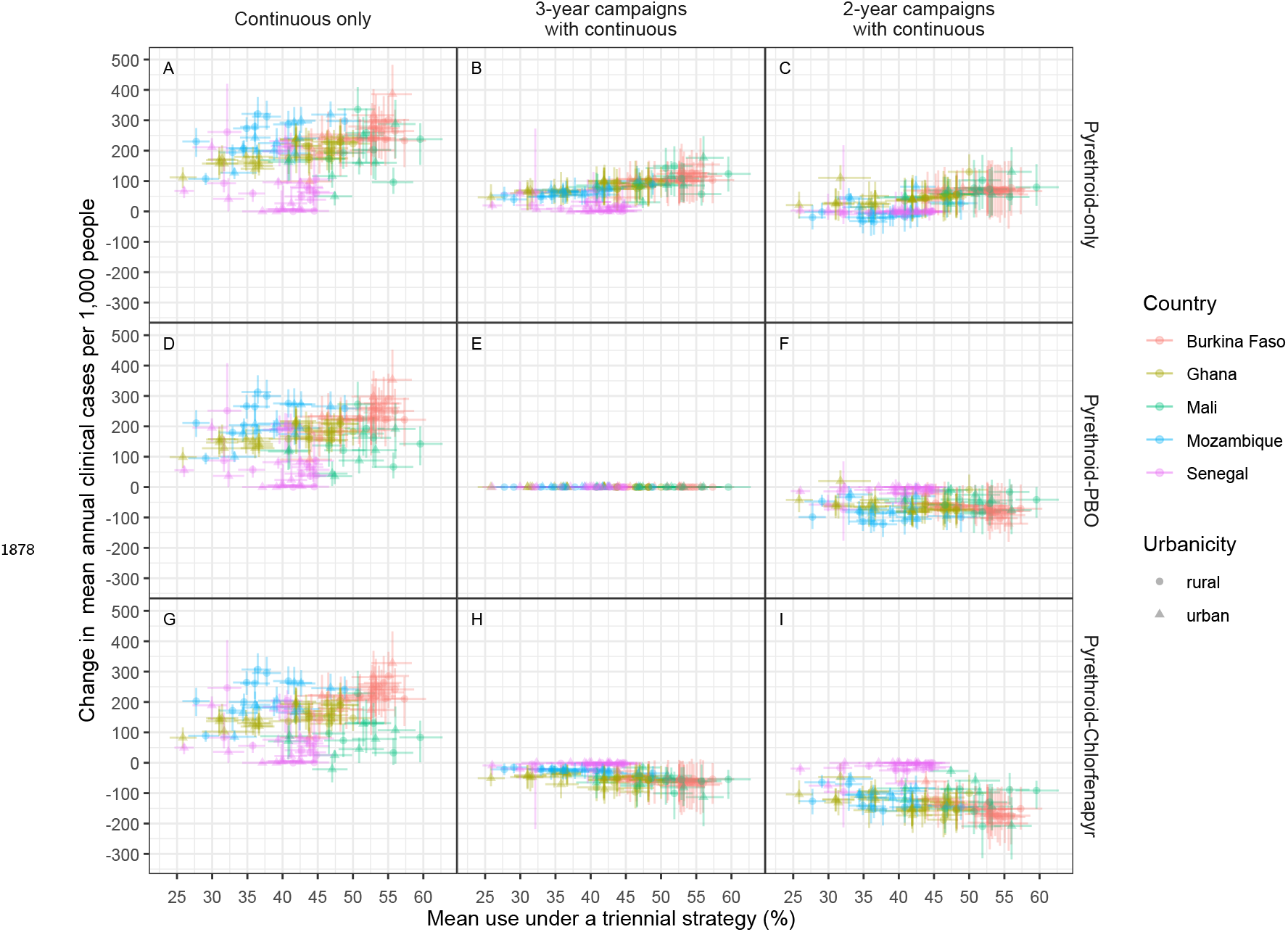
Change in cases vs use. Points show median estimates of the change in clinical cases following a switch from triennial pyrethroid-PBO distribution to alternative strategies against mean use over any triennial distribution strategy. Figure features otherwise remain unchanged from figure 6.

**Figure 6—figure supplement 8.**
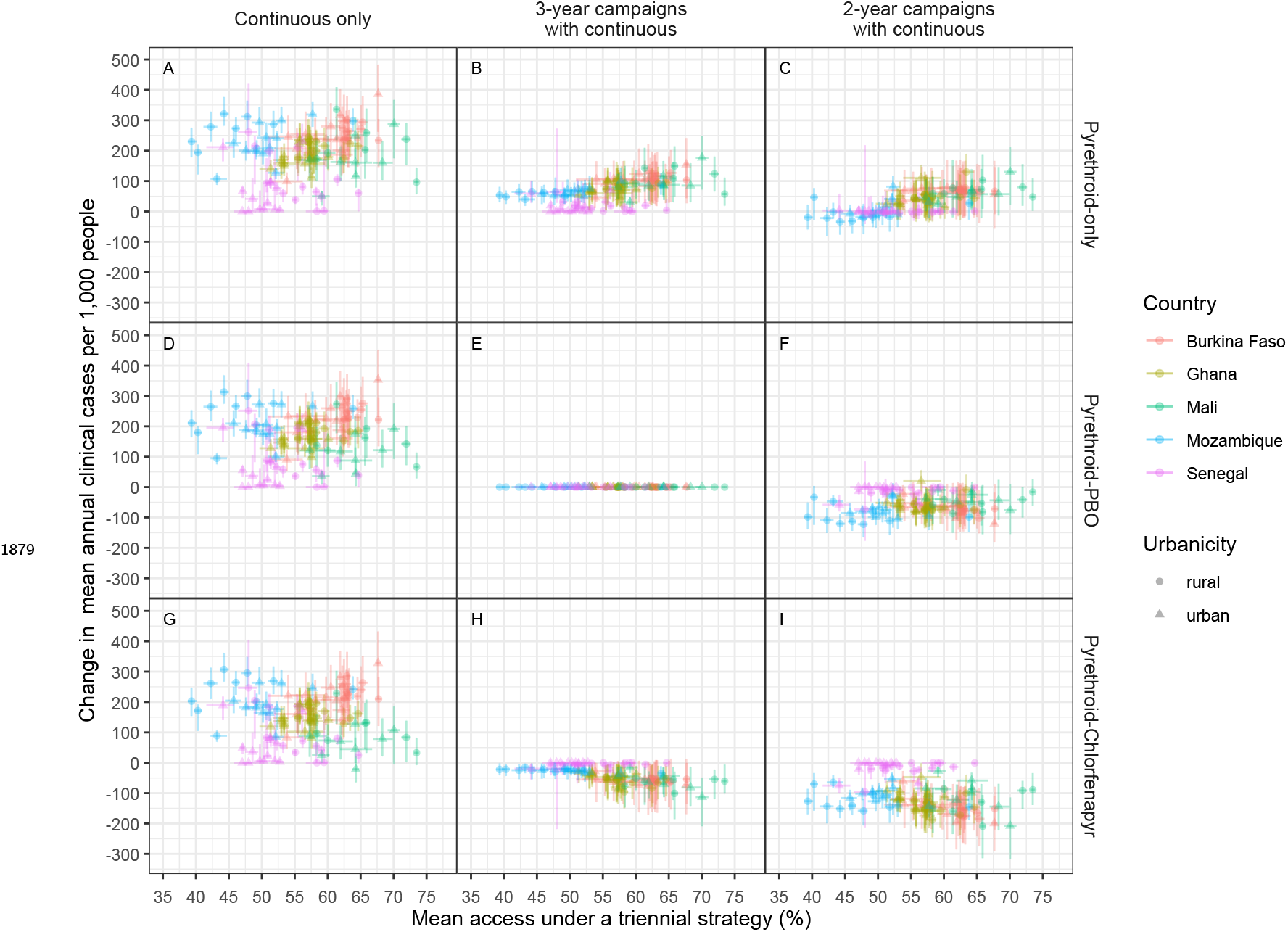
Change in cases vs access. Points show median estimates of the change in clinical cases following a switch from triennial pyrethroid-PBO distribution to alternative strategies against mean access over any triennial distribution strategy. Figure features otherwise remain unchanged from figure 6.

**Figure 6—figure supplement 9.**
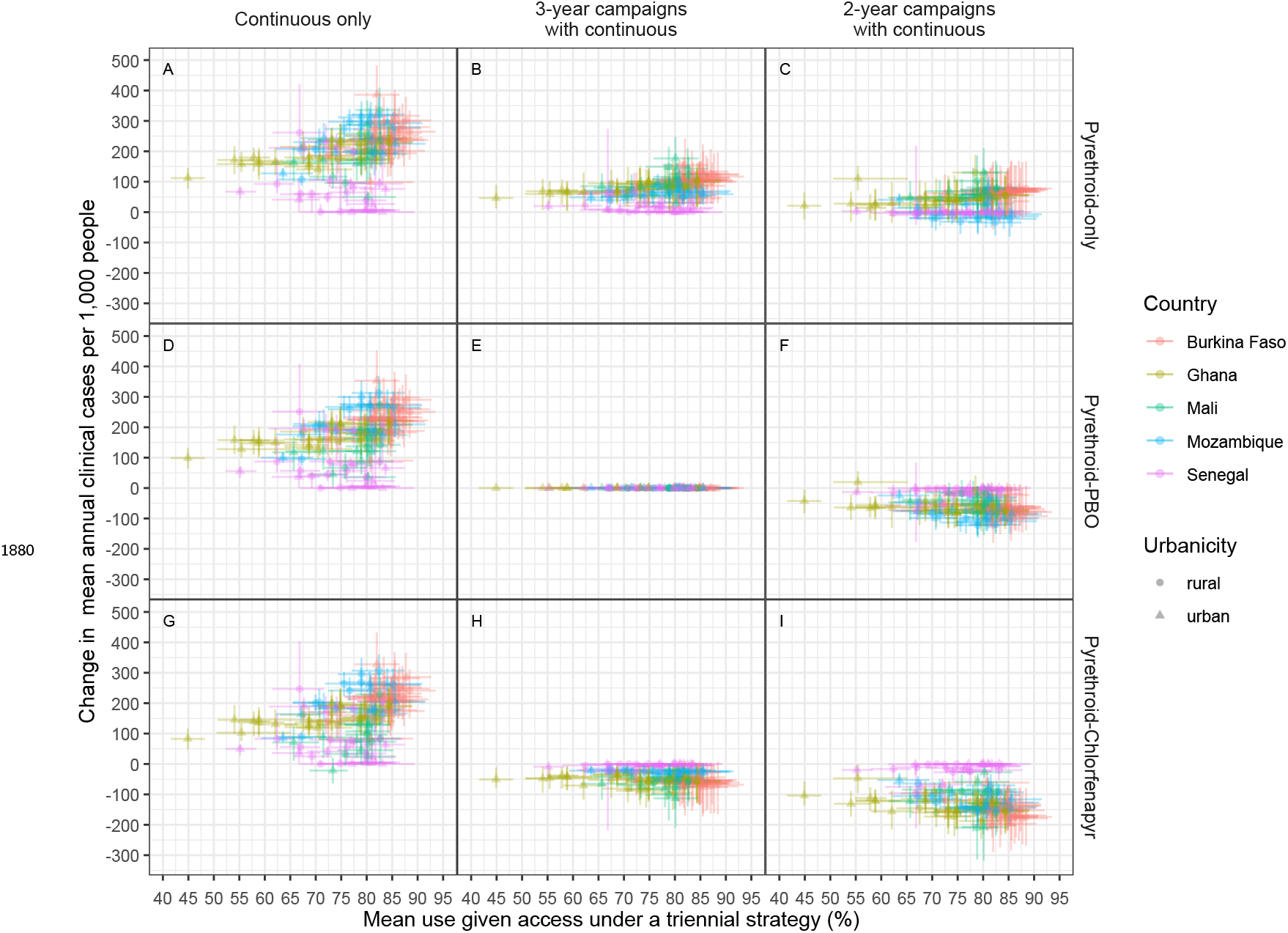
Change in cases vs use given access. Points show median estimates of the change in clinical cases following a switch from triennial pyrethroid-PBO distribution to alternative strategies against mean use given access over any triennial distribution strategy. Figure features otherwise remain unchanged from figure 6.

